# Impaired Pathways of Detoxification and Risk of Alzheimer’s Disease in Humans - Differential Gene Expression Studies via Gene Set Expression Analysis

**DOI:** 10.1101/2025.05.29.25328282

**Authors:** Mark E. McCaulley

**Affiliations:** Independent researcher, unaffiliated; Steamboat Springs, Colorado, US

**Keywords:** Alzheimer’s, dementia, detoxification, RNA-seq, gene expression

## Abstract

**Background:** Alzheimer’s disease (AD) is an all-too-common dementing disease associated with aging, impacting individuals, their families, and society. No specific etiology has been determined despite extensive research over the last century.

**Objective:** Detoxification is a multifaceted systemic process of variable efficacy that eliminates potentially hazardous exogenous and endogenous substances from the body. Multiple genes responsible for detoxification have been previously identified.

**Methods:** Utilizing gene set expression analysis, this paper reports two differential gene expression studies comparing AD and non-AD cohorts, first in a Gene Expression Omnibus (GEO) dataset study and second in a tissue RNA-seq study.

**Results:** We identified several detoxification genes with statistically significant differential expression in AD compared to non-AD samples in both phases of this study.

**Conclusions:** This set of findings suggests a possible etiologic association of impaired detoxification and AD. Subsequent research to follow up on these findings is needed.

**Graphical abstract:** 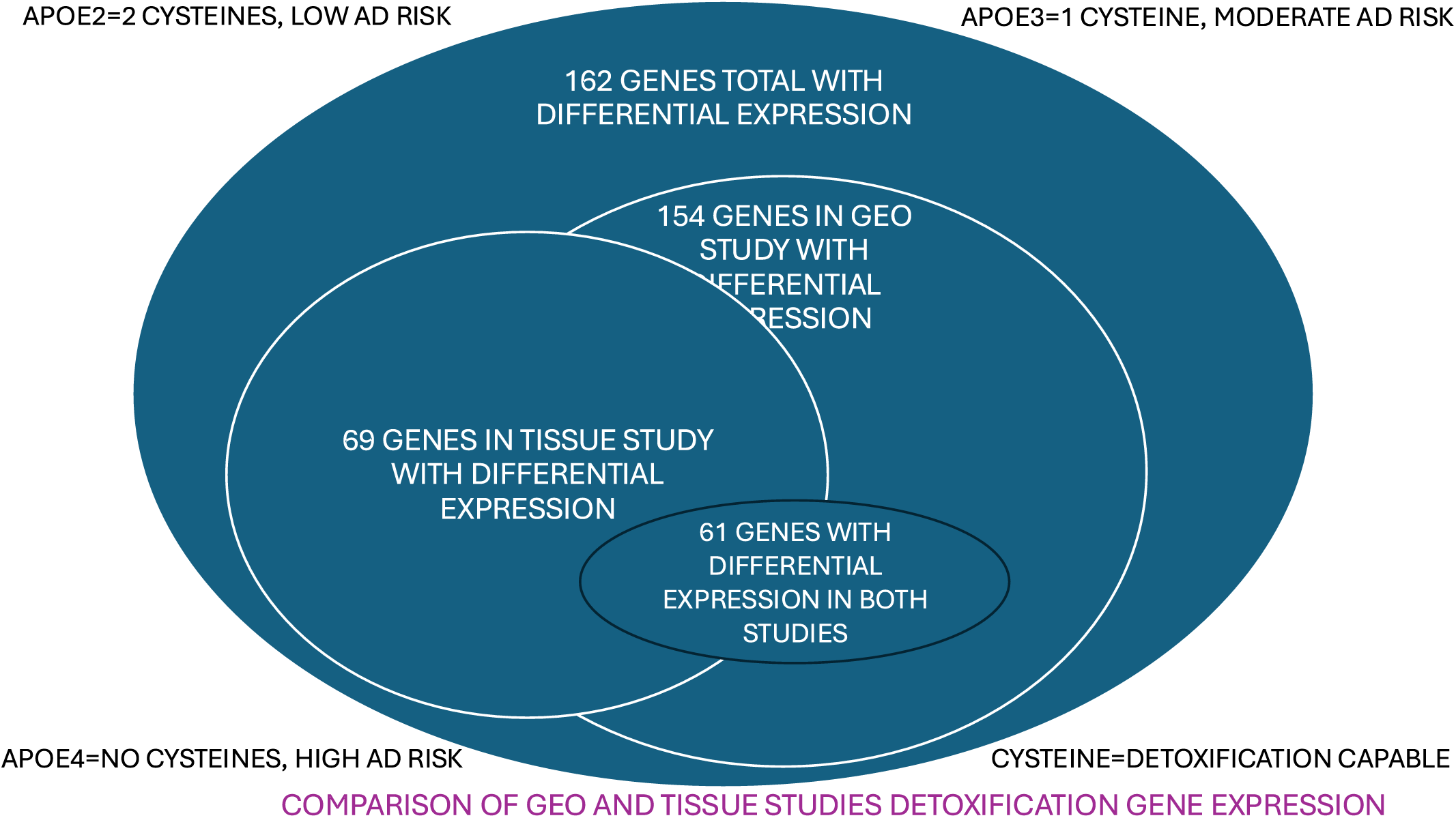

## INTRODUCTION

Alzheimer’s disease (AD) afflicts 6.7 million people in the United States as of 2023 and tens of millions worldwide [1]. More than 11 million caregivers in the US are also impacted. The dominant paradigm in AD for the past several decades has been the amyloid hypothesis. Amyloid beta (AB) is the forty-two amino acid peptide that is the chief component comprising the amyloid plaques that characterize AD.

Although universally present in plaques and tangles of neurons in AD brains, removal of AB using monoclonal antibody treatment has resulted in limited clinical improvement and considerable toxicity, including fatalities [2–5]. Given that there is a large and growing body of evidence that AB is both an antimicrobial protein and a component of the inflammatory response, and as AB is "evolutionarily conserved and is present throughout life in cognitively normal individuals," it is not surprising that the risks and benefits of AB are more nuanced [6]. Why are these abnormal proteins so prominent in AD brains? This paper presents a potential answer to that question.

Exposure to xenobiotics such as metals, pesticides, herbicides, solvents, pharmaceuticals, long lived synthetically produced chemicals, fine grade particulate matter, and endogenously produced substances (mostly hormones and neurotransmitters) is ubiquitous [7, 8]. A 2024 paper by Dhapola et al. presents a comprehensive compilation of environmental toxins associated with AD [9]. Optimally, these various substances are ultimately broken down and/or eliminated from the body, though some are long lived, and some may be permanent. Multiple genetically determined pathways for detoxification and elimination of these substances have been identified. The effectiveness of these pathways varies, depending on the expression of the pertinent genes, resulting in each human having an innate capability for detoxification [10, 11]. This capability is specific to the individual and the substance being detoxified. Research on the potential for the variable capability of these pathways being associated with any disease is limited. The present paper presents research documenting an association of detoxification gene expression and risk of AD.

### Existing evidence of impaired detoxification in AD

#### APOE

APOE is well known for being associated with AD risk[12, 13]. An apolipoprotein, APOE has three isoforms, annotated as APOE2, APOE3, and APOE4. We all have two of these in our genomes, one from each parent. APOE4 is associated with highest risk of AD, APOE2 with lowest risk, and APOE3 with intermediate risk. Structurally, APOE has 299 amino acids. All 3 isoforms have nearly the same amino acid sequence, except for positions 112 and 158. APOE2 has cysteine amino acids at both positions, APOE4 has arginine amino acids at both positions, and APOE3 has a cysteine amino acid at position 112 and an arginine at position 158. Since sulfur containing amino acids such as cysteine have detoxification properties, a potential explanation emerges for the difference in AD risk associated with the specific APOE status of individuals [14, 15], related to whether cysteine or arginine is present at positions 112 and 158 in the APOE isoforms.

#### Glutathione

The glutathione system is prominent in the defense of the body from oxidative stress and plays a major role in detoxification and excretion. At least 21 genes in the glutathione detoxification system are currently recognized as active in limiting susceptibility to toxic exposures. The body synthesizes glutathione from cysteine, which may be the most significant function of cysteine in the body. A variety of genetic variations in the glutathione system are associated with greater or lesser vulnerability to toxic substances [16–18].

Given the person-to-person variability in expression of detoxification genes, the subtle clues pointing to a possible association of detoxification in AD, and despite the lack of previous similar studies, I present herein an investigation of a new hypothesis: differential expression of detoxification genes may be associated with AD.

## RESULTS

Analysis of RNA-seq data from both phases of this evaluation revealed significant differential gene expression (DGE) in AD versus non-AD cohorts in multiple detoxification genes, with moderate to marked statistical significance, substantially supporting the hypothesis, that impaired routes of detoxification may impact the risk of AD. Resources of the Gene Expression Omnibus (GEO), a repository of gene expression data, were utilized in Phase 1. Analysis of the noted datasets in Phase 1 was done via the GEO2R platform, associated with the GEO data repository, and further described in materials and methods, below. Analysis of the Phase 2 results was done by LC Sciences, the lab that performed RNA-sequencing on purchased tissue in Phase 2, also described in materials and methods. In both phases, gene set expression analysis was performed, utilizing genes identified as genes of interest (GOI). The process of creating the GOI list is also described in materials and methods, below. All the significant DGE values reported below are in genes from the GOI list, and not further annotated in this manuscript. Throughout this manuscript, significant differential gene expression is defined as adjusted p-values less than or equal to 0.05 in the Phase 1 GEO study, or in the Phase 2 tissue study, Q values less than or equal to 0.05. In the gene expression tables, log2FoldChange refers to the number of gene copies detected, a positive number indicating increased expression, and a negative number indicating decreased expression.

### Phase 1

#### GEO study

DGE was sought in several publicly available datasets. GEO datasets used for the purpose of evaluating the present hypothesis are: GSE104704, GSE159699, GSE153873, GSE173955, GSE211993, GSE227221, and GSE190125.

The initial analysis utilized the GSE104704 dataset from an April 2018 publication [19], using human lateral temporal lobe samples, from AD (12, mean age 68), young control (CTL) (8, mean age 52) and old CTL (10, mean age 68) subjects (gender not specified). The analysis was performed for DGE in AD versus young CTL, AD versus old CTL, and AD versus CTL (combining young and old controls).

In AD versus young, 25 genes displayed significant DGE, 10 having decreased expression, and 15 having increased expression. In AD versus old, 16 genes were significantly differentially expressed, 11 with decreased expression, and 5 with increased expression. 19 GOI genes were significantly differentially expressed in the AD versus CTL samples (combining old CTL and young CTL), 8 with decreased expression, and 11 with increased expression.

Supplementary Tables S3-S5 summarize the results of DGE comparisons for GSE104704 AD versus young, AD versus old, and AD versus control.

The next analysis was of GSE159699 from October 2020 [20] using human lateral temporal lobe samples. DGE data in GSE159699 is identical to that in GSE104704; it is summarized in Supplementary Tables S6-S8.

The third dataset to be analyzed, GSE153873, from the same October 2020 publication as GSE159699, utilizing human hippocampus with External RNA Controls Consortium (ERCC) spike-in as control, was annotated into AD, young CTL, old CTL, and CTL (combining young CTL and old CTL).

### Metadata

- 12 AD (mean age 68, range 61-79, one female, and 11 males) not further subdivided into young or old groups.

-8 young CTL (mean age 52 range 42 to 60, 8 males and 1 female)

-10 old CTL (mean age 68, range 61 to 73, 9 males and 1 female).

The AD versus young CTL comparison had 27 GOI with significant DGEs, 10 with decreased expression and 17 with increased expression. The AD versus old CTL comparison had 19 GOI with significant DGEs, 10 with decreased expression, and 9 with increased expression. AD versus CTL had 19 GOI with significant DGEs, 6 with decreased expression, and 13 with increased expression. Supplementary Tables S9-S11 summarize the GSE153873 results.

These first three datasets, shown above, are very similar, though reported separately on two different dates in the GEO system. Datasets GSE104704, GSE159699, and GSE153873 report metadata from the same samples, though grouped differently. Author lists were mostly the same authors with minor changes. Running the GEO2R platform yielded results that were also nearly identical in the first two datasets, although there were minor differences. The GSE153873 dataset, though distinct from GSE159699, derived from the same manuscript. The ERCC "spike in" utilized in GSE153873 likely accounts for the differences in differential gene expression found in the present analysis of that study. Analyses of all three datasets were included in this report due to the comparable DGE results in the analyses of those datasets, adding support to the validity of the GEO2R platform as reported here.

GSE173955 is the next dataset reported in this GEO DGE AD analysis, from a publication in December 2021 [21]. The tissue used was human hippocampus.

### Metadata annotation

5 female samples in the AD group, age range 84 to100

5 female samples in the CTL group, age range 72 to 87

3 male samples in the AD group age range 83 to 99

5 male samples in the CTL group age range 74 to 83

38 GOI genes were differentially expressed in this DGE analysis, with 14 genes showing decreased expression, and 24 genes showing increased expression. GSE173955 results are summarized in Supplementary Table S12.

GSE211993 is a GEO dataset from a publication in March 2023 [22] reporting results of a novel DGE evaluation comparing human induced pluripotent stem cells (hiPSC) carrying either the familial Alzheimer’s Disease (fAD)-linked A79V PSEN1 mutation, or the fAD-linked L150P PSEN1 mutation, with gene corrected isogenic control (CTL), modified via precise CRISPR/Cas9 gene editing. The hiPSCs were incubated for 7 days prior to RNA-seq. This ingenious design eliminates the ambiguities inherent in most intergroup comparisons of diseased and control populations due to unavoidable subjectivity in determining which subjects have and which are free of the condition under investigation.

The first part of the GSE211993 dataset analysis is the L150P PSEN1 mutation analysis. In this analysis, 84 GOI genes showed significant DGE, with 39 of those genes showing decreased expression and 45 showing increased expression, many with remarkable adjusted p-values.

In the second part, the A79V mutation analysis, 76 genes from the GOI list were found to exhibit significant DGE, 45 of which showed decreased expression, with 31 showing increased expression. Many of these adjusted p-values also exhibited marked significance.

The third part of the GSE211993 dataset analysis is the combined A79V and L150P mutations analysis compared with gene corrected CTL. Significant DGE was found in 65 genes, with 43 genes showing decreased expression and 22 showing increased expression.

Results of the GSE211993 analysis are summarized in Supplementary Tables S13- S16.

GSE227221, associated with a publication from September 2023 [23], reports an investigation into microglial organization and factors impacting gene expression. The well-known association of microglia and AD adds pertinence to this study. In this dataset, modified non-diseased microglial cells were incubated with AB 1-42 fibrils, for zero hours (CTL), 4 hours, 24 hours, and 96 hours. DGE in GOI genes via RNA-seq was measured after the incubation period. Subsequently, after 4 hours incubation, significant DGE was found in 47 GOI genes, with decreased expression in 26 genes and increased expression in 21 genes. After 24 hours incubation, significant DGE was found in 75 GOI genes with decreased expression in 45 genes and increased expression in 30 genes. After 96 hours incubation, significant DGE was found in 62 GOI genes with decreased expression in 37 genes and increased expression in 25 genes. In GSE227211 adjusted p-values were marked in several of the DGEs and showed both increased expression in some genes and decreased expression in others. In the 24h incubation analysis the most extreme adjusted p-value was actually zero, with a positive log2FoldChange finding indicating increased expression. Although GSE227221 is not an AD versus control dataset, the value of discerning a potentially important role of AB in regulation of detoxification gene expression is obvious. Supplementary Tables S17- S19 summarize the findings from GSE227221.

GSE190125 is a GEO dataset from an April 2023 publication[24] investigating Down’s syndrome (trisomy 21), reporting RNA-seq data from the Crnic Institute Human Trisome Project, utilizing 304 trisomy 21 samples in comparison to 96 CTL samples. This dataset stands out in its utility due to its large size and the documented elevated risk of early onset AD in trisomy 21 as well as the clear segregation of the trisomy 21 samples from the non-trisomy 21 samples.

Analysis of GSE190125 reveals 81 GOI with significant DGE, 30 with decreased and 51 with increased expression, many with highly significant adjusted p-values. How to incorporate the strikingly positive findings from this trisomy 21 study into a theoretical construct of AD will await further research into these findings. Supplementary Tables S20-S21 summarize data from GSE190125.

### Phase 2

#### Tissue study

RNA sequencing (RNA-seq) was carried out on purchased, bio-banked liver tissue, comparing AD and control samples and calculating DGE data. Gene differential expression, transcript differential expression, known long non-coding RNA (lncRNA) differential expression, and novel lncRNA differential expression were calculated for each of the AD/control/male/female categories. Significance is expressed in adjusted P values and Q values, the latter adjusting for the false discovery rate and having fewer false negatives, and the former controlling the family-wise error rate with fewer false positives. Adjusted P and Q values generally identify the same genes as being significant. Given that there may be several splicing isoforms transcripts found in one gene, the result tables report more transcripts than genes.

Considerable to marked differential gene expression in the genes of interest was found in multiple subcategories. As in the phase 1 GEO datasets study, in the tissue study the DGE data show a mix of increased and decreased expression. Supplementary Table S22 summarizes GOI DGE data in the tissue study (8 pages), displaying significant DGE found in the subcategories of this phase of our study.

Note the 13 categories of long noncoding (lnc) RNA, 6 of which were novel, in the Supplementary Table S22.

**Bar Charts** The figures below are presented in a format that may be enlarged by use of the view and zoom tools.

**Figure 1.**
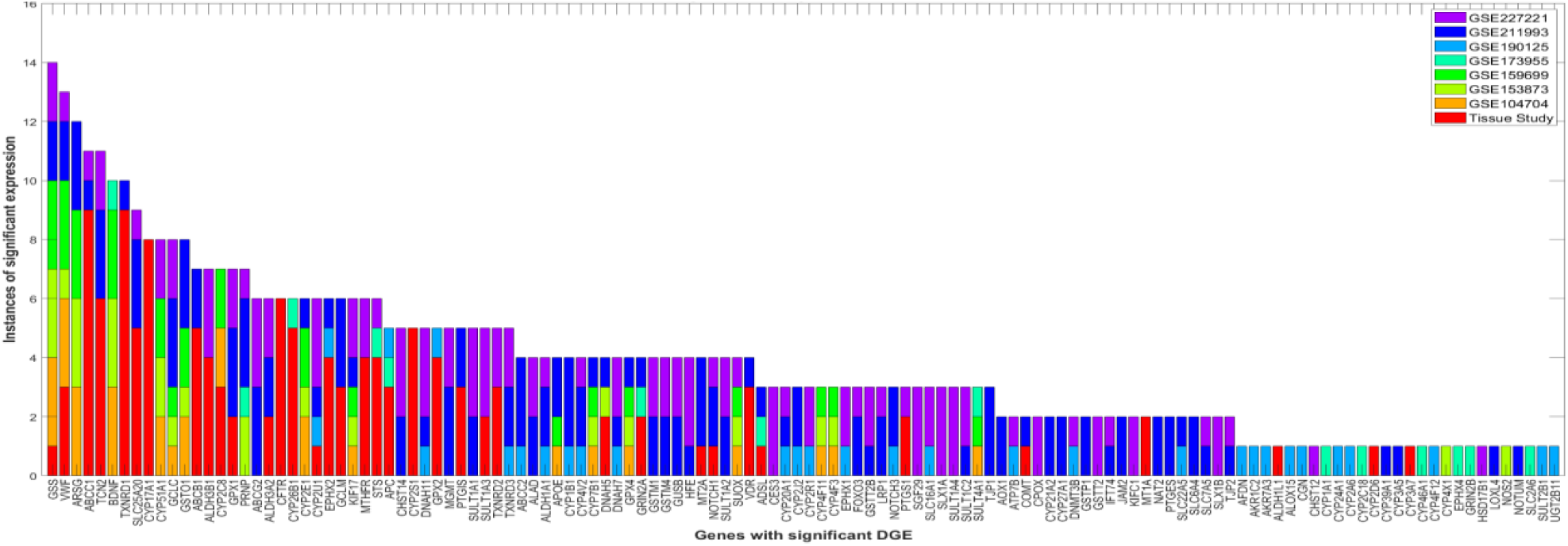
Genes with significantly decreased expression in AD vs. control in combined studies. displays data from the GEO study and tissue study combined, graphically displaying genes with decreased differential expression in AD groups versus controls.

**Figure 2.**
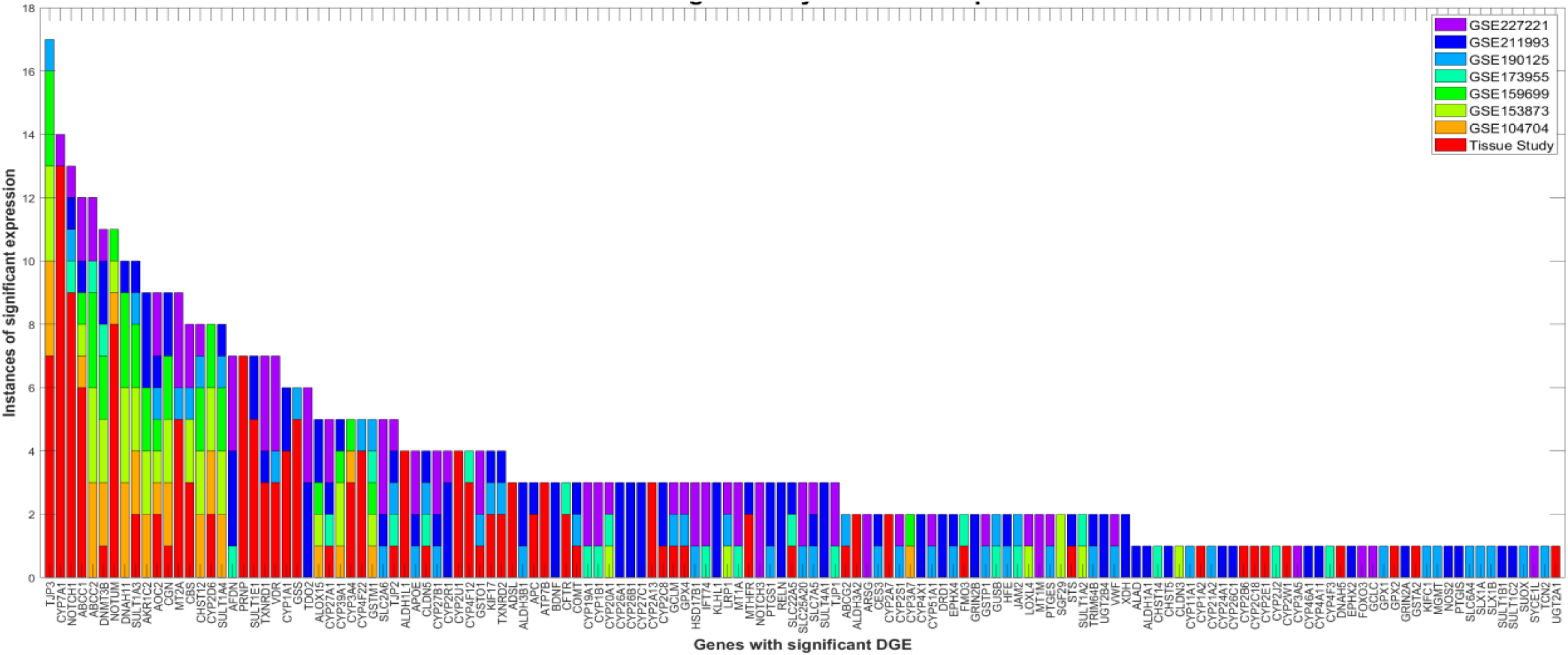
Genes with significantly increased expression in AD vs. control in phase 1. displays data from the GEO study and tissue study combined, showing genes with increased differential expression in AD groups versus controls.

**Figure 3.**
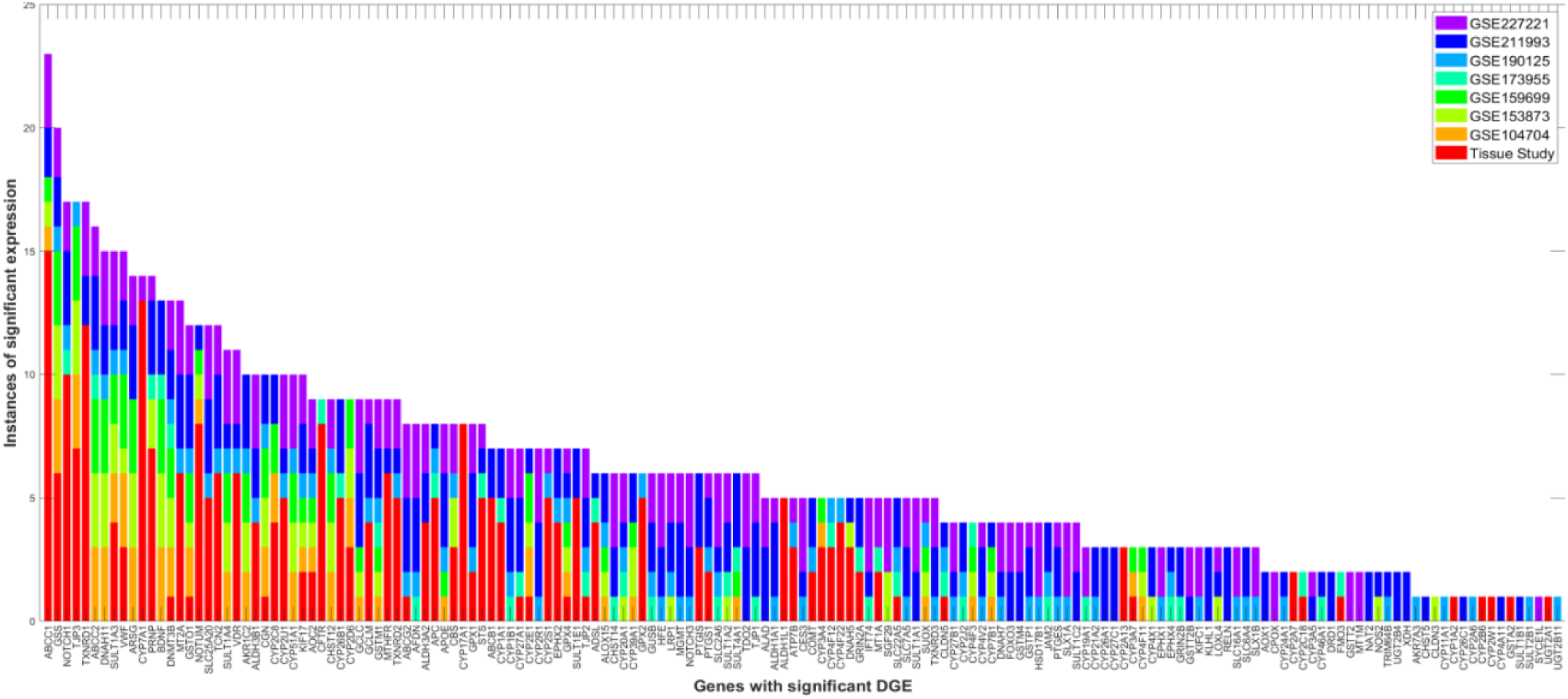
Genes with significant differential expression (both increased and decreased) in AD vs. control in combined studies. displays combined GEO and tissue study data displaying all genes with either increased or decreased differential expression. Note similar patterns of GOI differential expression in all datasets evaluated in both wings of this study.

**Figure 4.**
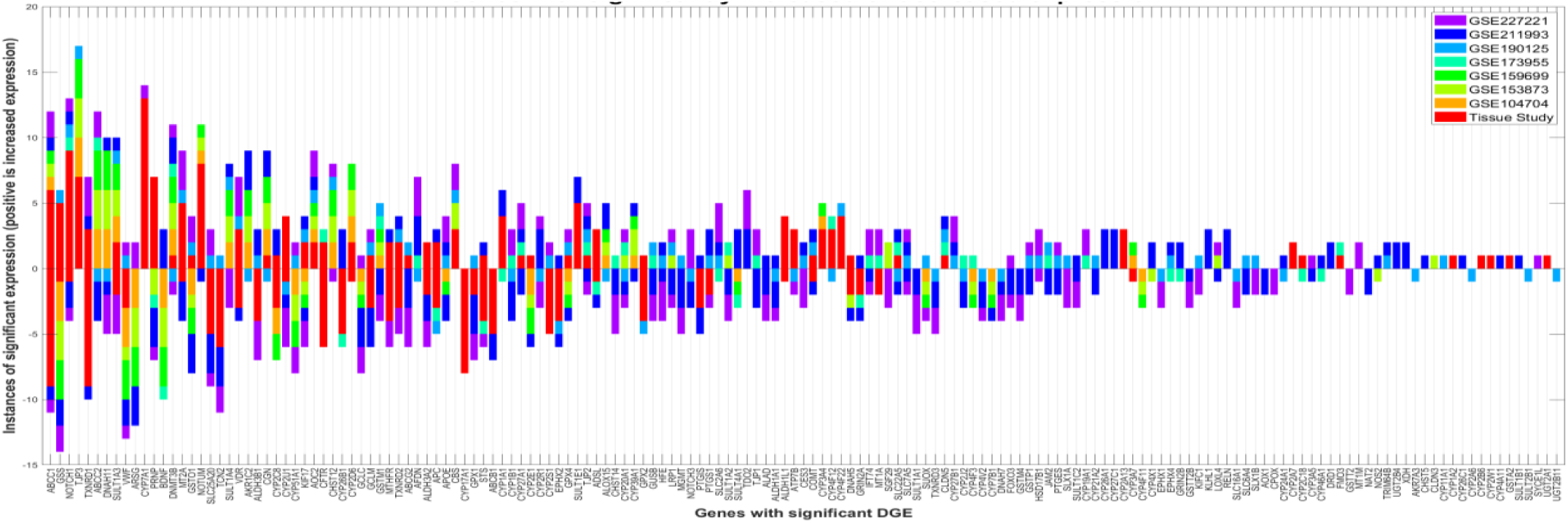
Genes with significantly increased or decreased differential expression in AD versus control in combined studies. displays combined GEO and tissue study data displaying all genes with either increased (bars above zero) and decreased differential expression (bars below zero), comparing AD to control. This data is the same data as in figure 3 but separating the increased and decreased expression genes.

**Figure 5.**
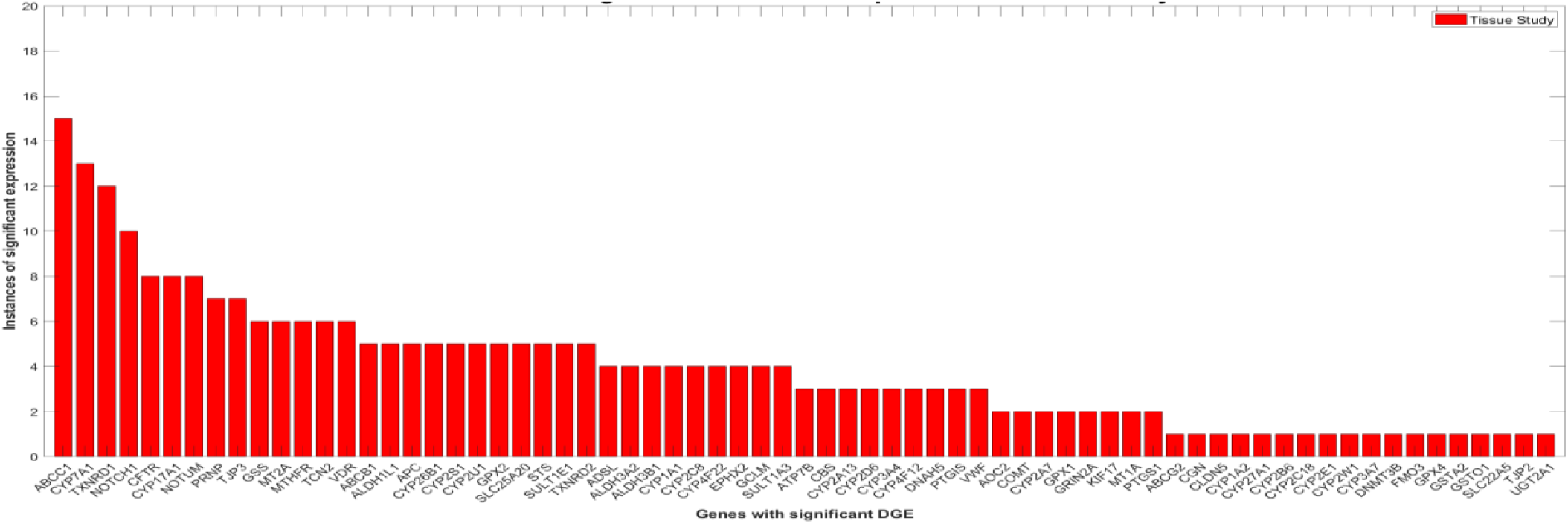
Genes with significant differential expression in phase 2. displays tissue study genes showing significant differential expression, comparing AD to control.

**Figure 6.**
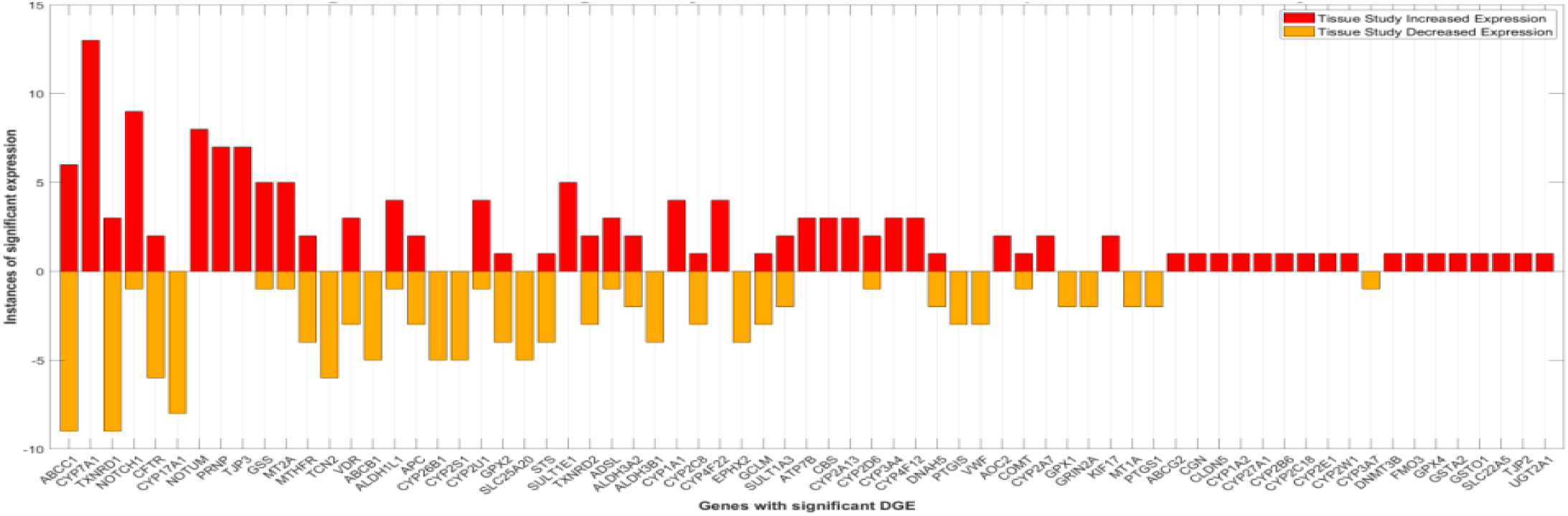
Genes with significantly increased or decreased differential expression in AD vs. control in phase 2. displays tissue study genes with significantly increased differential expression (bars above zero) and decreased differential expression (bars below zero), comparing AD to control. This data is the same data as in figure 5 but separating the increased and decreased expression genes.

**Figure 7.**
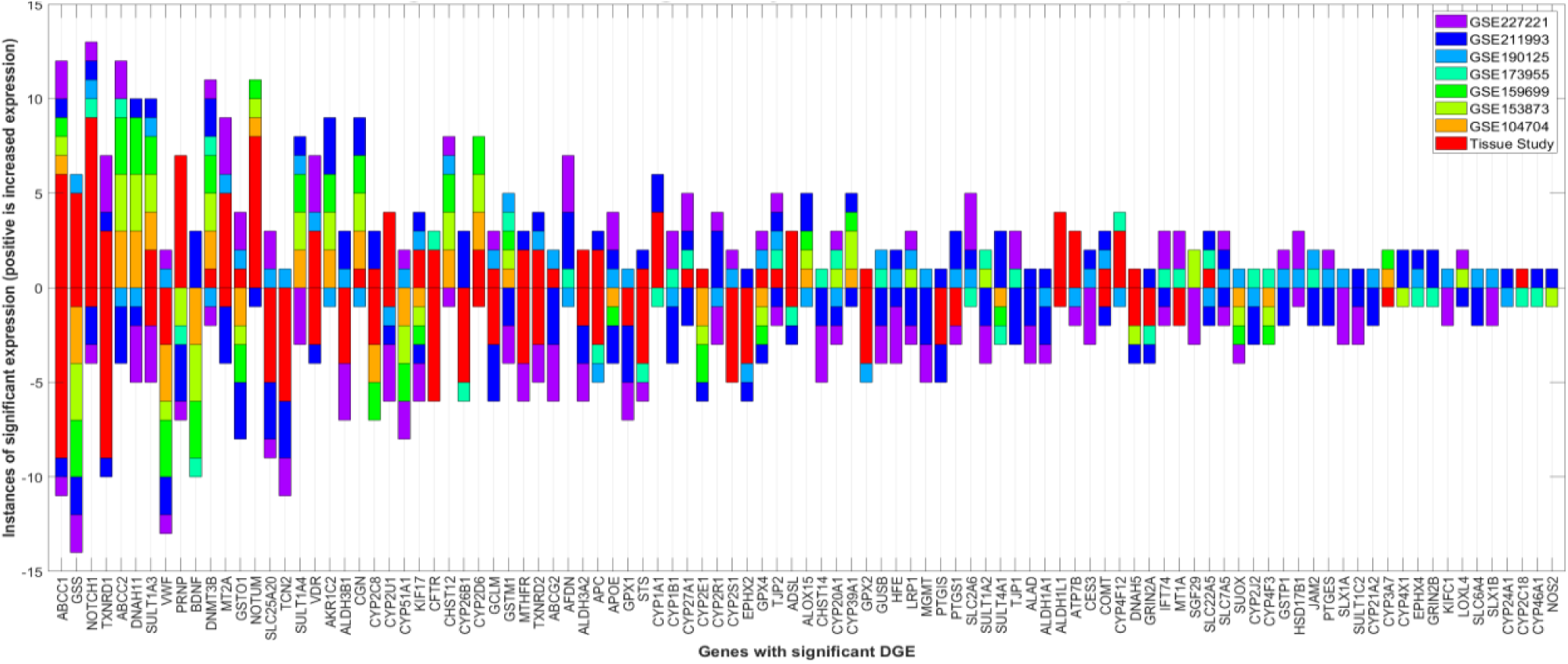
Genes with significantly increased and decreased differential expression in AD vs. control, in the same gene in combined studies. displays data from combined GEO and tissue study analyses showing all genes that have both increased and decreased significant differential expression in the same gene in different datasets. Bars above zero indicate increased differential expression. Bars below zero indicate decreased differential expression.

In aggregate, the bar charts provide graphic visualization of expression characteristics of our studies’ genes of interest, and by extension, detoxification genes more generally. Decreased expression of detoxification genes, while expected, appears to be accompanied by increased expression of other detoxification genes, perhaps in a compensatory response. Often the same genes show increased expression in one dataset and decreased expression in another dataset.

#### GEO and tissue dataset DGE comparisons

In total, 162 genes were found to be differentially expressed in the GEO datasets (Phase 1) evaluated plus the tissue study (Phase 2) analysis. 154 genes were differentially expressed in the GEO datasets; 69 genes were differentially expressed in the tissue study. 61 genes were differentially expressed in both arms of this study. See Table 1.

**Table 1.** Comparison of Differential Detoxification Gene Expression (DGE) in GEO and Tissue Studies.

| Category | Number of Genes | Description |
| --- | --- | --- |
| Total Unique Genes | 162 | Genes differentially expressed in either tissue or GEO study |
| GEO Study | 154 | Genes differentially expressed in GEO study |
| Tissue Study | 69 | Genes differentially expressed in tissue study |
| Overlap | 61 | Genes differentially expressed in both studies |

Another way to view the significance of the differentially expressed genes in this study is to assess the numbers of times a gene was found to be significant in the reported datasets (instances.). See Table 2.

**Table 2:** Top 50 Significant Gene Expression Instances.

| Gene Name | Instances |
| --- | --- |
| ABCC1 | 21 |
| GSS | 20 |
| TJP3 | 17 |
| TXNRD1 | 17 |
| NOTCH1 | 16 |
| ABCC2 | 15 |
| CYP7A1 | 15 |
| DNAH11 | 15 |
| SULT1A3 | 15 |
| VWF | 14 |
| ARSG | 13 |
| NOTUM | 13 |
| BDNF | 12 |
| DNMT3B | 12 |
| MT2A | 12 |
| PRNP | 12 |
| GSTO1 | 11 |
| SLC25A20 | 11 |
| SULT1A4 | 11 |
| TCN2 | 11 |
| VDR | 11 |
| ALDH3B1 | 10 |
| CYP2U1 | 10 |
| KIF17 | 10 |
| TXNRD2 | 10 |
| AKR1C2 | 9 |
| AOC2 | 9 |
| CFTR | 9 |
| CGN | 9 |
| CHST12 | 9 |
| CYP2C8 | 9 |
| CYP2D6 | 9 |
| GSTM1 | 9 |
| MTHFR | 9 |
| APC | 8 |
| CBS | 8 |
| CYP17A1 | 8 |
| GCLC | 8 |
| GCLM | 8 |
| STS | 8 |
| ABCG2 | 7 |
| AFDN | 7 |
| ALDH3A2 | 7 |
| APOE | 7 |
| CYP26B1 | 7 |
| CYP2S1 | 7 |
| CYP51A1 | 7 |
| EPHX2 | 7 |
| GPX1 | 7 |
| GPX4 | 7 |

Looking at the adjusted P value and Q values of the differentially expressed genes, two comparisons were created, one for each wing of the study. These comparisons are ranked in increasing order. Hence, the tops of the lists have the most significant values. Increased and decreased expression are not segregated. In another grouping the two wings of the study are co-mingled to focus on what may be the most significant genes in this study. See Table 3.

**Table 3:** Top 10 Adjusted P/Q values in GEO and tissue studies.

| Study Type | Gene | Adjusted P Value / Q Value | Expression Change |
| --- | --- | --- | --- |
| GEO Study (Adjusted P Value) |  |  |  |
|  | MT2A | 0 | Up |
|  | SLC2A6 | 4.80E-278 | Up |
|  | SLC2A6 | 1.58E-249 | Up |
|  | MT2A | 3.93E-248 | Up |
|  | CYP1B1 | 1.09E-177 | Up |
|  | VDR | 1.14E-140 | Up |
|  | PTGIS | 1.99E-130 | Down |
|  | CYP26A1 | 1.62E-125 | Up |
|  | APOE | 2.05E-121 | Down |
|  | CYP4X1 | 2.41E-120 | Up |
| Tissue Study (Q Value) |  |  |  |
|  | ABCC1 | 1.17E-14 | Down |
|  | MT2A | 5.17E-14 | Up |
|  | TXNRD1 | 1.20E-13 | Down |
|  | STS | 1.30E-13 | Down |
|  | MT2A | 2.19E-13 | Up |
|  | CYP17A1 | 3.89E-13 | Down |
|  | ABCB1 | 4.95E-13 | Down |
|  | APC | 9.05E-13 | Down |
|  | SLC25A20 | 4.26E-12 | Down |
|  | EPHX2 | 4.38E-12 | Down |

Adjusted P values: control family-wise error rate, more conservative, fewer false positives Q values: control false discovery rate, fewer false negatives

### Key Overlapping Genes (found in both studies)

1. ABC Transporters: ABCB1, ABCC1, ABCG2
2. Aldehyde Dehydrogenases: ALDH3A2, ALDH3B1
3. Cytochrome P450 Family: CYP1A1, CYP26B1, CYP27A1, CYP2A7, CYP2C18, CYP2C8, CYP2D6, CYP2E1, CYP2S1, CYP2U1, CYP3A4, CYP3A7, CYP4F12, CYP4F22, CYP7A1
4. Glutathione-related: GPX1, GPX2, GPX4, GSS, GSTO1
5. Metallothioneins: MT1A, MT2A
6. Sulfotransferases: SULT1A3, SULT1E1
7. Other Notable Genes: NOTCH1, VDR, VWF, MTHFR, TXNRD1, TXNRD2

For comprehensive table, see Supplementary material, Table S23.

## DISCUSSION

This paper articulates evidence supporting an association of AD and altered capacity for detoxification. The differential expression found in many detoxification genes in this study is supportive of this hypothesis. The GEO database portion of the present paper identified several genes from the GOI list with significant DGE. The liver tissue RNA-seq portion of this paper also identified several GOI genes with significant differential expression. Many of these genes were found multiple times in datasets examined. Support for the hypothesis is enhanced using two different platforms in these studies.

In the first three GEO datasets analyzed, a consistent finding was the higher number of genes showing significant DGE in AD versus young, as opposed to the AD versus old comparisons. This finding suggests an age-related general change in detoxification gene expression. The fewer genes with significant DGE in AD vs. old analyses would tend to implicate those specific genes in AD pathogenesis. Subsequent research into this age-related DGE association may be useful in confirming or refuting this association.

### DGE in 2023 GEO datasets compared to older GEO datasets

In the GEO portion of the present paper, the databases from 2023 compared to the older databases show considerably more significant DGE. In the older studies, GSE104704, GSE159699, GSE173955, and GSE153873, there were 25, 25, 38, and 27 genes showing significant DGE, respectively, in the most numerous age ranges, if noted. By contrast, in the 2023 studies, GSE211993-A79V/L150P, GSE227221, and GSE190125, significant DGE was found in 76/84/65, 47/75/62, and 81 genes respectively.

The 2023 datasets also demonstrated more extreme adjusted p-values:

Most significant adjusted p-value from older datasets: 4.3E-07 (GSE104704 AD V CTL, GSE159699 AD V CTL)

Most significant adjusted p-value from 2023 datasets: 5E-281 and zero, both from GSE227221 4H V CTL/24H V CTL respectively (the minimum positive number in Microsoft Excel is 2.2250738585072E-308, below which is zero.)

The increased DGE and adjusted p-value significance in the 2023 datasets is likely related to the design of the studies, utilizing affected and control groups that have little to no possibility of overlap or ambiguity.

The numbers of genes found to be significantly differentially expressed, and the finding of such DGE using 2 separate DGE experimental designs, adds significant weight to the possibility of changes in detoxification gene expression being a part of the pathophysiology of AD. Analyses of the specific functions of the identified genes in the DGE data is available in gene ontology databases as well as at www.genecards.org. How specific detoxification genes relate to AD pathophysiology is beyond the scope of this discussion but may be a useful topic for subsequent research to answer the question, "what is not being detoxified?" The decreased expression of the many detoxification genes identified in this data is consistent with toxicity related to toxins not optimally removed from the body being causative of the neurodegeneration of AD. The increased expression of other detoxification genes appears to be robust and may represent the body’s compensatory response to increased levels of toxins needing detoxification. The markedly increased expression of some detoxification genes may be an indication of the importance to the body of limiting the negative consequences of inadequate elimination of various toxins. The metabolic cost of secondarily increased detoxification gene expression needs additional scrutiny. Subsequent research into function and regulation of detoxification genes may be useful.

An interesting finding, from GSE227221, is the DGE noted with incubation of microglial derived cells with AB. After 4, 24, and 96 hours of incubation, compared to zero hours, the number of differentially expressed genes increased, peaking at 24 hours. After 96 hours of incubation, the number of differentially expressed genes declined somewhat. This suggests a nuanced regulatory role of AB in gene expression, potentially explaining some of the contradictory findings of studies of anti-AB medications in AD. How or if this finding relates to the emerging science of gene or neural networks in the regulation of gene expression is an intriguing question.

Gene expression is increasingly thought to be mediated in part through long noncoding RNA (lncRNA) [25]. The significant DGE we found related to both novel and known lncRNA in the tissue study is supportive of this concept.

Comparing the GEO dataset analyses and the tissue RNA-seq analyses, a few observations are pertinent. There are sixty-one genes showing significant GOI DGE in both phases of this study. Many genes show significant DGE but are not identical in the two phases of this study. This is not surprising given the different patient populations studied, and liver tissue being used for the tissue study, and brain, or other non-hepatic tissues used in the GEO dataset analyses. It is also worth considering that regional or ethnic differences could account for the differing DGE results in the two phases of this study.

One weakness of this investigation is the limited number of datasets utilized in the GEO portion of our analysis. The screening for GEO datasets appropriate for analysis led to only eight datasets that met the preset criteria for this study, two of which reported duplicate data. One may speculate that a larger number of useful datasets with an increased number of studies would have modified the findings of detoxification gene DGE, however there were a limited number of GEO datasets meeting the keyword criteria and passing further inspection. Subsequent investigators may want to consider evaluating differential detoxification gene expression in several additional databases, in larger numbers of subjects, possibly using different analytic platforms. The number of differentially expressed GOI genes required to support the hypothesis put forth is not clear, however, given the AD versus control format, even a small number of differentially expressed genes is likely highly pertinent.

Strengths of the present study include a plausible hypothesis, use of two separate platforms and experimental designs in analyzing the GEO versus the tissue datasets, and the use of control groups in each wing of this study, with impressive, adjusted p-values and Q values. The possibility of chance creating an illusion of significant association on which the present hypothesis rests is very low.

Insights into pathophysiology of AD derived through volumes of research into proposed mechanisms, while valuable, have not progressed to understanding the ultimate cause of AD. Already accepted mechanisms of AD include the abnormal proteins, AB and tau, and neuroinflammation. With the advent of therapeutic measures that remove AB from the brains of early AD or mild cognitive impairment patients, with progress in understanding of the risks of these measures, and now with a degree of benefit (albeit modest) being documented, neuroscientists are beginning to make progress in treating AD *after* the onset of neurodegeneration [26].

This manuscript addresses different questions: What is the cause of neuroinflammation, leading to excessive AB and tau protein? Is prevention possible?

The present paper presents data consistent with a new and plausible hypothesis, distilled from the above:

*Numerous genetically determined pathways function to detoxify a variety of xenobiotics and endogenously produced compounds. The effectiveness of these pathways, and hence vulnerability to toxic exposure differs in everyone. Unremoved toxins from liver, brain, and elsewhere in the body create toxic exposure leading to further oxidative (also known as free radical) stress in neural and other tissues, which is individually variable, causing neural (and other) inflammation. Prior reports have established that AD brains are consistently found to be inflamed*[27]*. AB and tau proteins accumulate in the brain as a response to injury, much as wound crusting and eschar occur from injury in other parts of the body. This response to injury has both beneficial and injurious properties. AB may influence detoxification gene expression via a nuanced feedback mechanism*.

## CONCLUSION

The above research findings and discussion present facts and logic that may lead to consideration that impairment of detoxification pathways contributes to the pathophysiology of AD. Perhaps this this paper will lead other investigators to further investigate DGE in detoxification genes in Alzheimer’s disease specifically and multiple other diseases of aging more generally.

## MATERIALS AND METHODS

### Study design

*Research objective:* Using gene set analysis I report an evaluation of differential gene expression (DGE) in detoxification genes in AD subjects versus control subjects, or in other pertinent groupings. The chance identification of a potential detoxification deficit in the AD associated APOE system led to the above hypothesis. I identified a specific gene set, called genes of interest (GOI), known to be active in detoxification processes. I embarked on a gene set expression analysis investigation. Two phases were planned. In Phase 1, I planned to analyze RNA sequencing (RNA- seq) data from Gene Expression Omnibus (GEO) datasets, using every GEO dataset that met preselected keyword and subjective screening criteria. The samples assayed were either neurologic tissue, undifferentiated stem cells, or whole blood. Using the associated GEO2R analytic platform and the GOI gene set, GEO datasets were processed several times, with identical results in each repeat analysis. In Phase 2, I compared gene expression in AD versus control cohorts in commercially sourced bio- banked tissue, again using gene set expression analysis with the same GOI list, in a commercial RNA-seq platform provided by LC Sciences in Houston, Texas. I planned to use liver tissue for RNA-seq as detoxification gene activity is known to be most prominent in the liver[28]. This led to sourcing challenges. However, hepatic samples for both AD (seven samples) and control (five samples) tissues were obtained after contacting several commercial biobanks. Given that banked liver tissue is not a common sample type for evaluation of neurodegenerative disease, the twelve samples obtained were all that were available from the biobanks I contacted.

Both phases of this study were observational. Randomization and blinding were not used in either phase of this study.

Hardy-Weinberg equilibrium was considered. Adjusted P-values and Q values reached significance (less than 0.05) in many genes. Hence, criteria were not met for Hardy-Weinberg equilibrium (null hypothesis.) HWE requires random mating, no mutation, no gene flow, no natural selection, and large population size.

Population stratification is noted, based on the Phase 1 populations being western European or American, and the Phase 2 samples coming from Russian subjects. Aside from speculation about the consequences of this stratification, no other means of controlling the results of this stratification was employed. No other sources of bias were identified. I could find no previous genomic study evaluating detoxification gene expression in AD or any other disease. Multiple comparisons were addressed in adjusted P-value and Q value computations. No genotyping was attempted aside from that reported in the metadata and RNA-seq results in this manuscript. Outcome data was not collected or reported. STREGA guidelines have been followed in the performance of the research and its reporting in this manuscript where relevant. A STREGA summary table from a 2009 publication[29] is included in Supplementary material.

In the Phase 1 (GEO) study the datasets studied were grouped into diseased versus control, normal stem cells versus mutation-containing stem cells, or microglia with no AB incubation versus microglia with 4, 24, or 96 hours of AB incubation. In the phase 2 study, we performed RNA-seq on RNA extracted from AD samples and non-AD samples, grouped in a variety of ways to facilitate useful comparisons.

*AI note:* For creation of tables 1-3 in this manuscript, artificial intelligence assistance was used - Gemini Advanced 1.5 Pro - using the prompt "create a table using the following dataset." The tables created were carefully reviewed for accuracy. AI was not used for composition of text in this manuscript.

### Phase 1

In Phase 1 of the present report, Gene Expression Omnibus (GEO) datasets were processed through the associated GEO2R platform. A resource provided by the NCBI (National Center for Biotechnology Information), GEO is a publicly funded, free, and authoritative repository for archiving and distributing functional genomic data. Datasets are publicly available from the GEO database[30]. The GEO2R tool is useful in evaluating gene expression values. In GEO2R, DESeq2, Next Generation Sequencing technologies, high-throughput sequencing, and bulk-cell transcriptomics analysis are utilized for DGE analysis, yielding expression profiling data. The data entry, analytic process, and DGE analysis are straightforward and easily understood. The performance of GEO2R is consistent and reproducible. There are several published reports on the use of GEO2R for gene set analysis[31, 32].

The following keywords were utilized to screen for appropriate GEO series (GSE) datasets:

-Alzheimer’s, RNA-seq, human, Gene Expression Omnibus, GEO2R

Exploring the present hypothesis on GEO datasets, the keyword search identified ninety potentially suitable datasets. The keyword-based search is objective and easily replicated. Further scrutiny of each of the ninety datasets and associated metadata was carried out, evaluating the peer reviewed manuscript associated with the specific GEO file, metadata, composition of target and control population, and other aspects of the datasets. By its nature, this part of a GEO dataset evaluation is subjective. An example of a subjectively included dataset is GSE211993, a transcriptome analysis which utilized two forms of the Presenilin 1 gene, A79V and L150P, comparing each to CRISPR/Cas9 gene corrected samples. Although not specifically an AD versus control study, the methods used, and the insight derived therefrom justified including this study in the selected datasets. An example of an excluded dataset might be GSE118158 which compared brain tissue from various regions in brains from subjects with neuropsychiatric conditions but had no specific disease versus control grouping in the described metadata and was deemed not useful in our study. The use of the specific keywords cast a net that identified many more non-useful (like GSE118158) than pertinent datasets, requiring individual, *subjective* analyses of each dataset’s characteristics to select or reject those that may or may not be useful in investigating the hypothesis. I identified eight datasets with useful characteristics that optimally addressed the research questions pertinent to the hypothesis, in my opinion.

I used previously sequenced GEO series (GSE) samples, comparing DGE between human brain tissues from AD subjects and from non-AD subjects, or utilizing other experimental designs when datasets were structured in a way conducive to a modified design. Table 4 presents a summary of the selected datasets used for GEO2R DGE analyses.

**Table 4.**
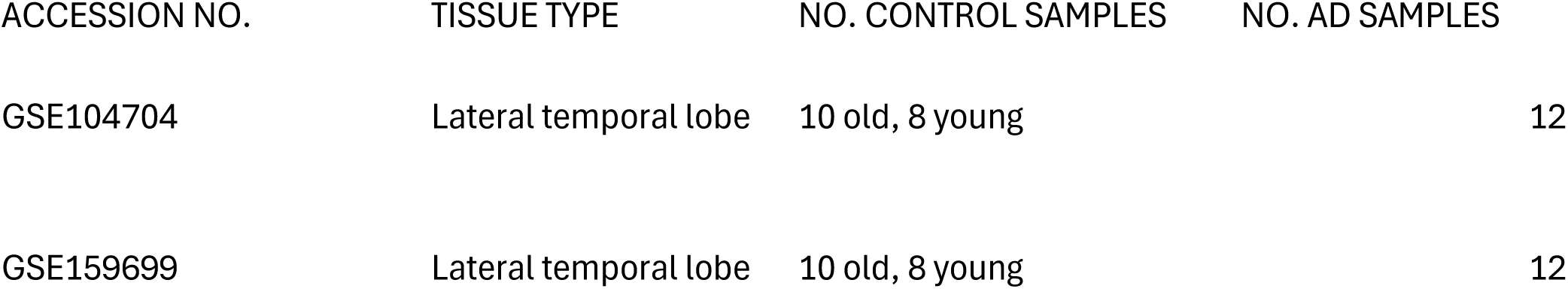
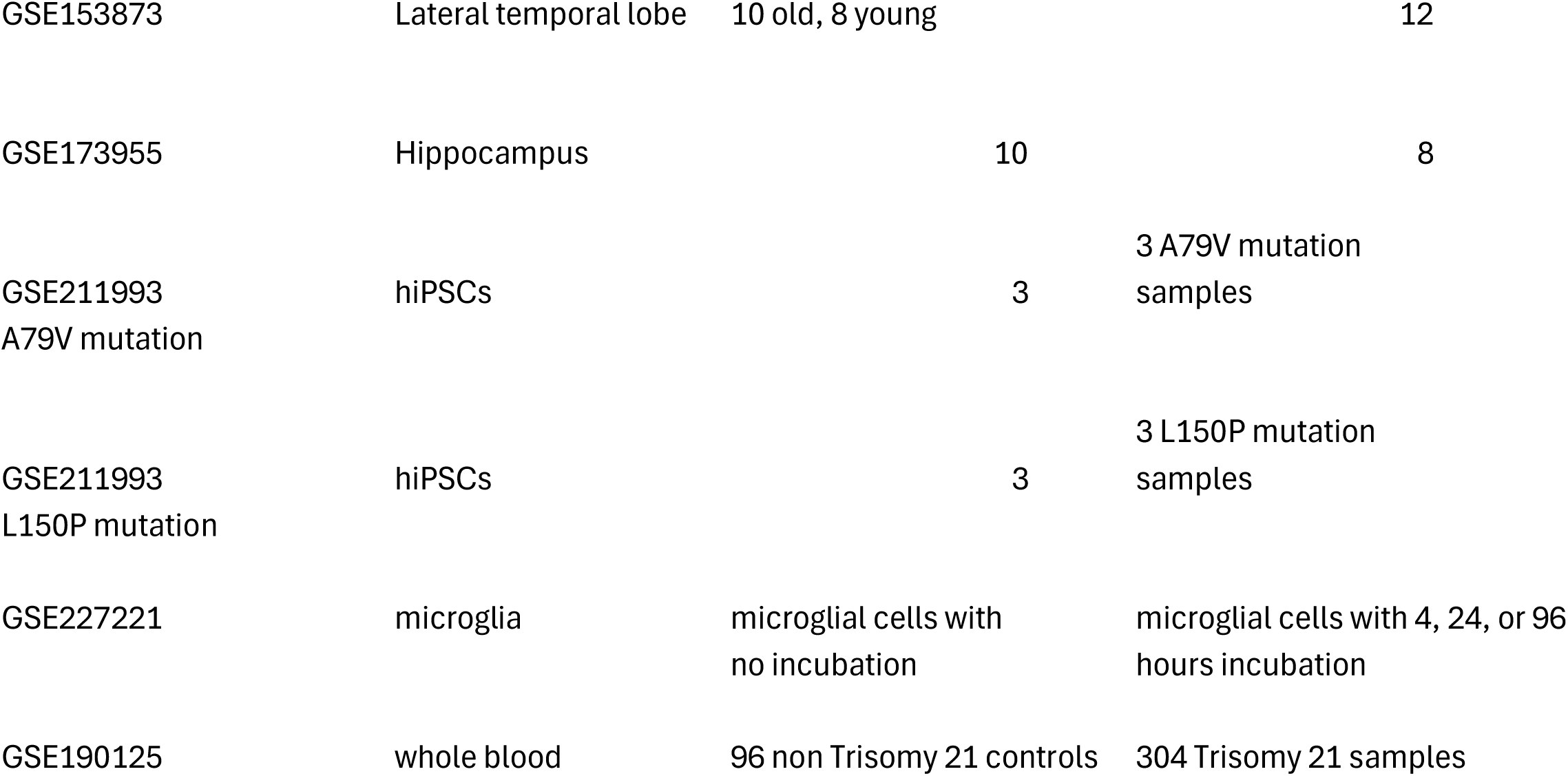

### Steps in GEO2R analysis

Follow the instructions in the NCBI website for GEO2R analysis.

Identify the GSE dataset of interest. For this example, I used GSE153873. Only some GSE datasets have the GEO2R option.

Click the "analyze with GEO2R" option.

A list of metadata appears.

Define the desired groups, usually AD as the first group, and control (CTL) the second one. If there are age identifiers, they are grouped separately as "young" or "old". No age delineation was seen in the AD groups.

Highlight the AD group first, and the desired CTL group separately, assigning each to the proper group designation. Errors in this step are common mistakes that will produce inaccurate DGE results. Only analyze two groups at a time, even if there are more groupings than two.

Click the "Analyze" option at the bottom of the page.

This takes a few seconds and returns the top 250 differentially expressed genes in a "Numbers" spreadsheet. This could be a stopping point for researchers interested only in looking at such data. As this project is related to understanding specific gene differential expression, we need to proceed further.

At the top of the table, click "download full table". This step takes about 30 seconds. The results are downloaded into the "downloads" folder. Open that file with a double click. Some browsers open this file automatically. A very large file labeled "GSE153873.top.table" (or whatever file is being analyzed) opens. Further analysis is done by your analytic platform of choice. We used Matlab or Excel for our analyses.

### Phase 2

In analyzing the present hypothesis, and in view of the striking DGE results from the GEO database portion, an investigation was organized utilizing hepatic tissue samples, from which RNA was extracted. One goal in performing the tissue study was to diversify the platforms used for data analysis to minimize the chance of spurious similar results. Another goal was to confirm the findings from the GEO dataset analyses. Additionally, since existing scientific reports have documented much detoxification activity in hepatic tissues, we planned to utilize *liver* tissue from AD and control samples in seeking DGE data. The research was conducted in accordance with prevailing legal requirements. Despite the challenges in procuring liver tissue from neurologically afflicted patients, through a commercial source we obtained bio-banked liver tissue from seven AD patients, and five controls, all deceased. The analysis was carried out with comparisons of AD and non-AD tissue, and male/female and combined samples. Annotation was:

ADFVADM: AD females versus AD males

ADFVCTL: AD females versus control (gender not specified)

ADFVCTLF: AD females versus control females

ADFVCTLM: AD females versus control males

ADMVADF: AD males versus AD females

ADMVCTL: AD males versus control

ADMVCTLF: AD males versus control female

ADMVCTLM: AD males versus control males

ADVCTL: AD (gender not specified) versus control

These annotated categories are further subdivided into subcategories of gene differential expression, transcript differential expression, known long non-coding RNA (lncRNA) differential expression, and novel lncRNA differential expression.

For details of specifics of RNA-seq in the tissue study,

see Supplementary Material.

### Genes if interest (GOI)

Throughout this paper, a list of 203 genes involved in detoxification processes was utilized in seeking DGE. The GOI are the sole genes for which DGE was sought and measured. The process used and criteria used for selection of the GOI genes is discussed in Supplementary Material. The genes of interest (GOI) are listed in Supplementary Material Table 1. Table 2 in Supplementary Material is a graphical illustration of the GOI list with the associated literature sources supporting the inclusion of each GOI in the list.

## Data Availability

All data produced in the present study are available upon reasonable request to the author.

## Acknowledgments

For their insights and advice, I thank Donald Moss, and Zhiyi Liu. For organization and creation of figures, I thank Evan McCaulley. This research was conducted utilizing resources from the Gene Expression Omnibus and the associated platform, GEO2R. Tissue sequencing was done by Dr. Zhiyi Liu and associates at LC Sciences, LLC of Houston, Texas.

Tissue acquisition was through AMSBIO, Cambridge, Massachusetts.

The AI tool, Gemini Advanced 1.5 Pro, was used to assist in creating tables 1-3 in the main text.

