## Supplementary Material for "Impaired Pathways of Detoxification and Risk of Alzheimer’s Disease in Humans - Differential Gene Expression Studies via Gene Set Expression Analysis"

#### Specifics of RNA-seq in phase 2 - tissue study

RNA sequencing (RNA-seq) was carried out according to the description below. We compared liver tissue from the AD and control samples and calculated DGE data.

Detailed description of RNA sequencing of banked tissue, performed by LC Sciences, Houston, Texas, follows (text provided by LC Sciences).

For Total RNA sequencing, RNA integrity was checked with Agilent Technologies 2100 Bioanalyzer. Ribosomal RNA was removed, followed by fragmentation with divalent cation buffers in elevated temperature. The sequencing library was prepared following Illumina's TruSeq-stranded-total-RNA-sample preparation protocol. Quality control analysis and quantification of the sequencing library were performed using Agilent Technologies 2100 Bioanalyzer High Sensitivity DNA Chip. Paired-ended sequencing was performed on Illumina's NovaSeq 6000 sequencing system. For transcript assembly, initially, Cutadapt and perl scripts in house were used to remove the reads that contained adaptor contamination, low quality bases and undetermined bases. Then sequence quality was verified using FastQC. HISAT2 was used to map reads to the genome of homo sapiens (Version: v107). The mapped reads of each sample were assembled using StringTie. Then, all transcriptomes from our 12 samples were merged to reconstruct a comprehensive transcriptome using perl scripts and gffcompare. After the final transcriptome was generated, StringTie and ballgown were used to estimate the expression levels of all transcripts.

**lncRNA (long non-coding RNA) identification:**

First of all, transcripts that overlapped with known mRNAs, known lncRNAs and transcripts shorter than 200 bp were discarded. Then we utilized CPC[ and CNCI to predict transcripts with coding potential. All transcripts with CPC score < 0.5 and CNCI score < 0 were retained and considered as novel lncRNAs. The remaining transcripts with class code (i, j, o, u, x) were considered as lncRNAs.

**Differential expression analysis of transcripts:**

StringTie [3] was used to perform expression levels for mRNAs and lncRNAs by calculating FPKM. mRNAs/lncRNAs differential expression analysis was performed by DESeq2[6] software between two different groups (and by edgeR[7] between two samples). The genes/mRNAs/lncRNAs with the parameter of false discovery rate (FDR) below 0.05 and absolute fold change ≥ 2 were considered differentially expressed genes/mRNAs/lncRNAs.

**Alternative Splicing Analysis:**

rMATS[8] (version 4.1.1) (http://rnaseq-mats.sourceforge.net) was used to identify alternative splicing events and analyze differential alternative splicing events between samples. We identified AS events with a false discovery rate (FDR) < 0.05 in a comparison as significant AS events.

**Gene Set Enrichment Analysis (GSEA):**

We performed gene set enrichment analysis using software GSEA[9] (v4.1.0) and MSigDB to identify whether in a set of genes in specific GO terms, KEGG pathways show significant differences in two groups. Briefly, we input gene expression matrix and rank genes by Signal2Noise normalization method. Enrichment scores and p value was calculated in default parameters. GO terms, KEGG pathways meeting this condition with |NES|>1, NOM p-val<0.05, FDR q-val<0.25 were considered to be different in two groups.

**Target gene prediction and functional analysis of lncRNAs:**

To explore the function of lncRNAs, we predicted the cis-target genes of lncRNAs. lncRNAs may play a cis role acting on neighboring target genes. In this study, coding genes in 100,000 upstream and downstream were selected by perl script. Then, we showed functional analysis of the target genes for lncRNAs by using the scripts in house. Significance was expressed as a p value < 0.05.

**Localization of lncRNAs on genome:**

Two diagrams were generated to show the localization and abundance of lncRNAs in genome using the program Circos[10]. One diagram showed lncRNA expression level in different samples. The lncRNAs in the other diagram were subdivided into five categories according to their class code generated by StringTie: (i) a transfrag falling entirely within a reference intron (intronic); (j) potentially novel isoform or fragment at least one splice junction is shared with a reference transcript; (o) generic exonic overlap with a reference transcript; (u) unknown, intergenic transcript (intergenic); (x) Exonic overlap with reference on the opposite strand (antisense).

**Genes of interest - discussion**

A group of detoxification genes, called "genes of interest" (GOI), was identified to study differential gene expression (DGE) for this study. The GOI list was created using several methods. Firstly, known detoxification gene families were utilized. These families include:

-The cytochrome P450 (CYP) family[33] yielding the most GOI genes for this study, fifty-eight. The CYP family is important in the metabolism, biotransformation, and elimination of xenobiotics (drugs, pesticides, heavy metals, and other substances not naturally occurring in an individual), hormones, neurotransmitters and other endogenous substances.

-The UTP-Glycotransferase (UGT) family genes, which are involved in metabolism and elimination of many xenobiotics as well as endogenous compounds [34]. They also serve signaling functions involving regulation of various aspects of cellular physiology. Twenty-one UGT genes are in the GOI list assembled for the current manuscript.

-The glutathione family, which is a wide-spread compound in humans accounting for considerable metabolizing and detoxifying activity, cancer risk, and susceptibility to oxidative and free radical reactions [35]. Fifteen glutathione related genes are included in the GOI list in this paper.

-The cytosolic sulfotransferase gene family (SULT), by means of sulfation, is involved in the metabolism and elimination of various xenobiotics and endogenous compounds including catecholamines, steroid and thyroid hormones [36].

Fourteen SULT genes are included in the GOI list in this manuscript.

-Solute carrier genes (SLC), which code for proteins that involve membrane transport. These genes/proteins regulate cell volume and remove waste and other un-needed substances [37]. There are seven SLC genes in the GOI list.

-EPHX genes code for enzymes important in metabolism and elimination of xenobiotics and endogenous substances. Deficiencies in EPHX activities are associated with a variety of diseases [38]. Four EPHX genes are in the GOI list associated with this paper.

-ABC genes (ATP synthase-binding cassette transporters) are involved in energy-requiring movement of substances across membranes either for elimination or uptake of those substances. These substances are both xenobiotics as well as endogenous compounds [39]. Four ABC genes are included in the GOI list for the present paper.

-ALDH genes are involved in the metabolism and excretion of aldehydes, both exogenous and endogenous. These substances include pharmaceutical and endogenously produced agents [40]. Three ALDH genes are included in the GOI list for this manuscript.

-The CHST (carbohydrate sulfotransferase) gene family is involved in sulfation of cellular components in specific configurations. Under or over sulfation is associated with toxicity and various disease states [41]. Three CHST genes are reported in the GOI list in this paper.

-DNAH genes are involved in structure and function of dynein structures which impact function of cilia and flagella, being involved in multiple conditions as a result [42, 43]. Three DNAH genes are included in the GOI list in the present paper.

-TJP genes are related to cell-cell junction integrity, potentially allowing inflammation-causing substances entry into physiologic spaces [44]. Three TJP genes are included in the GOI list in the present paper.

-TXNRD genes are related to oxidative stress by bocking reactive oxygen species [45]. Three TXNRD genes are reported in the GOI list in the present paper.

The SLX gene family is involved in repair of DNA damage and genome stability[46]. Two SLX genes are in the GOI list.

The following eleven genes are included as single genes, not as members of a gene family.

HSD17B1 is a gene involved in estrogen metabolism [47].

TCN2 is involved in production and metabolism of transcobalamin [48].

HFE is involved in iron uptake and metabolism [49].

KLHL1 is involved in risk of asthma from early life tobacco exposure [50].

MGMT is a gene modulating the activity of chemo-therapeutic agents [51].

The PRNP gene is related to copper and iron regulation, in association with breast cancer risk [52].

PTGES codes for a gene involved in prostaglandin synthesis and regulation of inflammation [53].

PTGS1 is involved in cyclooxygenase activity and associated with cardiovascular and other risks [54].

RELN variants are associated with a high risk of dementia, and with impaired excretion of specific mutant proteins [55].

SYCE1L has been associated with male infertility [56].

VDR relates to the vitamin D receptor [57].

Additionally, other peer reviewed studies identifying detoxification gene groups were noted. These studies include:

-Santos, et at. who, in a 2022 publication[58], reported genes impacting permeability of neurologic structures, such as blood brain barrier, as well as placental genes, in addition to detoxification genes. The complete Santos gene list includes 519 genes. The detoxification portion of the Santos list includes 297 genes.

-Andreoli and Sprovieri, in a 2017 report [11], described a group of gene variants which they found to impact risk of mercury toxicity. Seventeen of these variants were included in the genes of interest list,

Woods, et al. in the Casa Pia Children's Amalgam Clinical Trial reported genetic risk of susceptibility to mercury in a 2014 report [9]. Eleven genes from this trial are included in the GOI list.

Supplementary Table S1 GOI list

ABCB1

ABCC1

ABCC2

ABCG2

ADSL

AFDN

AKR1C2

AKR7A3

ALAD

ALDH1A1

ALDH3A2

ALDH3B1

ALOX15

AOC2

AOX1

APC

APOE

ARSG

ATP7B

BDNF

CBS

CES3

CFTR

CGN

CHST12

CHST14

CHST5

CLDN3

CLDN5

COMT

CPOX

CYP11A1

CYP11B1

CYP11B2

CYP17A1

CYP19A1

CYP1A1

CYP1A2

CYP1B1

CYP20A1

CYP21A2

CYP24A1

CYP26A1

CYP26B1

CYP26C1

CYP27A1

CYP27B1

CYP27C1

CYP2A

CYP2A13

CYP2A6

CYP2A7

CYP2B6

CYP2C18

CYP2C19

CYP2C8

CYP2C9

CYP2D6

CYP2E1

CYP2F1

CYP2J2

CYP2R1

CYP2S1

CYP2U1

CYP2W1

CYP39A1

CYP3A4

CYP3A5

CYP3A7

CYP46A1

CYP4A11

CYP4A22

CYP4B1

CYP4F11

CYP4F12

CYP4F2

CYP4F22

CYP4F3

CYP4F8

CYP4V2

CYP4X1

CYP4Z1

CYP4Z2P

CYP51A1

CYP5A1

CYP7A1

CYP7B1

CYP8A1

CYP8B1

DNAH11

DNAH5

DNAH7

DNMT3B

DRD1

EPHX1

EPHX2

EPHX3

EPHX4

FMO3

FOXO3

GCLC

GCLM

GPX1

GPX2

GPX4

GRIN2A

GRIN2B

GSS

GSTA1

GSTA2

GSTM1

GSTM4

GSTO1

GSTP1

GSTT1

GSTT2

GSTT2B

GUSB

HFE

HSD17B1

IFT74

JAM2

KIF17

KIFC1

KLHL1

LOXL4

LRP1

MGMT

MT1A

MT1M

MT2A

MT4

MTHFR

NAT2

NOS2

NOTCH1

NOTCH3

NOTUM

PRNP

PTGES

PTGIS

PTGS1

RELN

SGF29

SLC16A1

SLC22A5

SLC22A8

SLC25A20

SLC2A6

SLC6A4

SLC7A5

SLX1A

SLX1B

STS

SULT1A1

SULT1A2

SULT1A3

SULT1A4

SULT1B1

SULT1C2

SULT1C3

SULT1C4

SULT1D1P

SULT1E1

SULT2A1

SULT2B1

SULT4A1

SULT6B1

SUOX

SYCE1L

TCN2

TDO2

TJP1

TJP2

TJP3

TRIM64B

TXNRD1

TXNRD2

TXNRD3

UGT1A1

UGT1A10

UGT1A3

UGT1A4

UGT1A5

UGT1A6

UGT1A7

UGT1A8

UGT1A9

UGT2A1

UGT2A2

UGT2A3

UGT2A3P7

UGT2B

UGT2B10

UGT2B11

UGT2B15

UGT2B17

UGT2B28

UGT2B4

UGT2B7

VDR

VWF

XDH

Supplementary Table S2: Genes of interest worksheet

| | Santos | PRL | CYP | EPHX | Casa Pia | Andr. Sprov. |
|---|---|---|---|---|---|---|
| ABCB1 | x | x | | | | x |
| ABCC1 | x | x | | | | x |
| ABCC2 | | x | | | | x |
| ABCG2 | x | x | | | | |
| ADSL | x | | | | | |
| AFDN | x | | | | | |
| AKR1C2 | x | | | | | |
| AKR7A3 | x | | | | | |
| ALAD | x | | | | | |

| | | | | | | |
|---|---|---|---|---|---|---|
| ALDH1A1 | x | x | | | | |
| ALDH3A2 | x | x | | | | |
| ALDH3B1 | x | x | | | | |
| ALOX15 | x | | | | | |
| AOC2 | x | | | | | |
| AOX1 | x | | | | | |
| APC | x | | | | | |
| APOE | | | | | x | |
| ARSG | x | | | | | |
| ATP7B | | | | | | x |
| BDNF | x | | | | x | x |
| CBS | x | | | | | |
| CES3 | x | | | | | |
| CFTR | x | | | | | |
| CGN | x | | | | | |
| CHST12 | | x | | | | |
| CHST14 | | x | | | | |
| CHST5 | | x | | | | |
| CLDN3 | x | | | | | |
| CLDN5 | x | | | | | |
| COMT | x | | | | x | |
| CPOX | | | | | x | |
| CYP11A1 | | | x | | | |
| CYP11B1 | | | x | | | |
| CYP11B2 | | | x | | | |
| CYP17A1 | | | x | | | |
| CYP19A1 | | | x | | | |
| CYP1A1 | | | x | | | |
| CYP1A2 | | | x | | | |
| CYP1B1 | | | x | | | |
| CYP20A1 | | | x | | | |
| CYP21A2 | | | x | | | |
| CYP24A1 | | | x | | | |
| CYP26A1 | | | x | | | |
| CYP26B1 | | | x | | | |
| CYP26C1 | | | x | | | |
| CYP27A1 | | | x | | | |
| CYP27B1 | | | x | | | |
| CYP27C1 | | | x | | | |
| CYP2A | | | x | | | |
| CYP2A13 | | | x | | | |
| CYP2A6 | | | x | | | |
| CYP2A7 | | | x | | | |

| | | | | |
|---|---|---|---|---|
| CYP2B6 | | | x | |
| CYP2C18 | | | x | |
| CYP2C19 | | | x | |
| CYP2C8 | | | x | |
| CYP2C9 | | | x | |
| CYP2D6 | | | x | |
| CYP2E1 | | | x | |
| CYP2F1 | | | x | |
| CYP2J2 | | | x | |
| CYP2R1 | | | x | |
| CYP2S1 | | | x | |
| CYP2U1 | | | x | |
| CYP2W1 | | | x | |
| CYP39A1 | | | x | |
| CYP3A4 | | | x | |
| CYP3A5 | | | x | |
| CYP3A7 | | | x | |
| CYP46A1 | | | x | |
| CYP4A11 | | | x | |
| CYP4A22 | | | x | |
| CYP4B1 | | | x | |
| CYP4F11 | | | x | |
| CYP4F12 | | | x | |
| CYP4F2 | | | x | |
| CYP4F22 | | | x | |
| CYP4F3 | | | x | |
| CYP4F8 | | | x | |
| CYP4V2 | | | x | |
| CYP4X1 | | | x | |
| CYP4Z1 | | | x | |
| CYP4Z2P | | | x | |
| CYP51A1 | | | x | |
| CYP5A1 | | | x | |
| CYP7A1 | | | x | |
| CYP7B1 | | | x | |
| CYP8A1 | | | x | |
| CYP8B1 | | | x | |
| DNAH11 | x | | | |
| DNAH5 | x | | | |
| DNAH7 | x | | | |
| DNMT3B | x | | | |
| DRD1 | x | | | |
| EPHX1 | x | | | x |

| | | | | | | |
|---|---|---|---|---|---|---|
| EPHX2 | x | | | x | | |
| EPHX3 | x | | | x | | |
| EPHX4 | x | | | x | | |
| FMO3 | x | | | | | |
| FOXO3 | | x | | | | |
| GCLC | | | | | | x |
| GCLM | | | | | | x |
| GPX1 | | | | | | x |
| GPX2 | | x | | | | |
| GPX4 | | | | | | x |
| GRIN2A | | | | | x | |
| GRIN2B | | | | | x | |
| GSS | | | | | | x |
| GSTA1 | x | | | | | x |
| GSTA2 | x | | | | | |
| GSTM1 | x | | | | | x |
| GSTM4 | x | | | | | |
| GSTO1 | x | | | | | |
| GSTP1 | x | | | | | |
| GSTT1 | x | | | | x | x |
| GSTT2 | x | | | | | |
| GSTT2B | x | | | | | |
| GUSB | x | | | | | |
| HFE | | x | | | | |
| HSD17B1 | | | | | | |
| IFT74 | x | | | | | |
| JAM2 | x | | | | | |
| KIF17 | x | | | | | |
| KIFC1 | x | | | | | |
| KLHL1 | | x | | | | |
| LOXL4 | x | | | | | |
| LRP1 | x | | | | | |
| MGMT | | x | | | | |
| MT1A | x | | | | | |
| MT1M | | | | | x | x |
| MT2A | | | | | x | |
| MT4 | | | | | | x |
| MTHFR | x | | | | | |
| NAT2 | x | | | | | |
| NOS2 | x | | | | | |
| NOTCH1 | x | | | | | |
| NOTCH3 | x | | | | | |
| NOTUM | x | | | | | |

| | | | | | | |
|---|---|---|---|---|---|---|
| PRNP | | x | | | | |
| PTGES | | x | | | | |
| PTGIS | x | | | | | |
| PTGS1 | | x | | | | |
| RELN | | x | | | | |
| SGF29 | x | | | | | |
| SLC16A1 | x | | | | | |
| SLC22A5 | x | | | | | |
| SLC22A8 | | | | | | x |
| SLC25A20 | | | | | | |
| SLC2A6 | | | | | | |
| SLC6A4 | x | | | | x | |
| SLC7A5 | x | | | | | x |
| SLX1A | | | | | | |
| SLX1B | | | | | | |
| STS | x | | | | | |
| SULT1A1 | x | | | | | |
| SULT1A2 | x | | | | | |
| SULT1A3 | x | | | | | |
| SULT1A4 | x | | | | | |
| SULT1B1 | x | | | | | |
| SULT1C2 | x | | | | | |
| SULT1C3 | x | | | | | |
| SULT1C4 | x | | | | | |
| SULT1D1P | | | | | | |
| SULT1E1 | x | | | | | |
| SULT2A1 | x | | | | | |
| SULT2B1 | x | | | | | |
| SULT4A1 | x | | | | | |
| SULT6B1 | | | | | | |
| SUOX | x | | | | | |
| SYCE1L | | x | | | | |
| TCN2 | | x | | | | |
| TDO2 | | | | | x | |
| TJP1 | x | | | | | |
| TJP2 | x | | | | | |
| TJP3 | x | | | | | |
| TRIM64B | x | | | | | |
| TXNRD1 | x | | | | | |
| TXNRD2 | x | | | | | |
| TXNRD3 | x | | | | | |
| UGT1A1 | x | | | | | |
| UGT1A10 | x | | | | | |

| | | |
|---|---|---|
| UGT1A3 | x | |
| UGT1A4 | x | |
| UGT1A5 | x | |
| UGT1A6 | x | |
| UGT1A7 | x | |
| UGT1A8 | x | |
| UGT1A9 | x | |
| UGT2A1 | x | |
| UGT2A2 | x | |
| UGT2A3 | x | |
| UGT2A3P7 | | |
| UGT2B | | |
| UGT2B10 | x | |
| UGT2B11 | x | |
| UGT2B15 | x | |
| UGT2B17 | x | |
| UGT2B28 | x | |
| UGT2B4 | x | |
| UGT2B7 | x | |
| VDR | | x |
| VWF | x | |
| XDH | x | |

Santos, et al. Gene Environment Interactions in ASD

PRL=peer reviewed literature

CYP= cytochrome P450 genes

EPHX=epoxide hydrolase genes

Casa Pia=Woods, et al. Children's Amalgam Clinical Trial

Andr. Sprov.=Andreoli and Sprovieri Genetic Aspects of Mercury Toxicity

Supplementary Table S3

| | | | | GSE104704 AD V YOUNG | | | | |
|---|---|---|---|---|---|---|---|---|
| GeneID | padj | pvalue | lfcSE | stat | log2FoldChange | baseMean | Symbol | Description |
| 2937 | 2E-06 | 7.5E-09 | 0.159 | -5.779914 | -0.91951627 | 291.51 | GSS | glutathione synthetase |
| 8701 | 3.1E-06 | 1.5E-08 | 0.23 | 5.66699 | 1.30239389 | 89.69 | DNAH11 | dynein axonemal heavy chain 11 |
| 9446 | 8.7E-06 | 6E-08 | 0.149 | -5.419326 | -0.80548328 | 368.6 | GSTO1 | glutathione S-transferase omega 1 |
| 1595 | 7.6E-05 | 1.4E-06 | 0.203 | -4.820235 | -0.97772232 | 827 | CYP51A1 | cytochrome P450 family 51 subfamily A member 1 |
| 1789 | 0.00098 | 5.1E-05 | 0.19 | 4.053104 | 0.76846258 | 42.58 | DNMT3B | DNA methyltransferase 3 beta |
| 445329 | 0.00145 | 8.6E-05 | 0.233 | 3.9281688 | 0.91528395 | 87.42 | SULT1A4 | sulfotransferase family 1A member 4 |
| 6818 | 0.00146 | 8.7E-05 | 0.232 | 3.9256083 | 0.90973415 | 87.67 | SULT1A3 | sulfotransferase family 1A member 3 |
| 246 | 0.00182 | 0.00012 | 0.413 | 3.851084 | 1.59213529 | 12.45 | ALOX15 | arachidonate 15-lipoxygenase |
| 6821 | 0.00377 | 0.00032 | 0.091 | -3.600549 | -0.32800787 | 1013.93 | SUOX | sulfite oxidase |
| 1244 | 0.00591 | 0.00059 | 0.289 | 3.4342476 | 0.99311625 | 28.7 | ABCC2 | ATP binding cassette subfamily C member 2 |
| 147111 | 0.00658 | 0.0007 | 0.451 | 3.390897 | 1.52934088 | 25.74 | NOTUM | notum, palmitoleoyl-protein carboxylesterase |
| 27134 | 0.00991 | 0.00123 | 0.279 | 3.2325358 | 0.90137962 | 56.4 | TJP3 | tight junction protein 3 |
| 9420 | 0.0106 | 0.00134 | 0.337 | -3.206351 | -1.07976728 | 645.57 | CYP7B1 | cytochrome P450 family 7 subfamily B member 1 |
| 2944 | 0.0163 | 0.0024 | 0.414 | 3.035192 | 1.25559038 | 156.54 | GSTM1 | glutathione S-transferase mu 1 |
| 314 | 0.0181 | 0.00282 | 0.22 | 2.9870456 | 0.65814209 | 75.54 | AOC2 | amine oxidase copper containing 2 |
| 55501 | 0.0181 | 0.00282 | 0.187 | 2.986628 | 0.55754202 | 870.89 | CHST12 | carbohydrate sulfotransferase 12 |
| 4363 | 0.0212 | 0.00351 | 0.106 | 2.9192262 | 0.30919681 | 457.97 | ABCC1 | ATP binding cassette subfamily C member 1 |
| 1565 | 0.0216 | 0.00358 | 0.264 | 2.9128214 | 0.76768797 | 155.6 | CYP2D6 | cytochrome P450 family 2 subfamily D member 6 |
| 1551 | 0.0224 | 0.00378 | 0.288 | 2.8958891 | 0.83387099 | 10.09 | CYP3A7 | cytochrome P450 family 3 subfamily A member 7 |
| 1576 | 0.0237 | 0.00408 | 0.229 | 2.8720505 | 0.65878675 | 22.05 | CYP3A4 | cytochrome P450 family 3 subfamily A member 4 |
| 22901 | 0.0292 | 0.00539 | 0.142 | -2.782875 | -0.39633775 | 222.07 | ARSG | arylsulfatase G |
| 4051 | 0.0293 | 0.00542 | 0.382 | -2.780778 | -1.06317033 | 43.07 | CYP4F3 | cytochrome P450 family 4 subfamily F member 3 |
| 2729 | 0.032 | 0.00613 | 0.316 | -2.740975 | -0.86578013 | 752.52 | GCLC | glutamate-cysteine ligase catalytic subunit |
| 627 | 0.0345 | 0.00685 | 0.268 | -2.703835 | -0.72506924 | 84.68 | BDNF | brain derived neurotrophic factor |
| 7450 | 0.0465 | 0.0104 | 0.319 | -2.562553 | -0.81654201 | 4017.58 | VWF | von Willebrand factor |

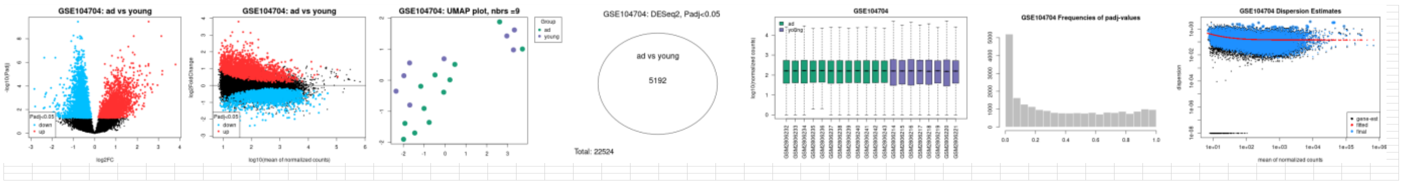

Supplementary Table S4

| | | | | GSE104704 AD V OLD | | | | |
|---|---|---|---|---|---|---|---|---|
| GeneID | padj | pvalue | lfcSE | stat | log2FoldChange | baseMean | Symbol | Description |
| 8701 | 0.00027 | 1.4E-06 | 0.214 | 4.82859714 | 1.03 | 93 | DNAH11 | dynein axonemal heavy chain 11 |
| 1571 | 0.00027 | 1.4E-06 | 0.159 | -4.8276574 | -0.768 | 85.16 | CYP2E1 | cytochrome P450 family 2 subfamily E member 1 |
| 1646 | 0.00056 | 4.3E-06 | 0.217 | 4.59734526 | 0.996 | 95 | AKR1C2 | aldo-keto reductase family 1 member C2 |
| 1558 | 0.00065 | 5.7E-06 | 0.22 | -4.5383884 | -0.996 | 24.33 | CYP2C8 | cytochrome P450 family 2 subfamily C member 8 |
| 1244 | 0.0013 | 1.7E-05 | 0.3 | 4.29753651 | 1.29 | 26.9 | ABCC2 | ATP binding cassette subfamily C member 2 |
| 27134 | 0.00132 | 1.8E-05 | 0.225 | 4.29301692 | 0.967 | 55.47 | TJP3 | tight junction protein 3 |
| 627 | 0.00177 | 2.7E-05 | 0.23 | -4.198984 | -0.966 | 99.06 | BDNF | brain derived neurotrophic factor |
| 57576 | 0.00566 | 0.00016 | 0.211 | -3.7744999 | -0.795 | 325.39 | KIF17 | kinesin family member 17 |
| 57834 | 0.0113 | 0.00047 | 0.267 | -3.4966646 | -0.934 | 217.89 | CYP4F11 | cytochrome P450 family 4 subfamily F member 11 |
| 2937 | 0.0116 | 0.00049 | 0.157 | -3.4848257 | -0.546 | 267.37 | GSS | glutathione synthetase |
| 57530 | 0.032 | 0.00249 | 0.274 | 3.02503276 | 0.83 | 238.68 | CGN | cingulin |
| 25830 | 0.0343 | 0.0028 | 0.269 | -2.9885148 | -0.802 | 1501.26 | SULT4A1 | sulfotransferase family 4A member 1 |
| 2879 | 0.0347 | 0.00286 | 0.107 | -2.9825911 | -0.319 | 1365.28 | GPX4 | glutathione peroxidase 4 |
| 22901 | 0.0396 | 0.0036 | 0.132 | -2.9112456 | -0.383 | 231.13 | ARSG | arylsulfatase G |
| 348 | 0.0479 | 0.00497 | 0.154 | -2.8090319 | -0.431 | 8536.54 | APOE | apolipoprotein E |
| 7450 | 0.0498 | 0.00532 | 0.216 | -2.787117 | -0.602 | 3917.22 | VWF | von Willebrand factor |

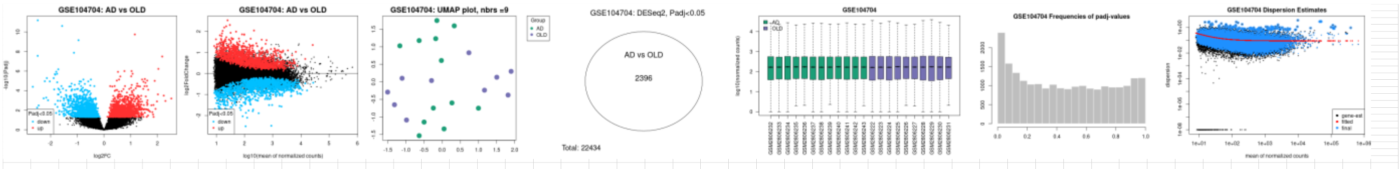

### Supplementary Table S5

| | | | | GSE104704 AD V CTL | | | | |
|---|---|---|---|---|---|---|---|---|
| GeneID | padj | pvalue | lfcSE | stat | log2FoldChange | baseMean | Symbol | Description |
| 8701 | 4.3E-07 | 9.6E-11 | 0.178 | 6.473973 | 1.1505933 | 81.15 | DNAH11 | dynein axonemal heavy chain 11 |
| 2937 | 3.1E-05 | 8.3E-08 | 0.134 | -5.36124 | -0.7193317 | 306.8 | GSS | glutathione synthetase |
| 1244 | 0.00057 | 9.1E-06 | 0.26 | 4.43762 | 1.1541335 | 24.67 | ABCC2 | ATP binding cassette subfamily C member 2 |
| 27134 | 0.00071 | 1.3E-05 | 0.218 | 4.365133 | 0.9521263 | 50.63 | TJP3 | tight junction protein 3 |
| 627 | 0.00127 | 3.1E-05 | 0.207 | -4.16279 | -0.8595717 | 102.78 | BDNF | brain derived neurotrophic factor |
| 1789 | 0.00655 | 0.00038 | 0.152 | 3.554873 | 0.541069 | 42.59 | DNMT3B | DNA methyltransferase 3 beta |
| 1646 | 0.00672 | 0.00039 | 0.216 | 3.54466 | 0.7642398 | 92.45 | AKR1C2 | aldo-keto reductase family 1 member C2 |
| 1595 | 0.00879 | 0.0006 | 0.193 | -3.43085 | -0.6626307 | 827.32 | CYP51A1 | cytochrome P450 family 51 subfamily A member 1 |
| 22901 | 0.0099 | 0.00073 | 0.114 | -3.3799 | -0.384584 | 240.07 | ARSG | arylsulfatase G |
| 55501 | 0.0165 | 0.00154 | 0.154 | 3.16659 | 0.4868202 | 851.07 | CHST12 | carbohydrate sulfotransferase 12 |
| 6818 | 0.0205 | 0.00216 | 0.19 | 3.066744 | 0.5815187 | 88.88 | SULT1A3 | sulfotransferase family 1A member 3 |
| 9446 | 0.0208 | 0.00221 | 0.168 | -3.06054 | -0.5146695 | 366.88 | GSTO1 | glutathione S-transferase omega 1 |
| 1571 | 0.0217 | 0.00236 | 0.166 | -3.04123 | -0.5059394 | 80.69 | CYP2E1 | cytochrome P450 family 2 subfamily E member 1 |
| 445329 | 0.0218 | 0.00236 | 0.191 | 3.040273 | 0.5816947 | 88.71 | SULT1A4 | sulfotransferase family 1A member 4 |
| 7450 | 0.0262 | 0.00313 | 0.236 | -2.95514 | -0.6976474 | 4349.53 | VWF | von Willebrand factor |
| 57530 | 0.0304 | 0.0039 | 0.252 | 2.88613 | 0.7281879 | 226.95 | CGN | cingulin |
| 51302 | 0.0357 | 0.00498 | 0.292 | 2.808322 | 0.818512 | 25.22 | CYP39A1 | cytochrome P450 family 39 subfamily A member 1 |
| 1558 | 0.0385 | 0.00553 | 0.263 | -2.77448 | -0.7286005 | 23.26 | CYP2C8 | cytochrome P450 family 2 subfamily C member 8 |
| 1565 | 0.0463 | 0.00721 | 0.24 | 2.687048 | 0.6434314 | 150.4 | CYP2D6 | cytochrome P450 family 2 subfamily D member 6 |

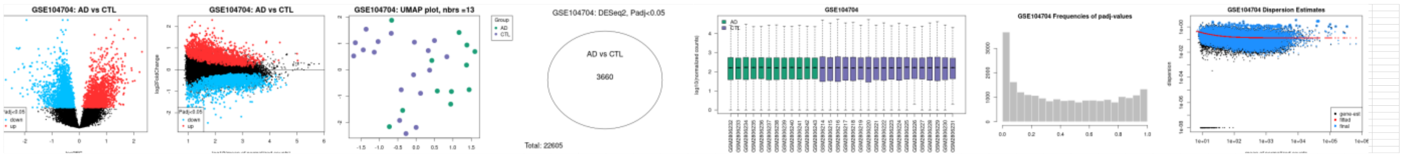

### Supplementary Table S6

| | | | | | | | | |
|---|---|---|---|---|---|---|---|---|
| | | | | GSE159699 AD V YOUNG | | | | |
| GeneID | padj | pvalue | lfcSE | stat | log2FoldChange | baseMean | Symbol | Description |
| 2937 | 2E-06 | 7.4E-09 | 0.159 | -5.782337 | -0.9197437 | 291.52 | GSS | glutathione synthetase |
| 8701 | 3.1E-06 | 1.5E-08 | 0.23 | 5.6658517 | 1.30216899 | 89.69 | DNAH11 | dynein axonemal heavy chain 11 |
| 9446 | 8.6E-06 | 5.9E-08 | 0.149 | -5.421512 | -0.80571845 | 368.61 | GSTO1 | glutathione S-transferase omega 1 |
| 1595 | 7.6E-05 | 1.4E-06 | 0.203 | -4.820848 | -0.97786544 | 826.86 | CYP51A1 | cytochrome P450 family 51 subfamily A member 1 |
| 1789 | 0.00099 | 5.1E-05 | 0.19 | 4.0520515 | 0.76820326 | 42.58 | DNMT3B | DNA methyltransferase 3 beta |
| 445329 | 0.00145 | 8.6E-05 | 0.233 | 3.9270585 | 0.91504295 | 87.42 | SULT1A4 | sulfotransferase family 1A member 4 |
| 6818 | 0.00147 | 8.7E-05 | 0.232 | 3.9244889 | 0.90949338 | 87.67 | SULT1A3 | sulfotransferase family 1A member 3 |
| 246 | 0.00183 | 0.00012 | 0.414 | 3.8499763 | 1.59184536 | 12.45 | ALOX15 | arachidonate 15-lipoxygenase |
| 6821 | 0.00379 | 0.00032 | 0.091 | -3.598222 | -0.32795855 | 1013.82 | SUOX | sulfite oxidase |
| 1244 | 0.00589 | 0.0006 | 0.289 | 3.433467 | 0.99287479 | 28.7 | ABCC2 | ATP binding cassette subfamily C member 2 |
| 147111 | 0.00659 | 0.0007 | 0.451 | 3.3900789 | 1.5291418 | 25.74 | NOTUM | notum, palmitoleoyl-protein carboxylesterase |
| 27134 | 0.00993 | 0.00123 | 0.279 | 3.2318003 | 0.90116284 | 56.4 | TJP3 | tight junction protein 3 |
| 9420 | 0.0106 | 0.00134 | 0.337 | -3.207404 | -1.07995875 | 645.56 | CYP7B1 | cytochrome P450 family 7 subfamily B member 1 |
| 2944 | 0.0164 | 0.00241 | 0.414 | 3.0344857 | 1.25524982 | 156.53 | GSTM1 | glutathione S-transferase mu 1 |
| 314 | 0.0181 | 0.00282 | 0.22 | 2.986215 | 0.65792596 | 75.54 | AOC2 | amine oxidase copper containing 2 |
| 55501 | 0.0182 | 0.00283 | 0.187 | 2.9851023 | 0.557181 | 870.96 | CHST12 | carbohydrate sulfotransferase 12 |
| 4363 | 0.0214 | 0.00354 | 0.106 | 2.9162256 | 0.30897278 | 457.97 | ABCC1 | ATP binding cassette subfamily C member 1 |
| 1565 | 0.0216 | 0.00359 | 0.264 | 2.9119103 | 0.7675055 | 155.6 | CYP2D6 | cytochrome P450 family 2 subfamily D member 6 |
| 1551 | 0.0225 | 0.0038 | 0.288 | 2.8946876 | 0.83363599 | 10.09 | CYP3A7 | cytochrome P450 family 3 subfamily A member 7 |
| 1576 | 0.0238 | 0.00409 | 0.229 | 2.8708229 | 0.65856007 | 22.05 | CYP3A4 | cytochrome P450 family 3 subfamily A member 4 |
| 22901 | 0.0289 | 0.00533 | 0.142 | -2.786101 | -0.39659432 | 222.06 | ARSG | arylsulfatase G |
| 4051 | 0.0293 | 0.00542 | 0.382 | -2.781129 | -1.06328814 | 43.07 | CYP4F3 | cytochrome P450 family 4 subfamily F member 3 |
| 2729 | 0.0318 | 0.0061 | 0.316 | -2.742093 | -0.86602653 | 752.51 | GCLC | glutamate-cysteine ligase catalytic subunit |
| 627 | 0.0344 | 0.00682 | 0.268 | -2.705344 | -0.72543485 | 84.68 | BDNF | brain derived neurotrophic factor |
| 7450 | 0.0465 | 0.0104 | 0.319 | -2.563469 | -0.81669385 | 4017.47 | VWF | von Willebrand factor |

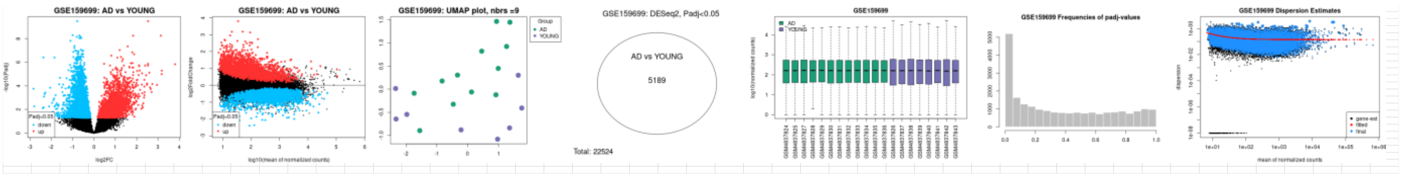

Supplementary Table S7

| | | | | GSE159699 AD V OLD | | | | |
|---|---|---|---|---|---|---|---|---|
| GeneID | padj | pvalue | lfcSE | stat | log2FoldChange | baseMean | Symbol | Description |
| 8701 | 0.00027 | 1.4E-06 | 0.214 | 4.8286952 | 1.03352419 | 93 | DNAH11 | dynein axonemal heavy chain 11 |
| 1571 | 0.00027 | 1.4E-06 | 0.159 | -4.826876 | -0.76844066 | 85.16 | CYP2E1 | cytochrome P450 family 2 subfamily E member 1 |
| 1646 | 0.00056 | 4.3E-06 | 0.217 | 4.5973205 | 0.99556639 | 95 | AKR1C2 | aldo-keto reductase family 1 member C2 |
| 1558 | 0.00065 | 5.7E-06 | 0.22 | -4.538102 | -0.99620229 | 24.33 | CYP2C8 | cytochrome P450 family 2 subfamily C member 8 |
| 1244 | 0.0013 | 1.7E-05 | 0.3 | 4.2976024 | 1.28882275 | 26.9 | ABCC2 | ATP binding cassette subfamily C member 2 |
| 27134 | 0.00132 | 1.8E-05 | 0.225 | 4.2928643 | 0.96666803 | 55.47 | TJP3 | tight junction protein 3 |
| 627 | 0.00177 | 2.7E-05 | 0.23 | -4.199083 | -0.965875 | 99.06 | BDNF | brain derived neurotrophic factor |
| 57576 | 0.00568 | 0.00016 | 0.211 | -3.774461 | -0.79540391 | 325.39 | KIF17 | kinesin family member 17 |
| 57834 | 0.0113 | 0.00047 | 0.267 | -3.496277 | -0.93421727 | 217.9 | CYP4F11 | cytochrome P450 family 4 subfamily F member 11 |
| 2937 | 0.0116 | 0.00049 | 0.157 | -3.484606 | -0.54565615 | 267.37 | GSS | glutathione synthetase |
| 57530 | 0.032 | 0.00249 | 0.274 | 3.0249465 | 0.82950902 | 238.68 | CGN | cingulin |
| 25830 | 0.0342 | 0.00281 | 0.269 | -2.988325 | -0.80234292 | 1501.27 | SULT4A1 | sulfotransferase family 4A member 1 |
| 2879 | 0.0346 | 0.00286 | 0.107 | -2.982445 | -0.31916326 | 1365.29 | GPX4 | glutathione peroxidase 4 |
| 22901 | 0.0392 | 0.00352 | 0.132 | -2.91812 | -0.38385736 | 231.18 | ARSG | arylsulfatase G |
| 348 | 0.0479 | 0.00496 | 0.154 | -2.809498 | -0.43127109 | 8536.48 | APOE | apolipoprotein E |
| 7450 | 0.0498 | 0.00532 | 0.216 | -2.787247 | -0.6023591 | 3917.23 | VWF | von Willebrand factor |

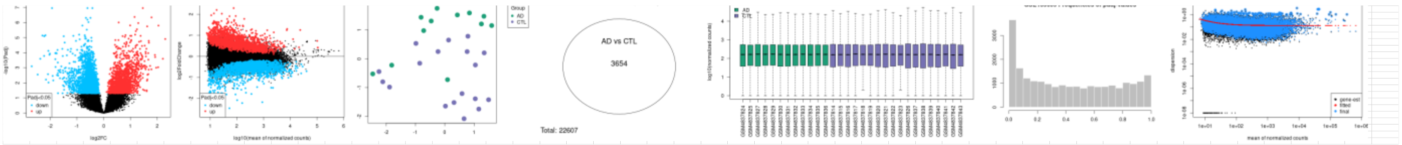

Supplementary Table S8

| | | | | GSE159699 ADV CTL | | | | |
|---|---|---|---|---|---|---|---|---|
| GeneID | padj | pvalue | lfcSE | stat | log2FoldChange | baseMean | Symbol | Description |
| 8701 | 4.3E-07 | 9.5E-11 | 0.178 | 6.474222 | 1.1505819 | 81.15 | DNAH11 | dynein axonemal heavy chain 11 |
| 2937 | 3E-05 | 8.3E-08 | 0.134 | -5.36101 | -0.7192924 | 306.8 | GSS | glutathione synthetase |
| 1244 | 0.00057 | 9.1E-06 | 0.26 | 4.437891 | 1.1541774 | 24.67 | ABCC2 | ATP binding cassette subfamily C member 2 |
| 27134 | 0.00071 | 1.3E-05 | 0.218 | 4.364769 | 0.9521552 | 50.63 | TJP3 | tight junction protein 3 |
| 627 | 0.00128 | 3.2E-05 | 0.207 | -4.16238 | -0.8595242 | 102.78 | BDNF | brain derived neurotrophic factor |
| 1789 | 0.00654 | 0.00038 | 0.152 | 3.554616 | 0.5411079 | 42.59 | DNMT3B | DNA methyltransferase 3 beta |
| 1646 | 0.00672 | 0.00039 | 0.216 | 3.544596 | 0.7642934 | 92.45 | AKR1C2 | aldo-keto reductase family 1 member C2 |
| 1595 | 0.00881 | 0.0006 | 0.193 | -3.4309 | -0.6625287 | 827.16 | CYP51A1 | cytochrome P450 family 51 subfamily A member 1 |
| 22901 | 0.00981 | 0.00071 | 0.114 | -3.38423 | -0.3848405 | 240.11 | ARSG | arylsulfatase G |
| 55501 | 0.0165 | 0.00154 | 0.154 | 3.166987 | 0.4869604 | 851.11 | CHST12 | carbohydrate sulfotransferase 12 |
| 6818 | 0.0205 | 0.00216 | 0.19 | 3.066682 | 0.5815381 | 88.88 | SULT1A3 | sulfotransferase family 1A member 3 |
| 9446 | 0.0208 | 0.00221 | 0.168 | -3.06044 | -0.5146061 | 366.87 | GSTO1 | glutathione S-transferase omega 1 |
| 1571 | 0.0218 | 0.00236 | 0.166 | -3.04058 | -0.505891 | 80.69 | CYP2E1 | cytochrome P450 family 2 subfamily E member 1 |
| 445329 | 0.0218 | 0.00236 | 0.191 | 3.040213 | 0.5817137 | 88.71 | SULT1A4 | sulfotransferase family 1A member 4 |
| 7450 | 0.0262 | 0.00313 | 0.236 | -2.9551 | -0.6976288 | 4349.51 | VWF | von Willebrand factor |
| 57530 | 0.0304 | 0.0039 | 0.252 | 2.886387 | 0.7282666 | 226.95 | CGN | cingulin |
| 51302 | 0.0357 | 0.00498 | 0.291 | 2.808592 | 0.8185481 | 25.22 | CYP39A1 | cytochrome P450 family 39 subfamily A member 1 |
| 1558 | 0.0385 | 0.00553 | 0.263 | -2.77413 | -0.728591 | 23.26 | CYP2C8 | cytochrome P450 family 2 subfamily C member 8 |
| 1565 | 0.0463 | 0.00721 | 0.24 | 2.687161 | 0.6434514 | 150.4 | CYP2D6 | cytochrome P450 family 2 subfamily D member 6 |

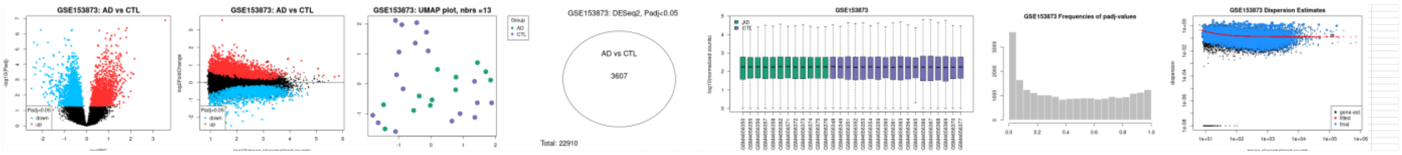

Supplementary Table S9

| | | | | GSE153873 AD V YOUNG | | | | |
|---|---|---|---|---|---|---|---|---|
| GeneID | padj | pvalue | lfcSE | stat | log2FoldChange | baseMean | Symbol | Description |
| 2937 | 1.1E-06 | 8.2E-10 | 0.16 | -6.1405 | -0.9813571 | 315.42 | GSS | glutathione synthetase |
| 8701 | 9.7E-06 | 3.4E-08 | 0.233 | 5.522274 | 1.2887631 | 105.74 | DNAH11 | dynein axonemal heavy chain 11 |
| 1595 | 0.00013 | 2.1E-06 | 0.222 | -4.7438 | -1.0523845 | 933.69 | CYP51A1 | cytochrome P450 family 51 subfamily A member 1 |
| 9446 | 0.00034 | 9.2E-06 | 0.165 | -4.43625 | -0.7304778 | 410.76 | GSTO1 | glutathione S-transferase omega 1 |
| 6818 | 0.00071 | 2.8E-05 | 0.223 | 4.192594 | 0.9352344 | 98.58 | SULT1A3 | sulfotransferase family 1A member 3 |
| 445329 | 0.00072 | 2.8E-05 | 0.222 | 4.188903 | 0.9285329 | 98.25 | SULT1A4 | sulfotransferase family 1A member 4 |
| 246 | 0.00088 | 3.8E-05 | 0.484 | 4.121772 | 1.99324 | 14.99 | ALOX15 | arachidonate 15-lipoxygenase |
| 27134 | 0.00192 | 0.00012 | 0.255 | 3.855199 | 0.9832502 | 70.28 | TJP3 | tight junction protein 3 |
| 55501 | 0.0034 | 0.00026 | 0.19 | 3.65297 | 0.6938023 | 964.08 | CHST12 | carbohydrate sulfotransferase 12 |
| 147111 | 0.00636 | 0.00062 | 0.45 | 3.421521 | 1.5399022 | 27.21 | NOTUM | notum, palmitoleoyl-protein carboxylesterase |
| 1565 | 0.00679 | 0.00069 | 0.244 | 3.394359 | 0.8269938 | 163.64 | CYP2D6 | cytochrome P450 family 2 subfamily D member 6 |
| 1789 | 0.00719 | 0.00074 | 0.226 | 3.373043 | 0.7625959 | 45.06 | DNMT3B | DNA methyltransferase 3 beta |
| 9420 | 0.00766 | 0.00081 | 0.349 | -3.34919 | -1.169187 | 736.05 | CYP7B1 | cytochrome P450 family 7 subfamily B member 1 |
| 84171 | 0.0103 | 0.00123 | 0.212 | 3.231018 | 0.6848384 | 57.89 | LOXL4 | lysyl oxidase like 4 |
| 627 | 0.0131 | 0.00174 | 0.254 | -3.13183 | -0.7955966 | 97.3 | BDNF | brain derived neurotrophic factor |
| 1244 | 0.0139 | 0.00188 | 0.315 | 3.109072 | 0.9796119 | 35.21 | ABCC2 | ATP binding cassette subfamily C member 2 |
| 2944 | 0.0142 | 0.00193 | 0.401 | 3.100769 | 1.2446348 | 180.15 | GSTM1 | glutathione S-transferase mu 1 |
| 22901 | 0.0149 | 0.00208 | 0.143 | -3.07839 | -0.4389725 | 254.26 | ARSG | arylsulfatase G |
| 1365 | 0.016 | 0.00229 | 0.477 | 3.050122 | 1.4555721 | 8.5 | CLDN3 | claudin 3 |
| 4363 | 0.0163 | 0.00233 | 0.107 | 3.044698 | 0.3266568 | 531.96 | ABCC1 | ATP binding cassette subfamily C member 1 |
| 6821 | 0.0169 | 0.00246 | 0.095 | -3.02809 | -0.2878072 | 1176.51 | SUOX | sulfite oxidase |
| 5621 | 0.0176 | 0.00261 | 0.202 | -3.01059 | -0.6085369 | 5268.48 | PRNP | prion protein |
| 2729 | 0.033 | 0.00624 | 0.333 | -2.73495 | -0.9114344 | 845.8 | GCLC | glutamate-cysteine ligase catalytic subunit |
| 314 | 0.0415 | 0.00862 | 0.22 | 2.626957 | 0.5783622 | 82.36 | AOC2 | amine oxidase copper containing 2 |
| 4051 | 0.0418 | 0.00868 | 0.377 | -2.62427 | -0.988613 | 41.88 | CYP4F3 | cytochrome P450 family 4 subfamily F member 3 |
| 6799 | 0.0458 | 0.00995 | 0.326 | 2.577681 | 0.8408976 | 66.02 | SULT1A2 | sulfotransferase family 1A member 2 |
| 4035 | 0.0472 | 0.0103 | 0.156 | 2.564125 | 0.3990087 | 19426.81 | LRP1 | LDL receptor related protein 1 |

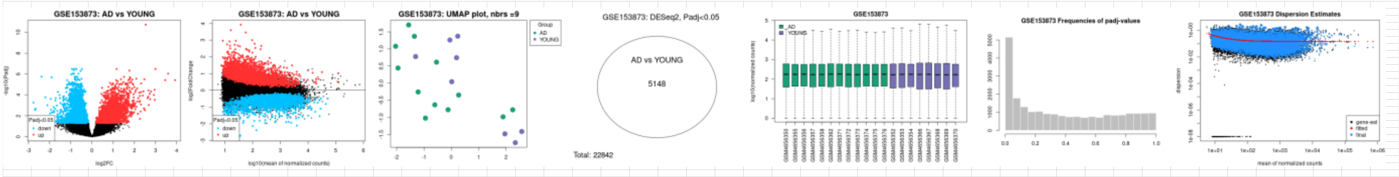

Supplementary Table S10

| | | | | GSE153873 AD V OLD | | | | |
|---|---|---|---|---|---|---|---|---|
| GeneID | padj | pvalue | lfcSE | stat | log2FoldChange | baseMean | Symbol | Description |
| 27134 | 5.2E-06 | 7.3E-09 | 0.209 | 5.784265 | 1.2093561 | 64.74 | TJP3 | tight junction protein 3 |
| 1646 | 0.00016 | 7E-07 | 0.225 | 4.960249 | 1.1159442 | 112.65 | AKR1C2 | aldo-keto reductase family 1 member C2 |
| 8701 | 0.00046 | 2.8E-06 | 0.212 | 4.683518 | 0.9938782 | 106.75 | DNAH11 | dynein axonemal heavy chain 11 |
| 627 | 0.00054 | 3.7E-06 | 0.219 | -4.62797 | -1.0123992 | 109.4 | BDNF | brain derived neurotrophic factor |
| 1244 | 0.00113 | 1.3E-05 | 0.256 | 4.368995 | 1.11841 | 32.96 | ABCC2 | ATP binding cassette subfamily C member 2 |
| 1571 | 0.00178 | 2.4E-05 | 0.158 | -4.21998 | -0.6678795 | 92.79 | CYP2E1 | cytochrome P450 family 2 subfamily E member 1 |
| 2937 | 0.0031 | 5.8E-05 | 0.155 | -4.02198 | -0.6216858 | 281.33 | GSS | glutathione synthetase |
| 51302 | 0.0062 | 0.00017 | 0.326 | 3.765269 | 1.2275517 | 31.34 | CYP39A1 | cytochrome P450 family 39 subfamily A member 1 |
| 57576 | 0.00771 | 0.00024 | 0.221 | -3.67843 | -0.8118031 | 352.84 | KIF17 | kinesin family member 17 |
| 112869 | 0.00955 | 0.00033 | 0.13 | 3.589399 | 0.464883 | 138.56 | SGF29 | SAGA complex associated factor 29 |
| 57834 | 0.0135 | 0.00058 | 0.265 | -3.44224 | -0.9121822 | 235.79 | CYP4F11 | cytochrome P450 family 4 subfamily F member 11 |
| 260293 | 0.0189 | 0.00099 | 0.201 | -3.29263 | -0.6632786 | 164.17 | CYP4X1 | cytochrome P450 family 4 subfamily X member 1 |
| 22901 | 0.0269 | 0.00171 | 0.124 | -3.13703 | -0.3901225 | 252.96 | ARSG | arylsulfatase G |
| 875 | 0.0402 | 0.00341 | 0.162 | 2.928014 | 0.4736338 | 766.96 | CBS | cystathionine beta-synthase |
| 57530 | 0.0419 | 0.00365 | 0.25 | 2.906973 | 0.7278947 | 276.79 | CGN | cingulin |
| 4843 | 0.0438 | 0.00399 | 0.288 | -2.87919 | -0.8303568 | 88.11 | NOS2 | nitric oxide synthase 2 |
| 1767 | 0.0461 | 0.0045 | 0.139 | -2.8406 | -0.3940042 | 138.39 | DNAH5 | dynein axonemal heavy chain 5 |
| 2879 | 0.0471 | 0.00467 | 0.119 | -2.82903 | -0.3356385 | 1417.15 | GPX4 | glutathione peroxidase 4 |
| 57404 | 0.0479 | 0.00482 | 0.169 | 2.818756 | 0.4755657 | 373.48 | CYP20A1 | cytochrome P450 family 20 subfamily A member 1 |

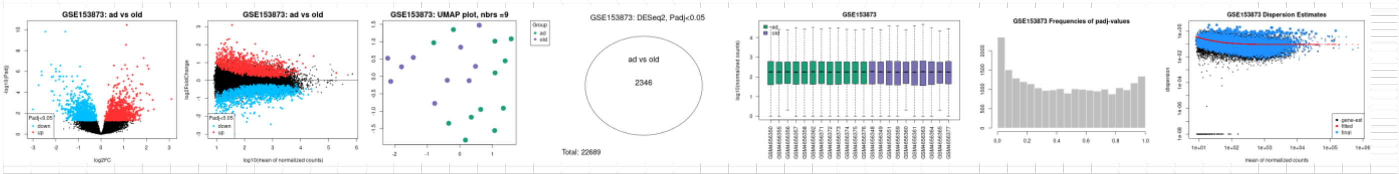

Supplementary Table S11

| | | | | | | | | |
|---|---|---|---|---|---|---|---|---|
| | | | | GSE153873 AD V CTL | | | | |
| GeneID | padj | pvalue | lfcSE | stat | log2FoldChange | baseMean | Symbol | Description |
| 8701 | 1.3E-06 | 6.8E-10 | 0.182 | 6.171442 | 1.1237569 | 94.9 | DNAH11 | dynein axonemal heavy chain 11 |
| 2937 | 2.5E-06 | 2.1E-09 | 0.131 | -5.98767 | -0.7870779 | 330.33 | GSS | glutathione synthetase |
| 27134 | 6.1E-06 | 7.6E-09 | 0.194 | 5.776564 | 1.1222307 | 60.05 | TJP3 | tight junction protein 3 |
| 627 | 0.00044 | 4.9E-06 | 0.2 | -4.56841 | -0.9137899 | 116.6 | BDNF | brain derived neurotrophic factor |
| 1244 | 0.00123 | 2.4E-05 | 0.251 | 4.223301 | 1.0597836 | 30.62 | ABCC2 | ATP binding cassette subfamily C member 2 |
| 1646 | 0.00404 | 0.00015 | 0.224 | 3.787941 | 0.8467457 | 111.45 | AKR1C2 | aldo-keto reductase family 1 member C2 |
| 22901 | 0.00532 | 0.00024 | 0.111 | -3.66829 | -0.4061388 | 269.76 | ARSG | arylsulfatase G |
| 55501 | 0.00807 | 0.00047 | 0.159 | 3.496671 | 0.5573379 | 932.14 | CHST12 | carbohydrate sulfotransferase 12 |
| 51302 | 0.00817 | 0.00048 | 0.286 | 3.492548 | 0.9973951 | 30.26 | CYP39A1 | cytochrome P450 family 39 subfamily A member 1 |
| 1595 | 0.00986 | 0.00066 | 0.205 | -3.40498 | -0.69849 | 909.51 | CYP51A1 | cytochrome P450 family 51 subfamily A member 1 |
| 1565 | 0.0175 | 0.00156 | 0.222 | 3.163625 | 0.7015054 | 154.69 | CYP2D6 | cytochrome P450 family 2 subfamily D member 6 |
| 57530 | 0.0263 | 0.00287 | 0.231 | 2.980993 | 0.6889252 | 265.66 | CGN | cingulin |
| 6818 | 0.027 | 0.003 | 0.191 | 2.967903 | 0.5667552 | 99.55 | SULT1A3 | sulfotransferase family 1A member 3 |
| 445329 | 0.0274 | 0.00307 | 0.19 | 2.96058 | 0.5637849 | 99.2 | SULT1A4 | sulfotransferase family 1A member 4 |
| 1789 | 0.0303 | 0.00362 | 0.188 | 2.909844 | 0.5482668 | 44.33 | DNMT3B | DNA methyltransferase 3 beta |
| 875 | 0.0316 | 0.00386 | 0.159 | 2.889548 | 0.4595639 | 748.37 | CBS | cystathionine beta-synthase |
| 112869 | 0.0342 | 0.00432 | 0.124 | 2.85407 | 0.3544911 | 140.25 | SGF29 | SAGA complex associated factor 29 |
| 5621 | 0.0349 | 0.00447 | 0.165 | -2.84258 | -0.4681491 | 5431.94 | PRNP | prion protein |
| 7450 | 0.0491 | 0.00766 | 0.234 | -2.66657 | -0.6238546 | 4969.11 | VWF | von Willebrand factor |

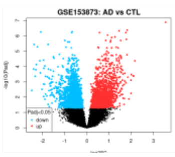

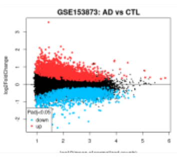

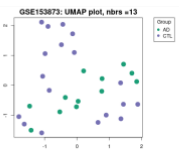

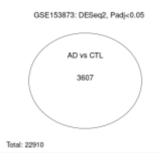

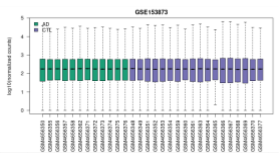

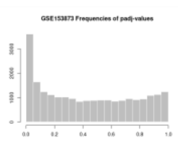

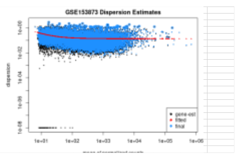

Supplementary Table S12

| | | | | | GSE173955 AD V CTL | | | |
|---|---|---|---|---|---|---|---|---|
| GeneID | padj | pvalue | lfcSE | stat | log2FoldChange | baseMean | Symbol | Description |
| 9414 | 0.0003806 | 3.7E-06 | 0.341 | 4.630195 | 1.5773302 | 2757.49 | TJP2 | tight junction protein 2 |
| 2903 | 0.0014415 | 3.3E-05 | 0.319 | -4.15248 | -1.3263385 | 2327.32 | GRIN2A | glutamate ionotropic receptor NMDA type subunit 2A |
| 412 | 0.0017082 | 4.7E-05 | 0.258 | -4.0727 | -1.0523557 | 859.35 | STS | steroid sulfatase |
| 66002 | 0.0019501 | 5.9E-05 | 0.584 | 4.017238 | 2.344321 | 107.64 | CYP4F12 | cytochrome P450 family 4 subfamily F member 12 |
| 7082 | 0.0036457 | 0.00018 | 0.165 | 3.743242 | 0.6179233 | 4089.51 | TJP1 | tight junction protein 1 |
| 11182 | 0.0039133 | 0.00021 | 0.181 | -3.71238 | -0.6704225 | 289.51 | SLC2A6 | solute carrier family 2 member 6 |
| 1080 | 0.0042885 | 0.00024 | 0.532 | 3.676011 | 1.9565934 | 38.13 | CFTR | CF transmembrane conductance regulator |
| 113189 | 0.0055415 | 0.00036 | 0.248 | 3.56684 | 0.886058 | 132.36 | CHST14 | carbohydrate sulfotransferase 14 |
| 1244 | 0.0057262 | 0.00039 | 0.29 | 3.549563 | 1.0285841 | 22.38 | ABCC2 | ATP binding cassette subfamily C member 2 |
| 25830 | 0.0057968 | 0.00039 | 0.375 | -3.54502 | -1.327993 | 4389.05 | SULT4A1 | sulfotransferase family 4A member 1 |
| 56603 | 0.0063487 | 0.00047 | 0.384 | -3.49777 | -1.3417971 | 576 | CYP26B1 | cytochrome P450 family 26 subfamily B member 1 |
| 1543 | 0.0078404 | 0.00066 | 0.484 | -3.40729 | -1.6489571 | 25.13 | CYP1A1 | cytochrome P450 family 1 subfamily A member 1 |
| 4489 | 0.009695 | 0.00092 | 0.672 | 3.312672 | 2.224748 | 91.4 | MT1A | metallothionein 1A |
| 2328 | 0.0113029 | 0.0012 | 0.389 | 3.239654 | 1.2592228 | 33.92 | FMO3 | flavin containing dimethylaniline monoxygenase 3 |
| 1789 | 0.0124547 | 0.00138 | 0.175 | 3.198368 | 0.5586502 | 70.17 | DNMT3B | DNA methyltransferase 3 beta |
| 80173 | 0.0144264 | 0.00172 | 0.167 | 3.134476 | 0.5247981 | 423.68 | IFT74 | intraflagellar transport 74 |
| 324 | 0.0149233 | 0.00181 | 0.206 | -3.12016 | -0.6426039 | 6057.37 | APC | APC regulator of WNT signaling pathway |
| 5621 | 0.0153169 | 0.00189 | 0.214 | -3.10662 | -0.665284 | 15263.18 | PRNP | prion protein |
| 4051 | 0.0171753 | 0.00227 | 0.499 | 3.051982 | 1.5241404 | 75 | CYP4F3 | cytochrome P450 family 4 subfamily F member 3 |
| 6799 | 0.0181977 | 0.00248 | 0.219 | 3.02569 | 0.6615866 | 52.34 | SULT1A2 | sulfotransferase family 1A member 2 |
| 4301 | 0.0203256 | 0.00291 | 0.111 | 2.976783 | 0.3308368 | 2074.24 | AFDN | afadin, adherens junction formation factor |
| 627 | 0.0205867 | 0.00298 | 0.39 | -2.96963 | -1.1569112 | 368.74 | BDNF | brain derived neurotrophic factor |
| 1573 | 0.0228163 | 0.0035 | 0.241 | 2.920235 | 0.7038384 | 523.56 | CYP2J2 | cytochrome P450 family 2 subfamily J member 2 |
| 1562 | 0.023288 | 0.00364 | 0.566 | -2.90762 | -1.6463837 | 10.17 | CYP2C18 | cytochrome P450 family 2 subfamily C member 18 |
| 10858 | 0.0283428 | 0.00494 | 0.32 | -2.81104 | -0.8996539 | 1942.54 | CYP46A1 | cytochrome P450 family 46 subfamily A member 1 |
| 57404 | 0.0290504 | 0.00516 | 0.221 | 2.797073 | 0.6178102 | 233.9 | CYP20A1 | cytochrome P450 family 20 subfamily A member 1 |
| 2904 | 0.0293104 | 0.00524 | 0.406 | -2.79218 | -1.1332207 | 2722.25 | GRIN2B | glutamate ionotropic receptor NMDA type subunit 2B |
| 6584 | 0.0333842 | 0.00644 | 0.201 | 2.72467 | 0.5482584 | 310.5 | SLC22A5 | solute carrier family 22 member 5 |
| 253152 | 0.0380073 | 0.00787 | 0.448 | -2.65765 | -1.1913441 | 153.3 | EPHX4 | epoxide hydrolase 4 |
| 58494 | 0.0387578 | 0.00811 | 0.218 | 2.647575 | 0.5764487 | 1150.71 | JAM2 | junctional adhesion molecule 2 |
| 7122 | 0.0401323 | 0.00858 | 0.456 | 2.628438 | 1.1990214 | 868.04 | CLDN5 | claudin 5 |
| 1588 | 0.0420447 | 0.00919 | 0.58 | 2.604776 | 1.5099389 | 14.27 | CYP19A1 | cytochrome P450 family 19 subfamily A member 1 |
| 1593 | 0.0428409 | 0.00949 | 0.318 | 2.593968 | 0.8247284 | 910.11 | CYP27A1 | cytochrome P450 family 27 subfamily A member 1 |
| 2990 | 0.0442756 | 0.01 | 0.198 | 2.57578 | 0.5097043 | 533.12 | GUSB | glucuronidase beta |
| 1545 | 0.0457205 | 0.0105 | 0.446 | 2.559864 | 1.1416078 | 351.78 | CYP1B1 | cytochrome P450 family 1 subfamily B member 1 |
| 158 | 0.0457517 | 0.0105 | 0.139 | -2.55941 | -0.3544486 | 423.01 | ADSL | adenylosuccinate lyase |
| 4851 | 0.0481565 | 0.0113 | 0.268 | 2.533251 | 0.6792334 | 672.47 | NOTCH1 | notch receptor 1 |
| 2944 | 0.0496878 | 0.0118 | 0.46 | 2.516934 | 1.1569915 | 1023.31 | GSTM1 | glutathione S-transferase mu 1 |

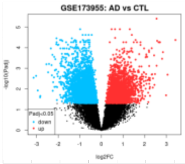

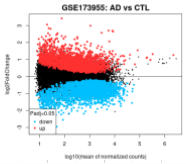

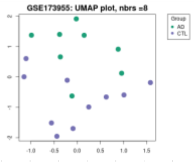

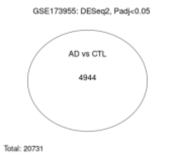

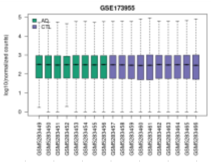

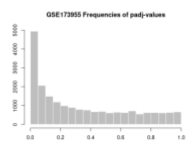

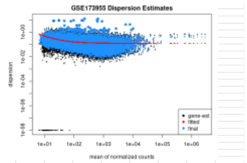

### Supplementary Table 13

| | | | | | GSE211993 L150P V CTL PART 1 | | | |
|---|---|---|---|---|---|---|---|---|
| GeneID | padj | pvalue | lfcSE | stat | log2FoldChange | baseMean | Symbol | Description |
| 1592 | 1.6E-125 | 2.5E-127 | 0.189 | 24.004654 | 4.5238266 | 98.17 | CYP26A1 | cytochrome P450 family 26 subfamily A member 1 |
| 260293 | 2.4E-120 | 4E-122 | 0.097 | 23.501271 | 2.2698525 | 315.94 | CYP4X1 | cytochrome P450 family 4 subfamily X member 1 |
| 5243 | 1.9E-109 | 3.5E-111 | 0.088 | -22.40518 | -1.9613046 | 552.8 | ABCB1 | ATP binding cassette subfamily B member 1 |
| 5649 | 2.6E-109 | 4.8E-111 | 0.088 | 22.390759 | 1.9597718 | 17243.54 | RELN | reelin |
| 1812 | 1.3E-100 | 2.7E-102 | 0.168 | 21.474592 | 3.6099965 | 89.2 | DRD1 | dopamine receptor D1 |
| 5621 | 3.3E-100 | 7.1E-102 | 0.042 | -21.42967 | -0.8992471 | 20187.1 | PRNP | prion protein |
| 2053 | 3.82E-91 | 9.26E-93 | 0.075 | 20.428895 | 1.5374168 | 362.26 | EPHX2 | epoxide hydrolase 2 |
| 57626 | 7.34E-59 | 2.89E-60 | 0.146 | 16.374925 | 2.3903066 | 663.52 | KLHL1 | kelch like family member 1 |
| 7082 | 3.06E-53 | 1.34E-54 | 0.058 | -15.5608 | -0.8965351 | 7860.72 | TJP1 | tight junction protein 1 |
| 224 | 1.67E-51 | 7.57E-53 | 0.042 | -15.30065 | -0.641262 | 6070.11 | ALDH3A2 | aldehyde dehydrogenase 3 family member A2 |
| 1789 | 3.66E-46 | 1.83E-47 | 0.074 | 14.471791 | 1.0719128 | 191.2 | DNMT3B | DNA methyltransferase 3 beta |
| 5742 | 1.24E-41 | 6.83E-43 | 0.318 | 13.728753 | 4.3657347 | 65.67 | PTGS1 | prostaglandin-endoperoxide synthase 1 |
| 51302 | 1.56E-40 | 8.82E-42 | 0.152 | 13.542099 | 2.0540769 | 102.1 | CYP39A1 | cytochrome P450 family 39 subfamily A member 1 |
| 7498 | 2.28E-39 | 1.31E-40 | 0.277 | 13.342357 | 3.6885285 | 61.22 | XDH | xanthine dehydrogenase |
| 788 | 7.13E-38 | 4.31E-39 | 0.071 | -13.07958 | -0.9219113 | 793.77 | SLC25A20 | solute carrier family 25 member 20 |
| 6783 | 8.61E-32 | 6.13E-33 | 0.412 | 11.954727 | 4.9215924 | 24.31 | SULT1E1 | sulfotransferase family 1E member 1 |
| 56603 | 1.69E-29 | 1.28E-30 | 0.338 | 11.502422 | 3.8917963 | 247.62 | CYP26B1 | cytochrome P450 family 26 subfamily B member 1 |
| 4301 | 5.72E-26 | 4.84E-27 | 0.051 | 10.768604 | 0.5519225 | 7835.52 | AFDN | afadin, adherens junction formation factor |
| 246 | 6.09E-26 | 5.17E-27 | 0.52 | 10.762507 | 5.5974638 | 22.96 | ALOX15 | arachidonate 15-lipoxygenase |
| 642446 | 1.04E-23 | 9.59E-25 | 0.305 | 10.270279 | 3.1360497 | 25.7 | TRIM64B | tripartite motif containing 64B |
| 9429 | 1.76E-23 | 1.63E-24 | 0.165 | -10.21889 | -1.683636 | 67.01 | ABCG2 | ATP binding cassette subfamily G member 2 (Junior blood group) |
| 4851 | 8.53E-22 | 8.45E-23 | 0.117 | -9.828917 | -1.151461 | 4402.89 | NOTCH1 | notch receptor 1 |
| 7122 | 8.17E-21 | 8.42E-22 | 0.435 | 9.594618 | 4.1745344 | 22.51 | CLDN5 | claudin 5 |
| 4502 | 9.72E-21 | 1.01E-21 | 0.219 | -9.576308 | -2.0924167 | 201.39 | MT2A | metallothionein 2A |
| 627 | 2.48E-20 | 2.6E-21 | 0.119 | 9.477531 | 1.1254602 | 658.98 | BDNF | brain derived neurotrophic factor |
| 216 | 3.7E-20 | 3.91E-21 | 0.2 | 9.434929 | 1.8897137 | 43.92 | ALDH1A1 | aldehyde dehydrogenase 1 family member A1 |
| 113612 | 8.02E-20 | 8.6E-21 | 0.057 | -9.352001 | -0.5347588 | 2364.99 | CYP2U1 | cytochrome P450 family 2 subfamily U member 1 |
| 8701 | 4.5E-19 | 4.96E-20 | 0.156 | 9.16482 | 1.4273309 | 283.47 | DNAH11 | dynein axonemal heavy chain 11 |
| 1558 | 8.79E-19 | 9.81E-20 | 0.315 | 9.091034 | 2.8651137 | 31.89 | CYP2C8 | cytochrome P450 family 2 subfamily C member 8 |
| 9446 | 1.64E-16 | 2.06E-17 | 0.063 | -8.49052 | -0.5353481 | 1290.4 | GSTO1 | glutathione S-transferase omega 1 |
| 221 | 3.21E-16 | 4.09E-17 | 0.196 | 8.410342 | 1.6455541 | 204.64 | ALDH3B1 | aldehyde dehydrogenase 3 family member B1 |
| 653689 | 1.55E-15 | 2.05E-16 | 0.39 | -8.218921 | -3.2050841 | 13.14 | GSTT2B | glutathione S-transferase theta 2B |
| 339761 | 1.94E-14 | 2.73E-15 | 0.171 | 7.90289 | 1.350897 | 60.88 | CYP27C1 | cytochrome P450 family 27 subfamily C member 1 |
| 348 | 5.39E-14 | 7.78E-15 | 0.22 | 7.771051 | 1.7123112 | 967.76 | APOE | apolipoprotein E |
| 57576 | 1.93E-13 | 2.88E-14 | 0.149 | -7.603516 | -1.1311731 | 151.6 | KIF17 | kinesin family member 17 |
| 340665 | 3.69E-13 | 5.59E-14 | 0.682 | 7.517306 | 5.1260895 | 9.31 | CYP26C1 | cytochrome P450 family 26 subfamily C member 1 |
| 25830 | 1.99E-12 | 3.12E-13 | 0.115 | 7.289045 | 0.8390737 | 785.48 | SULT4A1 | sulfotransferase family 4A member 1 |
| 3077 | 2.47E-12 | 3.9E-13 | 0.129 | 7.259102 | 0.9385924 | 162.79 | HFE | homeostatic iron regulator |
| 58494 | 2.74E-12 | 4.34E-13 | 0.159 | -7.244446 | -1.1500068 | 7674.91 | JAM2 | junctional adhesion molecule 2 |
| 7450 | 1.19E-11 | 1.95E-12 | 0.314 | -7.037756 | -2.2070097 | 10.12 | VWF | von Willebrand factor |

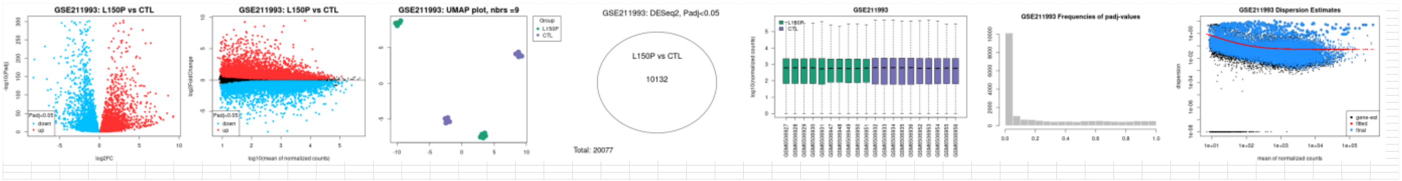

Supplementary Table 14

| | | | | | | | | |
|---|---|---|---|---|---|---|---|---|
| | | | | | GSE211993 L150P V CTL PART 2 | | | |
| 1545 | 1.79E-11 | 2.98E-12 | 0.081 | -6.978894 | -0.5652695 | 2030 | CYP1B1 | cytochrome P450 family 1 subfamily B member 1 |
| 1767 | 5.92E-11 | 1.02E-11 | 0.146 | -6.803663 | -0.9906178 | 801.68 | DNAH5 | dynein axonemal heavy chain 5 |
| 2903 | 9.16E-10 | 1.73E-10 | 0.057 | -6.38377 | -0.3616516 | 5922.78 | GRIN2A | glutamate ionotropic receptor NMDA type subunit 2A |
| 6584 | 3.08E-09 | 6.01E-10 | 0.097 | 6.190058 | 0.6030269 | 423.74 | SLC22A5 | solute carrier family 22 member 5 |
| 57530 | 1.3E-08 | 2.65E-09 | 0.105 | 5.951756 | 0.6235167 | 224.54 | CGN | cingulin |
| 7421 | 2.88E-08 | 6.08E-09 | 0.343 | -5.814646 | -1.99658 | 23.37 | VDR | vitamin D receptor |
| 1646 | 4.6E-08 | 9.91E-09 | 0.072 | 5.732324 | 0.4100751 | 1717.88 | AKR1C2 | aldo-keto reductase family 1 member C2 |
| 10587 | 3.37E-07 | 7.84E-08 | 0.204 | 5.370668 | 1.0930241 | 222.57 | TXNRD2 | thioredoxin reductase 2 |
| 9414 | 5.64E-07 | 1.34E-07 | 0.085 | 5.273417 | 0.4495014 | 625.94 | TJP2 | tight junction protein 2 |
| 6818 | 3.16E-06 | 8.06E-07 | 0.148 | 4.933803 | 0.7320968 | 36.76 | SULT1A3 | sulfotransferase family 1A member 3 |
| 445329 | 5.82E-06 | 1.53E-06 | 0.153 | 4.807486 | 0.7339983 | 36.61 | SULT1A4 | sulfotransferase family 1A member 4 |
| 210 | 1.55E-05 | 4.28E-06 | 0.229 | -4.597358 | -1.0503313 | 1541.49 | ALAD | aminolevulinate dehydratase |
| 1589 | 0.00003 | 8.58E-06 | 0.317 | -4.450269 | -1.4106203 | 21.43 | CYP21A2 | cytochrome P450 family 21 subfamily A member 2 |
| 6999 | 7.17E-05 | 2.15E-05 | 0.163 | 4.248817 | 0.6902587 | 52.4 | TDO2 | tryptophan 2,3-dioxygenase |
| 4854 | 9.21E-05 | 0.000028 | 0.327 | -4.188778 | -1.3703123 | 3295.32 | NOTCH3 | notch receptor 3 |
| 2730 | 0.000109 | 3.33E-05 | 0.102 | -4.149644 | -0.4220455 | 2008.23 | GCLM | glutamate-cysteine ligase modifier subunit |
| 27284 | 0.00014 | 4.34E-05 | 0.351 | 4.088517 | 1.4341512 | 13.84 | SULT1B1 | sulfotransferase family 1B member 1 |
| 2729 | 0.000152 | 4.75E-05 | 0.085 | -4.067678 | -0.344292 | 2245.29 | GCLC | glutamate-cysteine ligase catalytic subunit |
| 120227 | 0.000229 | 7.32E-05 | 0.102 | 3.965508 | 0.4032803 | 592.39 | CYP2R1 | cytochrome P450 family 2 subfamily R member 1 |
| 6948 | 0.000263 | 8.48E-05 | 0.142 | -3.930442 | -0.5581444 | 1461.01 | TCN2 | transcobalamin 2 |
| 8140 | 0.000314 | 0.000102 | 0.3 | -3.885019 | -1.1635126 | 3854.38 | SLC7A5 | solute carrier family 7 member 5 |
| 4363 | 0.000326 | 0.000106 | 0.187 | -3.875691 | -0.7259256 | 1720.06 | ABCC1 | ATP binding cassette subfamily C member 1 |
| 2876 | 0.000434 | 0.000144 | 0.164 | -3.800946 | -0.6247325 | 2632.36 | GPX1 | glutathione peroxidase 1 |
| 6819 | 0.0014 | 0.000506 | 0.316 | 3.477351 | 1.0975422 | 11.13 | SULT1C2 | sulfotransferase family 1C member 2 |
| 4843 | 0.00144 | 0.000519 | 0.188 | 3.470931 | 0.6529928 | 29.9 | NOS2 | nitric oxide synthase 2 |
| 22901 | 0.00342 | 0.00132 | 0.068 | -3.212355 | -0.2181975 | 682.46 | ARSG | arylsulfatase G |
| 84171 | 0.00352 | 0.00136 | 0.218 | -3.20323 | -0.6986015 | 38.64 | LOXL4 | lysyl oxidase like 4 |
| 1577 | 0.00387 | 0.00151 | 0.12 | -3.173431 | -0.379139 | 167.63 | CYP3A5 | cytochrome P450 family 3 subfamily A member 5 |
| 4524 | 0.004 | 0.00157 | 0.107 | 3.162308 | 0.3366611 | 1008.65 | MTHFR | methylenetetrahydrofolate reductase |
| 2937 | 0.00623 | 0.00253 | 0.085 | -3.019947 | -0.2573169 | 1628.58 | GSS | glutathione synthetase |
| 1593 | 0.0114 | 0.00489 | 0.186 | 2.814173 | 0.5230409 | 136.94 | CYP27A1 | cytochrome P450 family 27 subfamily A member 1 |
| 2948 | 0.0121 | 0.00523 | 0.269 | -2.792373 | -0.7507402 | 427.46 | GSTM4 | glutathione S-transferase mu 4 |
| 4255 | 0.014 | 0.00612 | 0.147 | -2.741407 | -0.4018099 | 354.16 | MGMT | O-6-methylguanine-DNA methyltransferase |
| 5740 | 0.0142 | 0.00624 | 0.117 | 2.735039 | 0.3190407 | 310.26 | PTGIS | prostaglandin I2 synthase |
| 7296 | 0.0147 | 0.00648 | 0.046 | 2.722661 | 0.1241743 | 12256.64 | TXNRD1 | thioredoxin reductase 1 |
| 23491 | 0.018 | 0.00805 | 0.242 | 2.649829 | 0.6408472 | 22.04 | CES3 | carboxylesterase 3 |
| 113189 | 0.0188 | 0.0085 | 0.254 | -2.63142 | -0.6671356 | 636.64 | CHST14 | carbohydrate sulfotransferase 14 |
| 158 | 0.0219 | 0.01 | 0.037 | -2.574595 | -0.0951736 | 2529.09 | ADSL | adenylosuccinate lyase |
| 1244 | 0.0222 | 0.0102 | 0.177 | -2.570328 | -0.4553121 | 39.24 | ABCC2 | ATP binding cassette subfamily C member 2 |
| 2944 | 0.0271 | 0.0127 | 0.293 | -2.4915 | -0.7307277 | 131.49 | GSTM1 | glutathione S-transferase mu 1 |
| 1312 | 0.0274 | 0.0129 | 0.159 | 2.486224 | 0.3956986 | 1261.89 | COMT | catechol-O-methyltransferase |
| 4035 | 0.0305 | 0.0145 | 0.254 | -2.443865 | -0.6211603 | 15405.72 | LRP1 | LDL receptor related protein 1 |
| 2309 | 0.0348 | 0.0168 | 0.042 | -2.391648 | -0.1005123 | 4226.63 | FOXO3 | forkhead box O3 |
| 1594 | 0.0411 | 0.0202 | 0.265 | 2.322133 | 0.6143564 | 12.57 | CYP27B1 | cytochrome P450 family 27 subfamily B member 1 |

### Supplementary Table S15

| | | | | | GSE211993 A79V V CTL | | | |
|---|---|---|---|---|---|---|---|---|
| GeneID | padj | pvalue | lfcSE | stat | log2FoldChange | baseMean | Symbol | Description |
| 6817 | 5E-131 | 2.2E-134 | 0.111 | -24.671184 | -2.7482585 | 142.69 | SULT1A1 | sulfotransferase family 1A member 1 |
| 348 | 2.47E-76 | 4.7E-79 | 0.175 | -18.825151 | -3.2925011 | 2969.34 | APOE | apolipoprotein E |
| 22901 | 6.6E-43 | 3.65E-45 | 0.107 | -14.102796 | -1.5079854 | 805.59 | ARSG | arylsulfatase G |
| 5740 | 5.88E-40 | 3.79E-42 | 0.156 | -13.604066 | -2.1185258 | 1402.92 | PTGIS | prostaglandin I2 synthase |
| 1545 | 2.76E-30 | 2.94E-32 | 0.164 | -11.823694 | -1.9346862 | 13803.35 | CYP1B1 | cytochrome P450 family 1 subfamily B member 1 |
| 1593 | 9.71E-19 | 2.57E-20 | 0.146 | -9.2354336 | -1.3519693 | 215.84 | CYP27A1 | cytochrome P450 family 27 subfamily A member 1 |
| 216 | 1.67E-18 | 4.5E-20 | 0.304 | -9.1753012 | -2.7888816 | 522.24 | ALDH1A1 | aldehyde dehydrogenase 1 family member A1 |
| 2904 | 5.63E-18 | 1.6E-19 | 0.23 | 9.0377843 | 2.0791726 | 9769.21 | GRIN2B | glutamate ionotropic receptor NMDA type subunit 2B |
| 3077 | 1.25E-17 | 3.67E-19 | 0.173 | -8.9464078 | -1.5441431 | 131.72 | HFE | homeostatic iron regulator |
| 7082 | 4.24E-17 | 1.33E-18 | 0.104 | -8.8034762 | -0.917771 | 8075.4 | TJP1 | tight junction protein 1 |
| 1573 | 1.36E-16 | 4.52E-18 | 0.351 | -8.6647546 | -3.0430186 | 29.83 | CYP2J2 | cytochrome P450 family 2 subfamily J member 2 |
| 1312 | 2.35E-16 | 8.04E-18 | 0.102 | -8.5989888 | -0.8797209 | 925.85 | COMT | catechol-O-methyltransferase |
| 2730 | 2.64E-15 | 1E-16 | 0.152 | -8.3043326 | -1.2629548 | 1263.46 | GCLM | glutamate-cysteine ligase modifier subunit |
| 1592 | 4.14E-15 | 1.59E-16 | 0.152 | 8.2494008 | 1.2539562 | 1306.46 | CYP26A1 | cytochrome P450 family 26 subfamily A member 1 |
| 9429 | 3.3E-14 | 1.41E-15 | 0.295 | -7.9841558 | -2.353674 | 100.29 | ABCG2 | ATP binding cassette subfamily G member 2 (Junior blood group) |
| 4301 | 3.81E-13 | 1.92E-14 | 0.063 | 7.6560256 | 0.4845353 | 13055.97 | AFDN | afadin, adherens junction formation factor |
| 7363 | 1.36E-11 | 8.44E-13 | 0.302 | 7.153807 | 2.1633192 | 31.01 | UGT2B4 | UDP glucuronosyltransferase family 2 member B4 |
| 1646 | 1.81E-11 | 1.14E-12 | 0.206 | 7.1121717 | 1.4660299 | 1060.32 | AKR1C2 | aldo-keto reductase family 1 member C2 |
| 412 | 2.88E-11 | 1.87E-12 | 0.17 | 7.0438785 | 1.195667 | 1857.58 | STS | steroid sulfatase |
| 2053 | 5.37E-11 | 3.62E-12 | 0.103 | -6.9513773 | -0.7187282 | 600.11 | EPHX2 | epoxide hydrolase 2 |
| 4255 | 6.57E-11 | 4.46E-12 | 0.178 | -6.9218958 | -1.2336 | 264.56 | MGMT | O-6-methylguanine-DNA methyltransferase |
| 316 | 4.14E-10 | 3.19E-11 | 0.499 | -6.6375252 | -3.3104039 | 264.74 | AOX1 | aldehyde oxidase 1 |
| 114112 | 6.62E-10 | 5.27E-11 | 0.137 | -6.5632095 | -0.8996947 | 624.38 | TXNRD3 | thioredoxin reductase 3 |
| 5649 | 2.23E-09 | 1.96E-10 | 0.203 | 6.3647611 | 1.2941334 | 10685.62 | RELN | reelin |
| 2729 | 6E-09 | 5.66E-10 | 0.093 | -6.1997172 | -0.5769368 | 1951.24 | GCLC | glutamate-cysteine ligase catalytic subunit |
| 788 | 1.05E-08 | 1.03E-09 | 0.147 | -6.1052382 | -0.8949178 | 502.64 | SLC25A20 | solute carrier family 25 member 20 |
| 6799 | 2.41E-08 | 2.52E-09 | 0.289 | -5.9603413 | -1.7218468 | 26.62 | SULT1A2 | sulfotransferase family 1A member 2 |
| 120227 | 3.93E-08 | 4.27E-09 | 0.102 | 5.8732838 | 0.5976039 | 1401.05 | CYP2R1 | cytochrome P450 family 2 subfamily R member 1 |
| 285440 | 1.33E-07 | 1.6E-08 | 0.12 | -5.6506736 | -0.6801833 | 460.31 | CYP4V2 | cytochrome P450 family 4 subfamily V member 2 |
| 147111 | 2.9E-07 | 3.72E-08 | 0.473 | -5.5035157 | -2.6029263 | 26.23 | NOTUM | notum, palmitoleoyl-protein carboxylesterase |
| 627 | 6.33E-07 | 8.66E-08 | 0.173 | 5.3528386 | 0.9231186 | 1855.74 | BDNF | brain derived neurotrophic factor |
| 1244 | 9.15E-07 | 1.29E-07 | 0.218 | -5.2804272 | -1.1520286 | 119.51 | ABCC2 | ATP binding cassette subfamily C member 2 |
| 1543 | 1.29E-06 | 1.86E-07 | 0.361 | 5.2125081 | 1.8824968 | 51.57 | CYP1A1 | cytochrome P450 family 1 subfamily A member 1 |
| 9414 | 2.94E-06 | 4.63E-07 | 0.144 | -5.0408592 | -0.7272926 | 969.92 | TJP2 | tight junction protein 2 |
| 10858 | 3.03E-06 | 4.79E-07 | 0.277 | 5.0345446 | 1.396271 | 249.97 | CYP46A1 | cytochrome P450 family 46 subfamily A member 1 |
| 2950 | 5.5E-06 | 9.15E-07 | 0.161 | -4.9090142 | -0.7893144 | 4562.59 | GSTP1 | glutathione S-transferase pi 1 |
| 1571 | 6.87E-06 | 1.17E-06 | 0.195 | -4.8614687 | -0.9490303 | 82.93 | CYP2E1 | cytochrome P450 family 2 subfamily E member 1 |
| 339761 | 7.14E-06 | 1.22E-06 | 0.237 | 4.8531755 | 1.1510904 | 207.32 | CYP27C1 | cytochrome P450 family 27 subfamily C member 1 |
| 25830 | 7.38E-06 | 1.26E-06 | 0.19 | 4.8460713 | 0.9188099 | 1308.73 | SULT4A1 | sulfotransferase family 4A member 1 |
| 324 | 2.04E-05 | 3.79E-06 | 0.159 | 4.6227374 | 0.7340259 | 22455.53 | APC | APC regulator of WNT signaling pathway |
| 314 | 2.37E-05 | 4.44E-06 | 0.306 | 4.589453 | 1.4042635 | 163.25 | AOC2 | amine oxidase copper containing 2 |
| 10 | 2.93E-05 | 5.61E-06 | 0.508 | -4.540515 | -2.3079578 | 13.53 | NAT2 | N-acetyltransferase 2 |
| 8701 | 0.000032 | 6.18E-06 | 0.201 | -4.5201347 | -0.9072253 | 353.34 | DNAH11 | dynein axonemal heavy chain 11 |
| 57626 | 0.000047 | 9.4E-06 | 0.207 | 4.4304194 | 0.9171625 | 1467.65 | KLHL1 | kelch like family member 1 |
| 1579 | 5.62E-05 | 1.15E-05 | 0.635 | 4.3875043 | 2.7861771 | 14.71 | CYP4A11 | cytochrome P450 family 4 subfamily A member 11 |
| 6532 | 9.82E-05 | 2.14E-05 | 0.333 | -4.2502787 | -1.4165116 | 85.69 | SLC6A4 | solute carrier family 6 member 4 |
| 57576 | 0.000135 | 3.03E-05 | 0.207 | 4.1715638 | 0.8621881 | 95.83 | KIF17 | kinesin family member 17 |
| 23563 | 0.000318 | 7.82E-05 | 0.201 | 3.949944 | 0.7922231 | 45.07 | CHST5 | carbohydrate sulfotransferase 5 |
| 11182 | 0.000402 | 0.000101 | 0.137 | 3.8871814 | 0.533226 | 445.99 | SLC2A6 | solute carrier family 2 member 6 |
| 9446 | 0.000408 | 0.000103 | 0.17 | -3.883018 | -0.6589894 | 956.6 | GSTO1 | glutathione S-transferase omega 1 |
| 51302 | 0.000473 | 0.000121 | 0.225 | -3.8439623 | -0.8659093 | 356.19 | CYP39A1 | cytochrome P450 family 39 subfamily A member 1 |
| 56603 | 0.000511 | 0.000132 | 0.343 | 3.8224254 | 1.3119748 | 307.25 | CYP26B1 | cytochrome P450 family 26 subfamily B member 1 |
| 4502 | 0.00116 | 0.000328 | 0.457 | -3.5917222 | -1.6397913 | 201.81 | MT2A | metallothionein 2A |
| 7296 | 0.00122 | 0.000348 | 0.108 | -3.5763897 | -0.3873064 | 6804.29 | TXNRD1 | thioredoxin reductase 1 |
| 6948 | 0.00138 | 0.000402 | 0.162 | -3.5389043 | -0.5743405 | 1555.96 | TCN2 | transcobalamin 2 |
| 4851 | 0.00182 | 0.000543 | 0.106 | 3.4586567 | 0.3678885 | 2608.96 | NOTCH1 | notch receptor 1 |
| 2903 | 0.00186 | 0.000557 | 0.153 | 3.451632 | 0.5264726 | 2322.63 | GRIN2A | glutamate ionotropic receptor NMDA type subunit 2A |
| 2879 | 0.00268 | 0.000838 | 0.14 | -3.3398149 | -0.4670295 | 3274.94 | GPX4 | glutathione peroxidase 4 |
| 9420 | 0.00279 | 0.000877 | 0.131 | -3.3273787 | -0.4356287 | 976.47 | CYP7B1 | cytochrome P450 family 7 subfamily B member 1 |
| 6819 | 0.00355 | 0.00115 | 0.477 | -3.2499709 | -1.5504121 | 9.64 | SULT1C2 | sulfotransferase family 1C member 2 |
| 1591 | 0.00387 | 0.00127 | 0.357 | 3.2227452 | 1.1504054 | 24.56 | CYP24A1 | cytochrome P450 family 24 subfamily A member 1 |
| 2990 | 0.00485 | 0.00163 | 0.173 | -3.1497039 | -0.5457124 | 1033.47 | GUSB | glucuronidase beta |
| 6999 | 0.00564 | 0.00194 | 0.302 | 3.099526 | 0.9364024 | 39.16 | TDO2 | tryptophan 2,3-dioxygenase |
| 1789 | 0.0069 | 0.00244 | 0.109 | 3.0305368 | 0.3304045 | 640.75 | DNMT3B | DNA methyltransferase 3 beta |
| 210 | 0.00744 | 0.00266 | 0.247 | 3.0050488 | 0.7416588 | 1277.05 | ALAD | aminolevulinate dehydratase |
| 80173 | 0.00903 | 0.00331 | 0.115 | -2.937041 | -0.3390114 | 1796.82 | IFT74 | intraflagellar transport 74 |
| 9536 | 0.0132 | 0.00509 | 0.306 | -2.8012066 | -0.8572951 | 87.22 | PTGES | prostaglandin E synthase |
| 8140 | 0.0136 | 0.00526 | 0.26 | 2.7904957 | 0.7265332 | 4228.21 | SLC7A5 | solute carrier family 7 member 5 |
| 57404 | 0.0138 | 0.00538 | 0.257 | -2.783124 | -0.7158039 | 1877.9 | CYP20A1 | cytochrome P450 family 20 subfamily A member 1 |
| 57530 | 0.015 | 0.00588 | 0.135 | 2.7545359 | 0.3707124 | 463.79 | CGN | cingulin |
| 253152 | 0.027 | 0.0116 | 0.205 | 2.5251049 | 0.5171749 | 108.85 | EPHX4 | epoxide hydrolase 4 |
| 2876 | 0.0324 | 0.0142 | 0.202 | -2.4509619 | -0.4938413 | 1426.19 | GPX1 | glutathione peroxidase 1 |
| 5621 | 0.034 | 0.015 | 0.137 | -2.4316239 | -0.3324151 | 12994.77 | PRNP | prion protein |
| 56171 | 0.0434 | 0.0199 | 0.265 | -2.3277508 | -0.6169531 | 1906.12 | DNAH7 | dynein axonemal heavy chain 7 |
| 6584 | 0.0468 | 0.0218 | 0.159 | -2.2940934 | -0.3652063 | 407.69 | SLC22A5 | solute carrier family 22 member 5 |
| 4363 | 0.0487 | 0.0228 | 0.239 | 2.276426 | 0.5431649 | 1831.21 | ABCC1 | ATP binding cassette subfamily C member 1 |

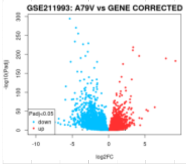
GSE211993: A79V vs GENE CORRECTED
-log10(Padj)
log2FC
Padj<0.05
down
up

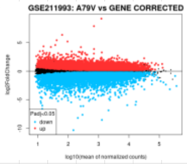
GSE211993: A79V vs GENE CORRECTED
log2FoldChange
log10(mean of normalized counts)
Padj<0.05
down
up

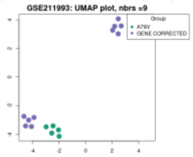
GSE211993: UMAP plot, nbrs =9
Group
A79V
GENE CORRECTED

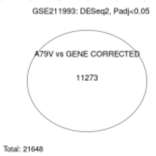
GSE211993: DESeq2, Padj<0.05
A79V vs GENE CORRECTED
11273
Total: 21648

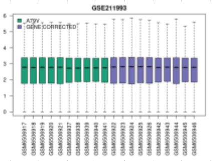
GSE211993
A79V
GENE CORRECTED
log10(normalized counts)

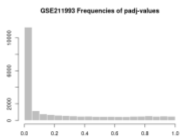
GSE211993 Frequencies of padj-values

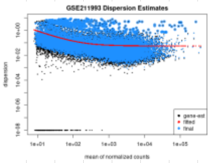
GSE211993 Dispersion Estimates
dispersion
mean of normalized counts
gene-est
fitted
final

Supplementary Table S16

| | | | | GSE211993 A79V/L150P V CTL | | | | |
|---|---|---|---|---|---|---|---|---|
| GeneID | padj | pvalue | lfcSE | stat | log2FoldChange | baseMean | Symbol | Description |
| 7082 | 1.9E-85 | 8.4E-89 | 0.046 | -19.97903 | -0.921389 | 8583.01 | TJP1 | tight junction protein 1 |
| 9429 | 2.4E-26 | 3E-28 | 0.178 | -11.02262 | -1.9578409 | 90.97 | ABCG2 | ATP binding cassette subfamily G member 2 (Junior blood group) |
| 5649 | 4.5E-19 | 1.2E-20 | 0.19 | 9.3145 | 1.7650471 | 13847.96 | RELN | reelin |
| 4502 | 3.5E-18 | 1E-19 | 0.226 | -9.085889 | -2.0508693 | 249.8 | MT2A | metallothionein 2A |
| 7363 | 3.2E-15 | 1.3E-16 | 0.309 | 8.271153 | 2.5519473 | 17.37 | UGT2B4 | UDP glucuronosyltransferase family 2 member B4 |
| 22901 | 5.8E-13 | 3.1E-14 | 0.117 | -7.593939 | -0.8911702 | 829.03 | ARSG | arylsulfatase G |
| 57626 | 1.9E-11 | 1.2E-12 | 0.216 | 7.106239 | 1.532265 | 957.67 | KLHL1 | kelch like family member 1 |
| 6817 | 1E-10 | 7E-12 | 0.266 | -6.857888 | -1.8245032 | 129.68 | SULT1A1 | sulfotransferase family 1A member 1 |
| 627 | 1.4E-09 | 1.2E-10 | 0.184 | 6.443376 | 1.1822142 | 1096.12 | BDNF | brain derived neurotrophic factor |
| 4301 | 1.6E-09 | 1.3E-10 | 0.077 | 6.426124 | 0.4927607 | 10127.05 | AFDN | afadin, adherens junction formation factor |
| 788 | 2.9E-09 | 2.5E-10 | 0.142 | -6.325328 | -0.899489 | 695.57 | SLC25A20 | solute carrier family 25 member 20 |
| 6948 | 3.5E-09 | 3.1E-10 | 0.111 | -6.294915 | -0.6974697 | 1671.25 | TCN2 | transcobalamin 2 |
| 316 | 1.1E-08 | 1E-09 | 0.439 | -6.105437 | -2.6779674 | 179.26 | AOX1 | aldehyde oxidase 1 |
| 58494 | 1.9E-08 | 1.9E-09 | 0.164 | -6.003896 | -0.9832596 | 6848.91 | JAM2 | junctional adhesion molecule 2 |
| 25830 | 2.8E-08 | 2.9E-09 | 0.124 | 5.937416 | 0.7371242 | 1020.97 | SULT4A1 | sulfotransferase family 4A member 1 |
| 1812 | 1.6E-07 | 1.8E-08 | 0.278 | 5.629647 | 1.5659715 | 97.12 | DRD1 | dopamine receptor D1 |
| 5740 | 1.6E-06 | 2.2E-07 | 0.341 | -5.182275 | -1.7688043 | 1096.25 | PTGIS | prostaglandin I2 synthase |
| 9446 | 2.5E-06 | 3.6E-07 | 0.118 | -5.088482 | -0.5996656 | 1203 | GSTO1 | glutathione S-transferase omega 1 |
| 2937 | 5.8E-06 | 8.9E-07 | 0.058 | -4.915634 | -0.2863087 | 1752.65 | GSS | glutathione synthetase |
| 2729 | 7E-06 | 1.1E-06 | 0.087 | -4.874112 | -0.4242911 | 2195.01 | GCLC | glutamate-cysteine ligase catalytic subunit |
| 6783 | 1.3E-05 | 2.1E-06 | 0.395 | 4.740086 | 1.8699498 | 22.73 | SULT1E1 | sulfotransferase family 1E member 1 |
| 5621 | 3.4E-05 | 6E-06 | 0.15 | -4.525627 | -0.6787627 | 17378.25 | PRNP | prion protein |
| 1545 | 3.7E-05 | 6.6E-06 | 0.368 | -4.505953 | -1.6599753 | 8728.33 | CYP1B1 | cytochrome P450 family 1 subfamily B member 1 |
| 4255 | 4E-05 | 7.4E-06 | 0.154 | -4.483028 | -0.6923465 | 332.26 | MGMT | O-6-methylguanine-DNA methyltransferase |
| 260293 | 5.1E-05 | 9.5E-06 | 0.223 | 4.428408 | 0.9886011 | 367.72 | CYP4X1 | cytochrome P450 family 4 subfamily X member 1 |
| 113189 | 5.2E-05 | 9.7E-06 | 0.176 | -4.423968 | -0.778756 | 649.06 | CHST14 | carbohydrate sulfotransferase 14 |
| 216 | 7.4E-05 | 1.4E-05 | 0.507 | -4.341593 | -2.2015579 | 334.96 | ALDH1A1 | aldehyde dehydrogenase 1 family member A1 |
| 56603 | 7.4E-05 | 1.4E-05 | 0.366 | 4.338804 | 1.5874957 | 290.17 | CYP26B1 | cytochrome P450 family 26 subfamily B member 1 |
| 114112 | 7.9E-05 | 1.5E-05 | 0.1 | -4.324669 | -0.432178 | 542.69 | TXNRD3 | thioredoxin reductase 3 |
| 348 | 9.1E-05 | 1.8E-05 | 0.399 | -4.288001 | -1.7088094 | 2505.8 | APOE | apolipoprotein E |
| 4854 | 0.00011 | 2.2E-05 | 0.242 | -4.243624 | -1.0253383 | 3655.23 | NOTCH3 | notch receptor 3 |
| 246 | 0.00014 | 2.9E-05 | 0.454 | 4.184496 | 1.8985583 | 19.57 | ALOX15 | arachidonate 15-lipoxygenase |
| 6999 | 0.00015 | 3.1E-05 | 0.209 | 4.164551 | 0.8711255 | 44.64 | TDO2 | tryptophan 2,3-dioxygenase |
| 9536 | 0.00016 | 3.3E-05 | 0.159 | -4.148733 | -0.6601884 | 90.39 | PTGES | prostaglandin E synthase |
| 10 | 0.0002 | 4.2E-05 | 0.29 | -4.096392 | -1.1875039 | 14.53 | NAT2 | N-acetyltransferase 2 |
| 2944 | 0.00023 | 4.9E-05 | 0.213 | -4.061497 | -0.8631087 | 127.1 | GSTM1 | glutathione S-transferase mu 1 |
| 120227 | 0.00032 | 7.1E-05 | 0.146 | 3.972402 | 0.5811961 | 921.31 | CYP2R1 | cytochrome P450 family 2 subfamily R member 1 |
| 1589 | 0.00035 | 7.9E-05 | 0.266 | -3.948874 | -1.048481 | 37.37 | CYP21A2 | cytochrome P450 family 21 subfamily A member 2 |
| 4035 | 0.00039 | 8.8E-05 | 0.169 | -3.920697 | -0.6607692 | 20029.34 | LRP1 | LDL receptor related protein 1 |
| 2730 | 0.0005 | 0.00012 | 0.172 | -3.850953 | -0.6632808 | 1747.48 | GCLM | glutamate-cysteine ligase modifier subunit |
| 1244 | 0.00068 | 0.00017 | 0.254 | -3.76688 | -0.9578865 | 84.66 | ABCC2 | ATP binding cassette subfamily C member 2 |
| 5742 | 0.00076 | 0.00019 | 0.408 | 3.735376 | 1.5256059 | 59.18 | PTGS1 | prostaglandin-endoperoxide synthase 1 |
| 5243 | 0.00115 | 0.0003 | 0.487 | -3.61558 | -1.760461 | 329.61 | ABCB1 | ATP binding cassette subfamily B member 1 |
| 2990 | 0.00121 | 0.00032 | 0.165 | -3.601414 | -0.5947851 | 958.66 | GUSB | glucuronidase beta |
| 339761 | 0.00149 | 0.0004 | 0.266 | 3.542946 | 0.9411953 | 125.8 | CYP27C1 | cytochrome P450 family 27 subfamily C member 1 |
| 221 | 0.00247 | 0.0007 | 0.216 | 3.390682 | 0.7310989 | 186.92 | ALDH3B1 | aldehyde dehydrogenase 3 family member B1 |
| 1593 | 0.00308 | 0.0009 | 0.212 | -3.321054 | -0.7031065 | 204.1 | CYP27A1 | cytochrome P450 family 27 subfamily A member 1 |
| 7498 | 0.00372 | 0.00111 | 0.355 | 3.261966 | 1.1572581 | 90.25 | XDH | xanthine dehydrogenase |
| 1543 | 0.00487 | 0.00151 | 0.251 | 3.173524 | 0.7973119 | 35.06 | CYP1A1 | cytochrome P450 family 1 subfamily A member 1 |
| 4851 | 0.00523 | 0.00163 | 0.203 | -3.149904 | -0.6400153 | 3610.72 | NOTCH1 | notch receptor 1 |
| 7450 | 0.0064 | 0.00206 | 0.29 | -3.081853 | -0.892624 | 11.74 | VWF | von Willebrand factor |
| 2876 | 0.00735 | 0.0024 | 0.198 | -3.035438 | -0.6000501 | 2179.4 | GPX1 | glutathione peroxidase 1 |
| 1646 | 0.00814 | 0.0027 | 0.232 | 3.000357 | 0.6959945 | 1384.82 | AKR1C2 | aldo-keto reductase family 1 member C2 |
| 6532 | 0.00853 | 0.00284 | 0.441 | -2.984387 | -1.3151605 | 57.57 | SLC6A4 | solute carrier family 6 member 4 |
| 2904 | 0.00918 | 0.00309 | 0.242 | 2.958906 | 0.7166219 | 10208.26 | GRIN2B | glutamate ionotropic receptor NMDA type subunit 2B |
| 1573 | 0.0108 | 0.00371 | 0.316 | -2.901791 | -0.9168865 | 40.96 | CYP2J2 | cytochrome P450 family 2 subfamily J member 2 |
| 210 | 0.0159 | 0.00579 | 0.195 | -2.759523 | -0.5373558 | 1517.91 | ALAD | aminolevulinate dehydratase |
| 285440 | 0.0187 | 0.007 | 0.111 | -2.696716 | -0.2987435 | 514.56 | CYP4V2 | cytochrome P450 family 4 subfamily V member 2 |
| 6799 | 0.0233 | 0.00905 | 0.354 | -2.610223 | -0.9228113 | 18.61 | SULT1A2 | sulfotransferase family 1A member 2 |
| 2948 | 0.0234 | 0.00909 | 0.285 | -2.608684 | -0.7439555 | 326.92 | GSTM4 | glutathione S-transferase mu 4 |
| 1558 | 0.0314 | 0.0128 | 0.345 | 2.490556 | 0.8579135 | 53.67 | CYP2C8 | cytochrome P450 family 2 subfamily C member 8 |
| 1592 | 0.0362 | 0.015 | 0.583 | 2.431381 | 1.4176568 | 585.67 | CYP26A1 | cytochrome P450 family 26 subfamily A member 1 |
| 224 | 0.0431 | 0.0184 | 0.168 | -2.356526 | -0.3947508 | 5068.35 | ALDH3A2 | aldehyde dehydrogenase 3 family member A2 |
| 2309 | 0.0447 | 0.0193 | 0.058 | -2.340329 | -0.1346748 | 4325.02 | FOXO3 | forkhead box O3 |
| 2950 | 0.049 | 0.0215 | 0.156 | -2.299286 | -0.357977 | 5942.56 | GSTP1 | glutathione S-transferase pi 1 |

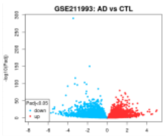

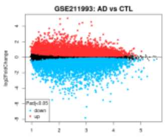

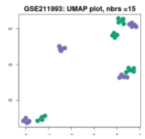

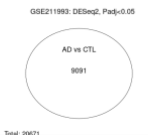

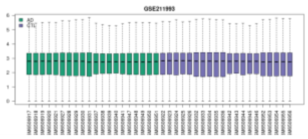

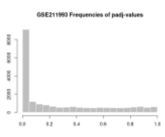

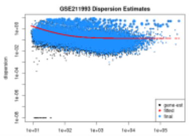

### Supplementary Table S17

| | | | | | GSE227221 4H V CTL | | | |
|---|---|---|---|---|---|---|---|---|
| GeneID | padj | pvalue | lfcSE | stat | log2FoldChange | baseMean | Symbol | Description |
| 11182 | 5E-278 | 5.5E-281 | 0.117 | 35.81964 | 4.1879377 | 4509.23 | SLC2A6 | solute carrier family 2 member 6 |
| 2730 | 1.2E-29 | 4.57E-31 | 0.112 | 11.591175 | 1.2923482 | 872.59 | GCLM | glutamate-cysteine ligase modifier subunit |
| 9414 | 3.4E-27 | 1.4E-28 | 0.169 | 11.090098 | 1.8709643 | 200.81 | TJP2 | tight junction protein 2 |
| 4854 | 8.4E-23 | 4.18E-24 | 0.283 | 10.127366 | 2.8605337 | 71.39 | NOTCH3 | notch receptor 3 |
| 7296 | 2.2E-22 | 1.1E-23 | 0.098 | 10.0325 | 0.9827159 | 4879.29 | TXNRD1 | thioredoxin reductase 1 |
| 1371 | 3.1E-18 | 1.89E-19 | 0.113 | -9.0192357 | -1.0172584 | 496.1 | CPOX | coproporphyrinogen oxidase |
| 2729 | 7.4E-17 | 5.07E-18 | 0.079 | 8.6518028 | 0.6852346 | 7972.97 | GCLC | glutamate-cysteine ligase catalytic subunit |
| 1593 | 1.7E-16 | 1.16E-17 | 0.113 | 8.5571216 | 0.9647953 | 2053.26 | CYP27A1 | cytochrome P450 family 27 subfamily A member 1 |
| 4301 | 9.2E-16 | 6.67E-17 | 0.102 | 8.3526477 | 0.8519864 | 1102.29 | AFDN | afadin, adherens junction formation factor |
| 224 | 1.4E-15 | 9.95E-17 | 0.107 | -8.305357 | -0.8872023 | 558.78 | ALDH3A2 | aldehyde dehydrogenase 3 family member A2 |
| 210 | 3E-13 | 2.68E-14 | 0.113 | -7.6129076 | -0.8573301 | 499.66 | ALAD | aminolevulinate dehydratase |
| 3077 | 1.9E-12 | 1.77E-13 | 0.127 | -7.3652579 | -0.9338363 | 623.96 | HFE | homeostatic iron regulator |
| 8140 | 2.2E-11 | 2.27E-12 | 0.132 | 7.0170102 | 0.9287049 | 916.53 | SLC7A5 | solute carrier family 7 member 5 |
| 4363 | 1.8E-10 | 1.98E-11 | 0.117 | 6.707325 | 0.7863648 | 928.48 | ABCC1 | ATP binding cassette subfamily C member 1 |
| 1595 | 2.4E-08 | 3.41E-09 | 0.088 | 5.9106096 | 0.5211586 | 3327.66 | CYP51A1 | cytochrome P450 family 51 subfamily A member 1 |
| 4035 | 6.4E-08 | 9.44E-09 | 0.148 | -5.7405037 | -0.8481518 | 6272.68 | LRP1 | LDL receptor related protein 1 |
| 285440 | 9.9E-08 | 1.5E-08 | 0.102 | -5.6621274 | -0.5797416 | 642.65 | CYP4V2 | cytochrome P450 family 4 subfamily V member 2 |
| 4502 | 2.5E-07 | 3.94E-08 | 0.265 | 5.4933541 | 1.4538421 | 56.57 | MT2A | metallothionein 2A |
| 113189 | 2.5E-07 | 3.98E-08 | 0.205 | -5.4918357 | -1.1255454 | 103.66 | CHST14 | carbohydrate sulfotransferase 14 |
| 2309 | 4E-07 | 6.52E-08 | 0.112 | -5.4039585 | -0.604254 | 583.63 | FOXO3 | forkhead box O3 |
| 7421 | 6.7E-07 | 1.11E-07 | 0.141 | 5.3069216 | 0.7501564 | 252.21 | VDR | vitamin D receptor |
| 221 | 1.2E-06 | 1.96E-07 | 0.092 | -5.2033944 | -0.4768808 | 1160.29 | ALDH3B1 | aldehyde dehydrogenase 3 family member B1 |
| 80173 | 0.00043 | 0.000112 | 0.158 | -3.8632545 | -0.6111673 | 161.11 | IFT74 | intraflagellar transport 74 |
| 445329 | 0.00068 | 0.000187 | 0.173 | -3.7355751 | -0.645317 | 147.28 | SULT1A4 | sulfotransferase family 1A member 4 |
| 114112 | 0.00073 | 0.000202 | 0.204 | -3.7170832 | -0.7596497 | 91.84 | TXNRD3 | thioredoxin reductase 3 |
| 6818 | 0.00085 | 0.000239 | 0.174 | -3.673347 | -0.6378938 | 148.53 | SULT1A3 | sulfotransferase family 1A member 3 |
| 8701 | 0.00099 | 0.000282 | 0.195 | -3.631323 | -0.7086406 | 101.06 | DNAH11 | dynein axonemal heavy chain 11 |
| 55501 | 0.00131 | 0.000385 | 0.162 | -3.5503354 | -0.5736247 | 174.98 | CHST12 | carbohydrate sulfotransferase 12 |
| 3292 | 0.002 | 0.000613 | 0.183 | -3.4256792 | -0.6261542 | 122.19 | HSD17B1 | hydroxysteroid 17-beta dehydrogenase 1 |
| 6817 | 0.00205 | 0.000631 | 0.089 | -3.417756 | -0.3036163 | 1377.93 | SULT1A1 | sulfotransferase family 1A member 1 |
| 57404 | 0.00438 | 0.00145 | 0.113 | 3.1841766 | 0.3602158 | 554.87 | CYP20A1 | cytochrome P450 family 20 subfamily A member 1 |
| 5742 | 0.00446 | 0.00148 | 0.109 | -3.1778591 | -0.3470034 | 1191.65 | PTGS1 | prostaglandin-endoperoxide synthase 1 |
| 4851 | 0.00539 | 0.00184 | 0.135 | -3.1144413 | -0.4208789 | 1968.01 | NOTCH1 | notch receptor 1 |
| 1594 | 0.0128 | 0.00488 | 0.594 | 2.814622 | 1.6718515 | 11.91 | CYP27B1 | cytochrome P450 family 27 subfamily B member 1 |
| 6999 | 0.0143 | 0.00555 | 0.297 | 2.7732481 | 0.8245996 | 590.17 | TDO2 | tryptophan 2,3-dioxygenase |
| 1789 | 0.0159 | 0.00629 | 0.217 | 2.7322992 | 0.5923762 | 80.25 | DNMT3B | DNA methyltransferase 3 beta |
| 788 | 0.0174 | 0.00695 | 0.165 | -2.6989946 | -0.445556 | 183.54 | SLC25A20 | solute carrier family 25 member 20 |
| 112869 | 0.0264 | 0.011 | 0.142 | -2.5416303 | -0.3609001 | 275.89 | SGF29 | SAGA complex associated factor 29 |
| 113612 | 0.0283 | 0.012 | 0.308 | -2.5135391 | -0.7730557 | 36.16 | CYP2U1 | cytochrome P450 family 2 subfamily U member 1 |
| 9429 | 0.0336 | 0.0146 | 0.146 | -2.4427785 | -0.3572533 | 297.69 | ABCG2 | ATP binding cassette subfamily G member 2 (Junior bl |
| 6821 | 0.0359 | 0.0157 | 0.097 | -2.4150175 | -0.2349857 | 1224.09 | SUOX | sulfite oxidase |
| 120227 | 0.0364 | 0.016 | 0.121 | 2.409387 | 0.2922995 | 420.81 | CYP2R1 | cytochrome P450 family 2 subfamily R member 1 |
| 23491 | 0.0384 | 0.017 | 0.458 | -2.3876084 | -1.0938755 | 20.23 | CES3 | carboxylesterase 3 |
| 2950 | 0.04 | 0.0178 | 0.087 | 2.3692434 | 0.2070282 | 5864.48 | GSTP1 | glutathione S-transferase pi 1 |
| 548593 | 0.0418 | 0.0187 | 0.1 | -2.3513704 | -0.2348808 | 650.28 | SLX1A | SLX1 homolog A, structure-specific endonuclease subu |
| 100130958 | 0.0444 | 0.0201 | 0.255 | 2.3241975 | 0.5931915 | 56.28 | SYCE1L | synaptonemal complex central element protein 1 like |
| 1581 | 0.0475 | 0.0217 | 0.772 | 2.29486 | 1.770823 | 7.36 | CYP7A1 | cytochrome P450 family 7 subfamily A member 1 |

#### Supplementary Table S18

| | | | | | GSE227221 24H V CTL | | | |
|---|---|---|---|---|---|---|---|---|
| GeneID | padj | pvalue | lfcSE | stat | log2FoldChange | baseMean | Symbol | Description |
| 4502 | 0 | 0 | 0.184 | 54.017878 | 9.9101006 | 14879.79 | MT2A | metallothionein 2A |
| 11182 | 1.6E-249 | 6.3E-252 | 0.113 | 33.901528 | 3.812292 | 3612.65 | SLC2A6 | solute carrier family 2 member 6 |
| 1545 | 1.1E-177 | 9E-180 | 0.097 | 28.589868 | 2.758345 | 6342.48 | CYP1B1 | cytochrome P450 family 1 subfamily B member 1 |
| 7421 | 1.1E-140 | 1.4E-142 | 0.152 | 25.422648 | 3.8510053 | 1484.97 | VDR | vitamin D receptor |
| 2729 | 3.7E-106 | 7.1E-108 | 0.095 | -22.0632 | -2.0987271 | 3860.32 | GCLC | glutamate-cysteine ligase catalytic subunit |
| 1593 | 6.82E-75 | 2.32E-76 | 0.124 | 18.493661 | 2.2941119 | 4206.29 | CYP27A1 | cytochrome P450 family 27 subfamily A member 1 |
| 2876 | 2.89E-72 | 1.05E-73 | 0.108 | -18.16126 | -1.9584531 | 30609.39 | GPX1 | glutathione peroxidase 1 |
| 3833 | 1.17E-65 | 4.79E-67 | 0.143 | -17.29896 | -2.4790712 | 411.89 | KIFC1 | kinesin family member C1 |
| 6817 | 4.28E-65 | 1.78E-66 | 0.109 | -17.22312 | -1.8718282 | 992.08 | SULT1A1 | sulfotransferase family 1A member 1 |
| 4854 | 1.21E-62 | 5.29E-64 | 0.252 | 16.890422 | 4.2472778 | 175.7 | NOTCH3 | notch receptor 3 |
| 2948 | 2.38E-54 | 1.23E-55 | 0.151 | -15.71317 | -2.3702241 | 310.04 | GSTM4 | glutathione S-transferase mu 4 |
| 57576 | 4.39E-47 | 2.7E-48 | 0.21 | -14.60271 | -3.0699856 | 168.7 | KIF17 | kinesin family member 17 |
| 2990 | 5.36E-46 | 3.39E-47 | 0.11 | -14.42916 | -1.5802202 | 1237.82 | GUSB | glucuronidase beta |
| 4255 | 2.43E-41 | 1.75E-42 | 0.174 | -13.66051 | -2.3794048 | 276.15 | MGMT | O-6-methylguanine-DNA methyltransferase |
| 4301 | 1.11E-39 | 8.28E-41 | 0.101 | 13.376643 | 1.3561941 | 1431.53 | AFDN | afadin, adherens junction formation factor |
| 6948 | 3.74E-38 | 2.9E-39 | 0.164 | -13.10971 | -2.1508124 | 217.17 | TCN2 | transcobalamin 2 |
| 9429 | 1.1E-35 | 9.07E-37 | 0.566 | -12.66651 | -7.1674354 | 172.01 | ABCG2 | ATP binding cassette subfamily G member 2 (Junior blood group) |
| 2052 | 1.65E-35 | 1.37E-36 | 0.131 | -12.63423 | -1.6570365 | 446.39 | EPHX1 | epoxide hydrolase 1 |
| 9536 | 5.59E-34 | 4.81E-35 | 0.847 | 12.351003 | 10.4653618 | 688 | PTGES | prostaglandin E synthase |
| 4499 | 1.94E-32 | 1.75E-33 | 1.184 | 12.058288 | 14.2777344 | 1785.88 | MT1M | metallothionein 1M |
| 7296 | 3.27E-24 | 3.97E-25 | 0.103 | 10.355156 | 1.0656464 | 5192.01 | TXNRD1 | thioredoxin reductase 1 |
| 3077 | 2.43E-23 | 3.04E-24 | 0.147 | -10.1584 | -1.4917066 | 568.28 | HFE | homeostatic iron regulator |
| 4363 | 4.32E-22 | 5.7E-23 | 0.123 | 9.868518 | 1.2091906 | 1154.35 | ABCC1 | ATP binding cassette subfamily C member 1 |
| 8701 | 2.68E-21 | 3.65E-22 | 0.278 | -9.680366 | -2.691873 | 74.27 | DNAH11 | dynein axonemal heavy chain 11 |
| 2944 | 3.15E-21 | 4.29E-22 | 0.341 | -9.663926 | -3.2973362 | 57.04 | GSTM1 | glutathione S-transferase mu 1 |
| 114112 | 1.11E-20 | 1.55E-21 | 0.291 | -9.531653 | -2.7695238 | 68.1 | TXNRD3 | thioredoxin reductase 3 |
| 1594 | 1.31E-20 | 1.84E-21 | 0.447 | 9.513629 | 4.2526069 | 57.78 | CYP27B1 | cytochrome P450 family 27 subfamily B member 1 |
| 9446 | 8.38E-19 | 1.27E-19 | 0.146 | 9.063211 | 1.3242041 | 8525.18 | GSTO1 | glutathione S-transferase omega 1 |
| 4489 | 3.96E-18 | 6.13E-19 | 1.189 | 8.889653 | 10.5690028 | 136.69 | MT1A | metallothionein 1A |
| 6799 | 2.36E-16 | 3.98E-17 | 0.193 | -8.413482 | -1.622467 | 122.99 | SULT1A2 | sulfotransferase family 1A member 2 |
| 113189 | 3.21E-15 | 5.76E-16 | 0.222 | -8.094227 | -1.7976125 | 94.05 | CHST14 | carbohydrate sulfotransferase 14 |
| 4524 | 4.11E-15 | 7.41E-16 | 0.105 | -8.063493 | -0.849367 | 1536.38 | MTHFR | methylenetetrahydrofolate reductase |
| 224 | 7.41E-14 | 1.45E-14 | 0.124 | -7.692189 | -0.9497509 | 563.16 | ALDH3A2 | aldehyde dehydrogenase 3 family member A2 |
| 221 | 1.2E-11 | 2.7E-12 | 0.104 | -6.992578 | -0.7238504 | 1109.4 | ALDH3B1 | aldehyde dehydrogenase 3 family member B1 |
| 2309 | 1.61E-11 | 3.64E-12 | 0.108 | 6.950314 | 0.7534157 | 968.22 | FOXO3 | forkhead box O3 |
| 6999 | 3.55E-11 | 8.27E-12 | 0.315 | 6.833808 | 2.1523114 | 1189.3 | TDO2 | tryptophan 2,3-dioxygenase |
| 210 | 9.24E-11 | 2.21E-11 | 0.118 | -6.691534 | -0.7915849 | 520.35 | ALAD | aminolevulinate dehydratase |
| 113612 | 1.36E-10 | 3.28E-11 | 0.497 | -6.63329 | -3.2977115 | 25.88 | CYP2U1 | cytochrome P450 family 2 subfamily U member 1 |
| 80173 | 2.29E-10 | 5.6E-11 | 0.142 | 6.553918 | 0.9288151 | 288.96 | IFT74 | intraflagellar transport 74 |
| 7082 | 3.37E-10 | 8.34E-11 | 0.378 | 6.49439 | 2.4529005 | 39.27 | TJP1 | tight junction protein 1 |
| 5621 | 3.85E-08 | 1.12E-08 | 0.121 | -5.710744 | -0.6912111 | 2296.51 | PRNP | prion protein |
| 120227 | 5.03E-08 | 1.48E-08 | 0.164 | -5.663309 | -0.9271753 | 295.61 | CYP2R1 | cytochrome P450 family 2 subfamily R member 1 |
| 1244 | 1.61E-07 | 4.97E-08 | 0.416 | 5.452365 | 2.2663646 | 29.23 | ABCC2 | ATP binding cassette subfamily C member 2 |
| 1595 | 2.5E-07 | 7.82E-08 | 0.112 | -5.371287 | -0.6032276 | 2320.56 | CYP51A1 | cytochrome P450 family 51 subfamily A member 1 |
| 445329 | 7.54E-07 | 2.48E-07 | 0.187 | -5.159492 | -0.9657342 | 138.84 | SULT1A4 | sulfotransferase family 1A member 4 |
| 6818 | 9.81E-07 | 3.25E-07 | 0.188 | -5.108181 | -0.959391 | 139.93 | SULT1A3 | sulfotransferase family 1A member 3 |
| 3292 | 1.21E-06 | 4.05E-07 | 0.153 | 5.066417 | 0.7760688 | 205.57 | HSD17B1 | hydroxysteroid 17-beta dehydrogenase 1 |
| 23491 | 1.34E-06 | 4.53E-07 | 1.246 | -5.045406 | -6.2842519 | 14.48 | CES3 | carboxylesterase 3 |
| 1588 | 1.44E-06 | 4.88E-07 | 0.676 | 5.0309 | 3.3986475 | 14.99 | CYP19A1 | cytochrome P450 family 19 subfamily A member 1 |
| 348 | 8.63E-06 | 3.17E-06 | 0.127 | 4.659313 | 0.5914497 | 4805.3 | APOE | apolipoprotein E |
| 412 | 1.01E-05 | 3.74E-06 | 0.167 | -4.625476 | -0.7699492 | 175.58 | STS | steroid sulfatase |
| 548593 | 1.22E-05 | 4.56E-06 | 0.112 | -4.584051 | -0.5144099 | 611.73 | SLX1A | SLX1 homolog A, structure-specific endonuclease subunit |
| 79008 | 1.69E-05 | 6.41E-06 | 0.125 | -4.51226 | -0.5648422 | 389.01 | SLX1B | SLX1 homolog B, structure-specific endonuclease subunit |
| 9414 | 2.74E-05 | 1.07E-05 | 0.268 | -4.402796 | -1.1795446 | 63.32 | TJP2 | tight junction protein 2 |
| 8140 | 2.77E-05 | 1.08E-05 | 0.154 | -4.400644 | -0.6765875 | 524.7 | SLC7A5 | solute carrier family 7 member 5 |
| 22901 | 0.000038 | 1.51E-05 | 0.112 | 4.32791 | 0.485114 | 656.5 | ARSG | arylsulfatase G |
| 1789 | 7.46E-05 | 3.07E-05 | 0.298 | -4.168093 | -1.2404 | 46.52 | DNMT3B | DNA methyltransferase 3 beta |
| 2937 | 0.000537 | 0.000248 | 0.097 | -3.664557 | -0.3559245 | 1741.25 | GSS | glutathione synthetase |
| 875 | 0.000686 | 0.000322 | 0.126 | 3.596926 | 0.4515901 | 423.63 | CBS | cystathionine beta-synthase |
| 10587 | 0.000852 | 0.000405 | 0.11 | -3.536621 | -0.3878307 | 686.46 | TXNRD2 | thioredoxin reductase 2 |
| 6819 | 0.00148 | 0.000729 | 0.942 | -3.378473 | -3.1812046 | 6.69 | SULT1C2 | sulfotransferase family 1C member 2 |
| 7450 | 0.0028 | 0.00144 | 0.404 | -3.186808 | -1.2873243 | 22.8 | VWF | von Willebrand factor |
| 788 | 0.00315 | 0.00164 | 0.166 | 3.14945 | 0.5238835 | 264.64 | SLC25A20 | solute carrier family 25 member 20 |
| 540 | 0.00502 | 0.00271 | 0.396 | -2.999008 | -1.188645 | 25.95 | ATP7B | ATPase copper transporting beta |
| 216 | 0.00772 | 0.00431 | 0.636 | -2.854749 | -1.815925 | 10.1 | ALDH1A1 | aldehyde dehydrogenase 1 family member A1 |
| 4851 | 0.00874 | 0.00492 | 0.133 | 2.8121 | 0.3730736 | 2642.16 | NOTCH1 | notch receptor 1 |
| 6566 | 0.0121 | 0.00699 | 0.15 | -2.697498 | -0.4045598 | 474.7 | SLC16A1 | solute carrier family 16 member 1 |
| 653689 | 0.015 | 0.00879 | 0.359 | -2.620026 | -0.9414528 | 28.98 | GSTT2B | glutathione S-transferase theta 2B |
| 314 | 0.0157 | 0.00924 | 0.39 | 2.603093 | 1.0150268 | 24.91 | AOC2 | amine oxidase copper containing 2 |
| 2879 | 0.0223 | 0.0135 | 0.095 | 2.471255 | 0.2341149 | 11838.55 | GPX4 | glutathione peroxidase 4 |
| 55501 | 0.0273 | 0.0168 | 0.142 | 2.391301 | 0.3405356 | 243.49 | CHST12 | carbohydrate sulfotransferase 12 |
| 2953 | 0.0337 | 0.0211 | 0.42 | -2.30556 | -0.9674116 | 20.74 | GSTT2 | glutathione S-transferase theta 2 (gene/pseudogene) |
| 112869 | 0.036 | 0.0227 | 0.16 | -2.27883 | -0.3653986 | 282.19 | SGF29 | SAGA complex associated factor 29 |
| 56171 | 0.042 | 0.0269 | 0.662 | -2.213557 | -1.4642223 | 8.8 | DNAH7 | dynein axonemal heavy chain 7 |
| 1577 | 0.0428 | 0.0274 | 0.25 | 2.205583 | 0.551866 | 66.38 | CYP3A5 | cytochrome P450 family 3 subfamily A member 5 |

Supplementary Table S19

| | | | | | GSE227221 96H V CTL | | | |
|---|---|---|---|---|---|---|---|---|
| GeneID | padj | pvalue | lfcSE | stat | log2FoldChange | baseMean | Symbol | Description |
| 4502 | 3.9E-248 | 5.9E-251 | 0.275 | 33.835901 | 9.309599 | 10151.35 | MT2A | metallothionein 2A |
| 1545 | 1.1E-117 | 8.3E-120 | 0.075 | 23.273067 | 1.754006 | 3680.8 | CYP1B1 | cytochrome P450 family 1 subfamily B member 1 |
| 7421 | 2.2E-108 | 1.8E-110 | 0.121 | 22.331165 | 2.709407 | 748.5 | VDR | vitamin D receptor |
| 11182 | 2.1E-100 | 2.1E-102 | 0.105 | 21.486215 | 2.256905 | 1429.87 | SLC2A6 | solute carrier family 2 member 6 |
| 6948 | 1.42E-73 | 2.56E-75 | 0.208 | -18.36376 | -3.814961 | 195.79 | TCN2 | transcobalamin 2 |
| 57576 | 1.09E-59 | 2.67E-61 | 0.23 | -16.51913 | -3.798503 | 166.73 | KIF17 | kinesin family member 17 |
| 9429 | 1.33E-55 | 3.71E-57 | 0.375 | -15.93351 | -5.976569 | 178.97 | ABCG2 | ATP binding cassette subfamily G member 2 (Junior blo |
| 348 | 1.08E-50 | 3.36E-52 | 0.082 | 15.203255 | 1.239355 | 6639.21 | APOE | apolipoprotein E |
| 2052 | 2.05E-43 | 7.8E-45 | 0.114 | -14.04912 | -1.604339 | 464.57 | EPHX1 | epoxide hydrolase 1 |
| 4301 | 1.22E-41 | 4.86E-43 | 0.087 | 13.753371 | 1.196658 | 1364.85 | AFDN | afadin, adherens junction formation factor |
| 6817 | 7.83E-36 | 3.69E-37 | 0.088 | -12.73692 | -1.119142 | 1172.97 | SULT1A1 | sulfotransferase family 1A member 1 |
| 4499 | 3.03E-32 | 1.66E-33 | 1.203 | 12.06284 | 14.514714 | 2168.95 | MT1M | metallothionein 1M |
| 2729 | 5.5E-31 | 3.15E-32 | 0.077 | -11.81806 | -0.907138 | 4945.66 | GCLC | glutamate-cysteine ligase catalytic subunit |
| 4854 | 9.42E-24 | 7.02E-25 | 0.28 | 10.300379 | 2.879077 | 76.21 | NOTCH3 | notch receptor 3 |
| 2948 | 2.92E-23 | 2.23E-24 | 0.133 | -10.18847 | -1.359582 | 372.79 | GSTM4 | glutathione S-transferase mu 4 |
| 2990 | 2.39E-22 | 1.88E-23 | 0.09 | -9.978937 | -0.898387 | 1468.46 | GUSB | glucuronidase beta |
| 3077 | 7.86E-21 | 6.68E-22 | 0.135 | -9.618458 | -1.301132 | 606.6 | HFE | homeostatic iron regulator |
| 4489 | 5.74E-20 | 5.06E-21 | 1.187 | 9.407943 | 11.17118 | 213.55 | MT1A | metallothionein 1A |
| 9446 | 3.28E-18 | 3.16E-19 | 0.109 | 8.963094 | 0.972678 | 7426.48 | GSTO1 | glutathione S-transferase omega 1 |
| 4524 | 1.27E-16 | 1.34E-17 | 0.084 | -8.540183 | -0.718195 | 1637.78 | MTHFR | methylenetetrahydrofolate reductase |
| 2876 | 1.12E-15 | 1.25E-16 | 0.092 | -8.277861 | -0.757041 | 39932.78 | GPX1 | glutathione peroxidase 1 |
| 221 | 1.77E-15 | 2E-16 | 0.09 | -8.221797 | -0.736312 | 1140.53 | ALDH3B1 | aldehyde dehydrogenase 3 family member B1 |
| 2944 | 1.51E-12 | 2.05E-13 | 0.244 | -7.345361 | -1.790867 | 68.82 | GSTM1 | glutathione S-transferase mu 1 |
| 8701 | 7.29E-11 | 1.13E-11 | 0.207 | -6.788367 | -1.402994 | 91.08 | DNAH11 | dynein axonemal heavy chain 11 |
| 1588 | 2.48E-10 | 4.04E-11 | 0.61 | 6.602561 | 4.030146 | 23.13 | CYP19A1 | cytochrome P450 family 19 subfamily A member 1 |
| 7450 | 8.9E-10 | 1.52E-10 | 0.251 | 6.40382 | 1.609678 | 67.73 | VWF | von Willebrand factor |
| 80173 | 1.45E-09 | 2.53E-10 | 0.131 | 6.324903 | 0.827528 | 284.58 | IFT74 | intraflagellar transport 74 |
| 4255 | 2.57E-09 | 4.58E-10 | 0.132 | -6.232831 | -0.825462 | 373.15 | MGMT | O-6-methylguanine-DNA methyltransferase |
| 29785 | 1.64E-08 | 3.17E-09 | 0.108 | 5.922726 | 0.641513 | 580.39 | CYP2S1 | cytochrome P450 family 2 subfamily S member 1 |
| 875 | 4.92E-08 | 9.93E-09 | 0.107 | 5.731995 | 0.613856 | 466.67 | CBS | cystathionine beta-synthase |
| 1595 | 2.25E-07 | 4.9E-08 | 0.104 | -5.454863 | -0.566297 | 2415.89 | CYP51A1 | cytochrome P450 family 51 subfamily A member 1 |
| 6799 | 2.7E-07 | 5.92E-08 | 0.164 | -5.4213 | -0.888548 | 147.64 | SULT1A2 | sulfotransferase family 1A member 2 |
| 112869 | 3.48E-07 | 7.72E-08 | 0.134 | -5.373595 | -0.720229 | 263.19 | SGF29 | SAGA complex associated factor 29 |
| 4035 | 3.27E-06 | 8.14E-07 | 0.104 | 4.931937 | 0.511623 | 10310.94 | LRP1 | LDL receptor related protein 1 |
| 6566 | 3.39E-06 | 8.45E-07 | 0.127 | -4.924667 | -0.626255 | 458.84 | SLC16A1 | solute carrier family 16 member 1 |
| 1244 | 1.13E-05 | 3.03E-06 | 0.409 | 4.668736 | 1.909021 | 24.57 | ABCC2 | ATP binding cassette subfamily C member 2 |
| 10587 | 1.26E-05 | 3.4E-06 | 0.1 | -4.644889 | -0.462643 | 692.69 | TXNRD2 | thioredoxin reductase 2 |
| 4363 | 3.93E-05 | 1.14E-05 | 0.118 | -4.387931 | -0.518815 | 610.06 | ABCC1 | ATP binding cassette subfamily C member 1 |
| 2937 | 9.82E-05 | 3.04E-05 | 0.08 | -4.170501 | -0.332325 | 1807.77 | GSS | glutathione synthetase |
| 6999 | 0.000146 | 4.67E-05 | 0.305 | 4.071739 | 1.243086 | 757.65 | TDO2 | tryptophan 2,3-dioxygenase |
| 113189 | 0.000256 | 8.49E-05 | 0.178 | -3.93001 | -0.697551 | 121.9 | CHST14 | carbohydrate sulfotransferase 14 |
| 6819 | 0.000291 | 9.74E-05 | 1.317 | -3.897035 | -5.131187 | 6.45 | SULT1C2 | sulfotransferase family 1C member 2 |
| 314 | 0.000326 | 0.00011 | 0.36 | 3.866596 | 1.392327 | 30.57 | AOC2 | amine oxidase copper containing 2 |
| 7082 | 0.00101 | 0.000374 | 0.443 | 3.557684 | 1.577293 | 25.14 | TJP1 | tight junction protein 1 |
| 445329 | 0.00105 | 0.000387 | 0.172 | -3.548688 | -0.609566 | 156.3 | SULT1A4 | sulfotransferase family 1A member 4 |
| 56171 | 0.00111 | 0.000413 | 0.853 | -3.531948 | -3.012216 | 7.54 | DNAH7 | dynein axonemal heavy chain 7 |
| 6818 | 0.00132 | 0.000496 | 0.172 | -3.482819 | -0.600186 | 157.71 | SULT1A3 | sulfotransferase family 1A member 3 |
| 788 | 0.00354 | 0.00145 | 0.143 | 3.185065 | 0.454021 | 264.98 | SLC25A20 | solute carrier family 25 member 20 |
| 3833 | 0.00363 | 0.00149 | 0.111 | -3.176615 | -0.352605 | 642.01 | KIFC1 | kinesin family member C1 |
| 120227 | 0.00638 | 0.00276 | 0.119 | -2.993716 | -0.357279 | 354.88 | CYP2R1 | cytochrome P450 family 2 subfamily R member 1 |
| 23491 | 0.00733 | 0.00321 | 0.433 | -2.946535 | -1.274726 | 21.02 | CES3 | carboxylesterase 3 |
| 57404 | 0.00831 | 0.00369 | 0.108 | -2.903874 | -0.312938 | 461.75 | CYP20A1 | cytochrome P450 family 20 subfamily A member 1 |
| 7296 | 0.00879 | 0.00392 | 0.093 | 2.884703 | 0.268044 | 3814.19 | TXNRD1 | thioredoxin reductase 1 |
| 113612 | 0.00901 | 0.00402 | 0.304 | -2.876205 | -0.874783 | 37.23 | CYP2U1 | cytochrome P450 family 2 subfamily U member 1 |
| 653689 | 0.0157 | 0.00748 | 0.341 | -2.674675 | -0.913056 | 29.93 | GSTT2B | glutathione S-transferase theta 2B |
| 79008 | 0.0161 | 0.00767 | 0.114 | -2.66647 | -0.304457 | 433.16 | SLX1B | SLX1 homolog B, structure-specific endonuclease subu |
| 1371 | 0.0163 | 0.00779 | 0.111 | -2.661174 | -0.29453 | 636.67 | CPOX | coproporphyrinogen oxidase |
| 548593 | 0.0225 | 0.0111 | 0.1 | -2.539663 | -0.254307 | 682.07 | SLX1A | SLX1 homolog A, structure-specific endonuclease subu |
| 3292 | 0.0232 | 0.0115 | 0.155 | 2.526612 | 0.392139 | 181.01 | HSD17B1 | hydroxysteroid 17-beta dehydrogenase 1 |
| 22901 | 0.0283 | 0.0144 | 0.099 | 2.448216 | 0.24274 | 615.85 | ARSG | arylsulfatase G |
| 2953 | 0.0457 | 0.0245 | 0.398 | -2.248689 | -0.894903 | 21.58 | GSTT2 | glutathione S-transferase theta 2 (gene/pseudogene) |
| 84171 | 0.0459 | 0.0247 | 0.704 | 2.246771 | 1.58238 | 7.3 | LOXL4 | lysyl oxidase like 4 |

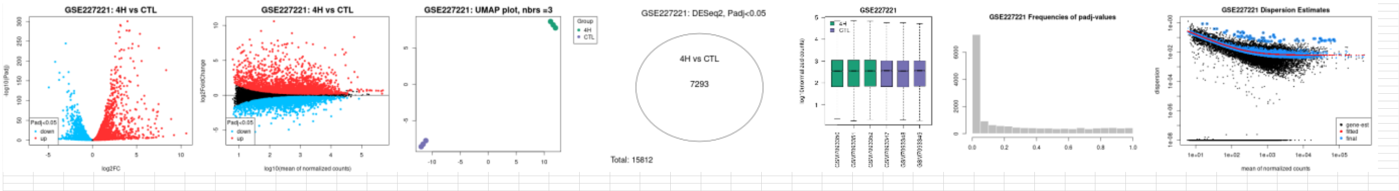
GSE227221: 4H vs CTL
GSE227221: 4H vs CTL
GSE227221: UMAP plot, nbrs =3
GSE227221: DESeq2, Padj<0.05
4H vs CTL
7293
Total: 15812
GSE227221
GSE227221 Frequencies of padj-values
GSE227221 Dispersion Estimates

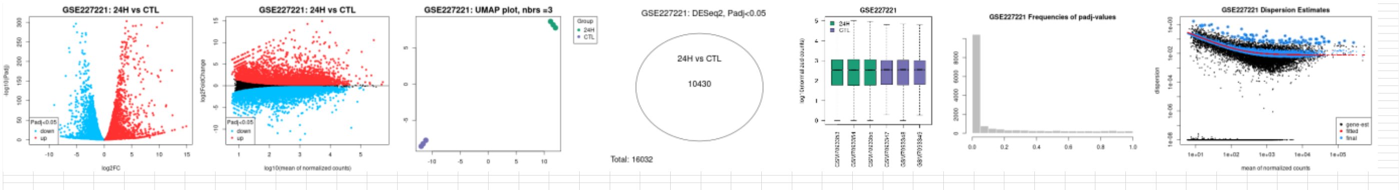
GSE227221: 24H vs CTL
GSE227221: 24H vs CTL
GSE227221: UMAP plot, nbrs =3
GSE227221: DESeq2, Padj<0.05
24H vs CTL
10430
Total: 16032
GSE227221
GSE227221 Frequencies of padj-values
GSE227221 Dispersion Estimates

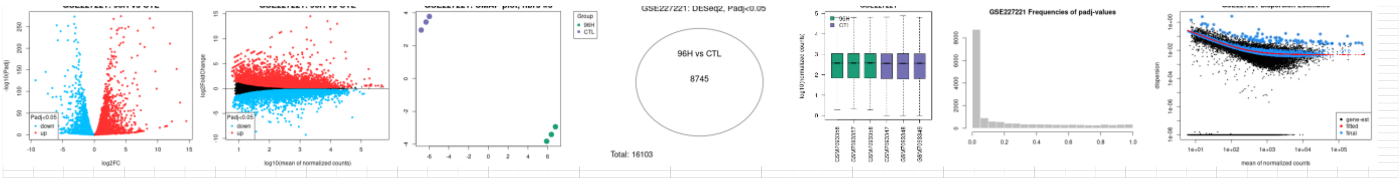
GSE227221: DESeq2, Padj<0.05
96H vs CTL
8745
Total: 16103
GSE227221 Frequencies of padj-values

Supplementary Table S20

| | | | | | GSE190125 T21 V CTL PART 1 | | | |
|---|---|---|---|---|---|---|---|---|
| GeneID | padj | pvalue | lfcSE | stat | log2FoldChange | baseMean | Symbol | Description |
| 2052 | 4.9E-43 | 3.4E-45 | 0.0276 | -14.10807 | -0.3887186 | 804.86 | EPHX1 | epoxide hydrolase 1 |
| 2876 | 8.9E-33 | 1.2E-34 | 0.0916 | 12.275002 | 1.1241043 | 30344.09 | GPX1 | glutathione peroxidase 1 |
| 8140 | 1.7E-25 | 4.4E-27 | 0.062 | 10.776632 | 0.668635 | 929.57 | SLC7A5 | solute carrier family 7 member 5 |
| 55501 | 3.4E-24 | 9.8E-26 | 0.0443 | 10.488358 | 0.4646903 | 2614.63 | CHST12 | carbohydrate sulfotransferase 12 |
| 540 | 3.5E-24 | 1E-25 | 0.0594 | -10.48444 | -0.6223539 | 52.01 | ATP7B | ATPase copper transporting beta |
| 875 | 1.6E-21 | 6.1E-23 | 0.1195 | 9.861759 | 1.1786797 | 154.95 | CBS | cystathionine beta-synthase |
| 253152 | 6.7E-21 | 2.8E-22 | 0.1112 | 9.707329 | 1.0795691 | 30.53 | EPHX4 | epoxide hydrolase 4 |
| 79008 | 7.7E-19 | 4.1E-20 | 0.0362 | 9.186626 | 0.3322111 | 852.93 | SLX1B | SLX1 homolog B, structure-specific endonuclease subu |
| 314 | 2.7E-18 | 1.5E-19 | 0.0597 | 9.045264 | 0.5402363 | 975.15 | AOC2 | amine oxidase copper containing 2 |
| 548593 | 1.4E-17 | 8.2E-19 | 0.0346 | 8.856858 | 0.3062319 | 1531.46 | SLX1A | SLX1 homolog A, structure-specific endonuclease subu |
| 246 | 9E-17 | 5.9E-18 | 0.1411 | -8.63403 | -1.2179822 | 513.99 | ALOX15 | arachidonate 15-lipoxygenase |
| 3077 | 1.5E-16 | 1E-17 | 0.0466 | 8.573156 | 0.3997919 | 378.63 | HFE | homeostatic iron regulator |
| 3833 | 1.5E-16 | 1.1E-17 | 0.0804 | 8.568175 | 0.6886228 | 105.1 | KIFC1 | kinesin family member C1 |
| 9446 | 4.6E-16 | 3.3E-17 | 0.0437 | 8.435342 | 0.3682029 | 2285.73 | GSTO1 | glutathione S-transferase omega 1 |
| 1591 | 5.1E-15 | 4.1E-16 | 0.1465 | -8.135543 | -1.1916493 | 8.37 | CYP24A1 | cytochrome P450 family 24 subfamily A member 1 |
| 5742 | 3.1E-14 | 2.8E-15 | 0.0716 | 7.900615 | 0.5660232 | 2887 | PTGS1 | prostaglandin-endoperoxide synthase 1 |
| 7122 | 4.7E-14 | 4.3E-15 | 0.1005 | 7.844737 | 0.7886671 | 372.44 | CLDN5 | claudin 5 |
| 7421 | 6.1E-14 | 5.7E-15 | 0.0643 | 7.81013 | 0.5018776 | 1222.37 | VDR | vitamin D receptor |
| 1548 | 6.8E-14 | 6.4E-15 | 0.1202 | -7.795111 | -0.9365832 | 7.92 | CYP2A6 | cytochrome P450 family 2 subfamily A member 6 |
| 10720 | 8.7E-14 | 8.4E-15 | 0.1409 | -7.761219 | -1.0936929 | 52.76 | UGT2B11 | UDP glucuronosyltransferase family 2 member B11 |
| 6818 | 2E-13 | 2E-14 | 0.038 | 7.64778 | 0.2904954 | 623.62 | SULT1A3 | sulfotransferase family 1A member 3 |
| 445329 | 3E-13 | 3E-14 | 0.0385 | 7.596528 | 0.2921561 | 614.05 | SULT1A4 | sulfotransferase family 1A member 4 |
| 126410 | 3.2E-13 | 3.3E-14 | 0.0857 | 7.587324 | 0.6504995 | 121.24 | CYP4F22 | cytochrome P450 family 4 subfamily F member 22 |
| 8701 | 3.4E-13 | 3.5E-14 | 0.0701 | -7.578248 | -0.5308801 | 137 | DNAH11 | dynein axonemal heavy chain 11 |
| 6532 | 1.1E-12 | 1.2E-13 | 0.0871 | 7.419304 | 0.6459202 | 35.32 | SLC6A4 | solute carrier family 6 member 4 |
| 4502 | 4.1E-12 | 4.9E-13 | 0.091 | 7.227129 | 0.657702 | 1530.86 | MT2A | metallothionein 2A |
| 4854 | 6.6E-12 | 8.1E-13 | 0.1064 | -7.1593 | -0.7614188 | 10.08 | NOTCH3 | notch receptor 3 |
| 113612 | 4E-11 | 5.4E-12 | 0.0364 | -6.894201 | -0.2510183 | 299.69 | CYP2U1 | cytochrome P450 family 2 subfamily U member 1 |
| 2990 | 4.7E-11 | 6.4E-12 | 0.0334 | 6.870307 | 0.2291551 | 1940.02 | GUSB | glucuronidase beta |
| 23491 | 5.7E-11 | 7.9E-12 | 0.102 | 6.840681 | 0.6978596 | 10.12 | CES3 | carboxylesterase 3 |
| 11182 | 6E-11 | 8.4E-12 | 0.0435 | 6.830848 | 0.2973603 | 1685.48 | SLC2A6 | solute carrier family 2 member 6 |
| 29785 | 1.3E-10 | 1.8E-11 | 0.0758 | 6.718207 | 0.5089096 | 250.23 | CYP2S1 | cytochrome P450 family 2 subfamily S member 1 |
| 120227 | 2.7E-10 | 4.1E-11 | 0.0443 | -6.602173 | -0.2921619 | 683.17 | CYP2R1 | cytochrome P450 family 2 subfamily R member 1 |
| 9429 | 1.1E-09 | 1.8E-10 | 0.1159 | 6.375408 | 0.7390635 | 45.85 | ABCG2 | ATP binding cassette subfamily G member 2 (Junior blo |
| 10587 | 1.7E-09 | 3E-10 | 0.0375 | 6.300449 | 0.2364772 | 1172.51 | TXNRD2 | thioredoxin reductase 2 |
| 1583 | 4.4E-09 | 7.9E-10 | 0.1572 | 6.146274 | 0.9660252 | 10.29 | CYP11A1 | cytochrome P450 family 11 subfamily A member 1 |
| 1545 | 5.5E-09 | 1E-09 | 0.078 | -6.107563 | -0.4764854 | 826.42 | CYP1B1 | cytochrome P450 family 1 subfamily B member 1 |

### Supplementary Table S21

| | | | | | | | | |
|---|---|---|---|---|---|---|---|---|
| | | | | | GSE190125 T21 V CTL PART 2 | | | |
| 2937 | 1.1E-08 | 2.1E-09 | 0.0336 | 5.990253 | 0.2014988 | 900.56 | GSS | glutathione synthetase |
| 348 | 1.5E-08 | 2.9E-09 | 0.0871 | 5.938834 | 0.5172595 | 14.98 | APOE | apolipoprotein E |
| 2879 | 2.1E-08 | 4.2E-09 | 0.0493 | 5.877612 | 0.2895182 | 5685.45 | GPX4 | glutathione peroxidase 4 |
| 66002 | 1E-07 | 2.3E-08 | 0.0893 | -5.584999 | -0.4984824 | 129.61 | CYP4F12 | cytochrome P450 family 4 subfamily F member 12 |
| 57530 | 1.2E-07 | 2.8E-08 | 0.1009 | -5.554018 | -0.5601689 | 9.86 | CGN | cingulin |
| 1589 | 1.2E-06 | 3.3E-07 | 0.0946 | 5.103517 | 0.4827723 | 14.67 | CYP21A2 | cytochrome P450 family 21 subfamily A member 2 |
| 6566 | 1.8E-06 | 5.1E-07 | 0.0338 | -5.02302 | -0.169575 | 314.24 | SLC16A1 | solute carrier family 16 member 1 |
| 57576 | 1.9E-06 | 5.3E-07 | 0.1101 | 5.016307 | 0.5523015 | 11.46 | KIF17 | kinesin family member 17 |
| 324 | 2E-06 | 5.6E-07 | 0.0426 | -5.003029 | -0.2132694 | 1215.78 | APC | APC regulator of WNT signaling pathway |
| 216 | 2.1E-06 | 5.9E-07 | 0.0707 | -4.995055 | -0.3532059 | 892.16 | ALDH1A1 | aldehyde dehydrogenase 1 family member A1 |
| 9536 | 3.8E-06 | 1.1E-06 | 0.1367 | 4.872019 | 0.6659596 | 74.86 | PTGES | prostaglandin E synthase |
| 1244 | 6.1E-06 | 1.9E-06 | 0.0443 | -4.767897 | -0.2112821 | 144.89 | ABCC2 | ATP binding cassette subfamily C member 2 |
| 3292 | 8.6E-06 | 2.7E-06 | 0.0229 | 4.693645 | 0.1074413 | 404.4 | HSD17B1 | hydroxysteroid 17-beta dehydrogenase 1 |
| 6584 | 9.5E-06 | 3E-06 | 0.0394 | -4.671056 | -0.1838515 | 289.28 | SLC22A5 | solute carrier family 22 member 5 |
| 2053 | 3.1E-05 | 1.1E-05 | 0.0633 | -4.401194 | -0.278453 | 909 | EPHX2 | epoxide hydrolase 2 |
| 6821 | 3.6E-05 | 1.3E-05 | 0.0395 | 4.365478 | 0.1722037 | 3330.6 | SUOX | sulfite oxidase |
| 6948 | 0.00005 | 1.8E-05 | 0.1021 | 4.289226 | 0.4378222 | 310.19 | TCN2 | transcobalamin 2 |
| 788 | 5.2E-05 | 1.9E-05 | 0.0447 | 4.282114 | 0.1915563 | 1232.94 | SLC25A20 | solute carrier family 25 member 20 |
| 1594 | 6.2E-05 | 2.3E-05 | 0.0766 | 4.236631 | 0.3243653 | 15.08 | CYP27B1 | cytochrome P450 family 27 subfamily B member 1 |
| 1646 | 8.3E-05 | 3.1E-05 | 0.1475 | -4.163658 | -0.614161 | 12.69 | AKR1C2 | aldo-keto reductase family 1 member C2 |
| 1595 | 0.00014 | 5.6E-05 | 0.0356 | 4.027688 | 0.1431795 | 327.67 | CYP51A1 | cytochrome P450 family 51 subfamily A member 1 |
| 57404 | 0.00021 | 8.7E-05 | 0.0354 | -3.925562 | -0.1390876 | 1370.84 | CYP20A1 | cytochrome P450 family 20 subfamily A member 1 |
| 22977 | 0.00032 | 0.00013 | 0.0736 | -3.820808 | -0.2812026 | 28.08 | AKR7A3 | aldo-keto reductase family 7 member A3 |
| 1312 | 0.0004 | 0.00017 | 0.0396 | 3.760564 | 0.1488342 | 1520.67 | COMT | catechol-O-methyltransferase |
| 2950 | 0.00051 | 0.00023 | 0.0498 | 3.68957 | 0.1839048 | 5469.93 | GSTP1 | glutathione S-transferase pi 1 |
| 2877 | 0.00125 | 0.00059 | 0.0657 | -3.437856 | -0.2258914 | 26.71 | GPX2 | glutathione peroxidase 2 |
| 56171 | 0.00136 | 0.00065 | 0.0616 | -3.411965 | -0.2102489 | 16.81 | DNAH7 | dynein axonemal heavy chain 7 |
| 4255 | 0.00146 | 0.0007 | 0.0517 | 3.389985 | 0.1753613 | 515.63 | MGMT | O-6-methylguanine-DNA methyltransferase |
| 1789 | 0.00162 | 0.00078 | 0.0532 | -3.359104 | -0.1786841 | 35.13 | DNMT3B | DNA methyltransferase 3 beta |
| 4301 | 0.00172 | 0.00083 | 0.0771 | -3.341386 | -0.257677 | 369.57 | AFDN | afadin, adherens junction formation factor |
| 2730 | 0.00185 | 0.0009 | 0.0321 | 3.319111 | 0.1064878 | 625.89 | GCLM | glutamate-cysteine ligase modifier subunit |
| 27134 | 0.00225 | 0.00112 | 0.0555 | 3.259767 | 0.1810538 | 273.96 | TJP3 | tight junction protein 3 |
| 642446 | 0.00305 | 0.00156 | 0.2219 | 3.163251 | 0.7020219 | 25.52 | TRIM64B | tripartite motif containing 64B |
| 4035 | 0.00382 | 0.00199 | 0.0612 | 3.091833 | 0.1891055 | 3516.6 | LRP1 | LDL receptor related protein 1 |
| 221 | 0.00617 | 0.00335 | 0.0611 | 2.934004 | 0.1793055 | 2563.68 | ALDH3B1 | aldehyde dehydrogenase 3 family member B1 |
| 285440 | 0.00659 | 0.0036 | 0.0464 | -2.911417 | -0.1351189 | 742.18 | CYP4V2 | cytochrome P450 family 4 subfamily V member 2 |
| 114112 | 0.00797 | 0.00442 | 0.0645 | -2.846306 | -0.1835776 | 42.37 | TXNRD3 | thioredoxin reductase 3 |
| 2944 | 0.00908 | 0.00511 | 0.2022 | 2.800128 | 0.5661763 | 470.46 | GSTM1 | glutathione S-transferase mu 1 |
| 1573 | 0.0147 | 0.00868 | 0.1056 | -2.624482 | -0.2771875 | 17.2 | CYP2J2 | cytochrome P450 family 2 subfamily J member 2 |
| 7450 | 0.0168 | 0.00997 | 0.1099 | 2.576928 | 0.2831345 | 187.19 | VWF | von Willebrand factor |
| 6820 | 0.0191 | 0.0115 | 0.1064 | -2.527844 | -0.268863 | 12.99 | SULT2B1 | sulfotransferase family 2B member 1 |
| 58494 | 0.0391 | 0.0251 | 0.0697 | 2.239379 | 0.1561444 | 33.32 | JAM2 | junctional adhesion molecule 2 |
| 4851 | 0.0404 | 0.0261 | 0.0487 | 2.2246 | 0.1083441 | 10355.52 | NOTCH1 | notch receptor 1 |
| 9414 | 0.0405 | 0.0262 | 0.0536 | 2.223637 | 0.1191769 | 565.93 | TJP2 | tight junction protein 2 |

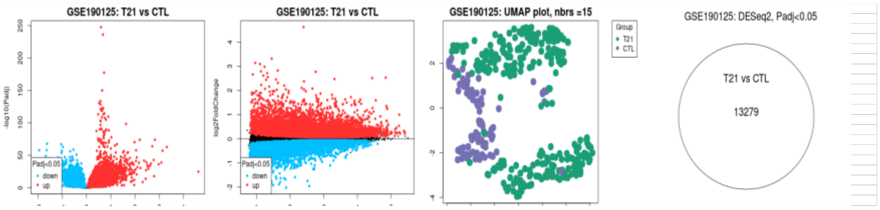

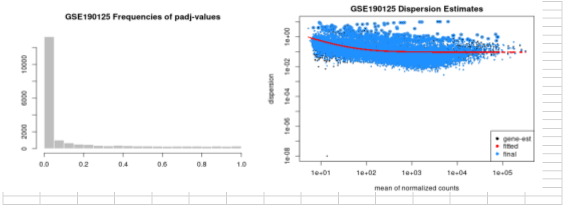

Supplementary Table S22 - Phase 2, tissue study

Genes_of_interest_ADFVSADM_Known_lncRNA_differential_expression

| Gene name | Log 2F | P value | Q value |
|---|---|---|---|
| DNAH5 | 9.814 | 3.92E-08 | 3.89E-06 |

Genes_of_interest_ADFVSADM_Novel_lncRNA_differential_expression

| Gene name | Log 2F | P value | Q value |
|---|---|---|---|
| MT2A | 18.35 | 2.821E-12 | 5.086E-09 |

Genes_of_interest_ADFVSADM_Transcript_differential_expression

| Gene name | Log 2F | P value | Q value |
|---|---|---|---|
| CFTR | -13.34 | 3.595E-12 | 5.584E-09 |
| APC | -13.04 | 4.56E-12 | 6.57E-09 |
| ABCC1 | 14.19 | 5.073E-11 | 2.687E-08 |
| ABCC1 | -11.92 | 5.887E-11 | 2.912E-08 |
| NOTCH1 | -11.42 | 8.219E-11 | 3.642E-08 |
| ALDH3A2 | -11.66 | 5.111E-10 | 1.177E-07 |
| GSS | -12.35 | 5.229E-10 | 1.194E-07 |
| TXNRD1 | 11.6 | 7.183E-10 | 1.493E-07 |
| SULT1A3 | -12.99 | 7.28E-10 | 1.506E-07 |
| CYP2U1 | 11.14 | 8.856E-10 | 1.704E-07 |
| ABCC1 | 10.84 | 2.244E-09 | 3.307E-07 |
| GCLM | 11.62 | 3.817E-09 | 5.281E-07 |
| VDR | 10.92 | 7.976E-09 | 0.000001009 |
| MTHFR | 11.37 | 2.494E-08 | 0.000002687 |
| GPX2 | 13.13 | 0.000003886 | 0.0003531 |
| TXNRD2 | 12.88 | 0.00001585 | 0.001401 |
| TXNRD1 | -10.05 | 0.00003103 | 0.002699 |
| ALDH1L1 | 12.31 | 0.00004736 | 0.004061 |
| CYP2D6 | 3.138 | 0.0002201 | 0.01748 |

Genes_of_interest_ADFVSCTL_Gene_differential_expression

| Gene name | Log 2F | P value | Q value |
|---|---|---|---|
| CYP7A1 | 4.409 | 7.025E-10 | 7.42E-06 |
| NOTUM | 4.409 | 2.429E-07 | 0.001173 |

Genes_of_interest_ADFVSCTL_Novel_lncRNA_differential_expression

| Gene name | Log 2F | P value | Q value |
|---|---|---|---|
| MT2A | 18.35 | 9.955E-17 | 2.192E-13 |

Genes_of_interest_ADFVSCTL_Transcript_differential_expression

| Gene name | Log 2F | P value | Q value |
|---|---|---|---|
| APC | -13.41 | 5.514E-16 | 9.05E-13 |

| | | | |
|---|---|---|---|
| TXNRD1 | -13.53 | 4.097E-14 | 2.376E-11 |
| CFTR | -12.24 | 9.441E-14 | 4.404E-11 |
| STS | -10.7 | 9.544E-14 | 4.42E-11 |
| TXNRD1 | -12.76 | 1.678E-13 | 6.62E-11 |
| CYP2U1 | 11.14 | 2.23E-13 | 8.185E-11 |
| ABCB1 | -12.23 | 3.216E-13 | 1.064E-10 |
| ABCC1 | -12.57 | 4.158E-13 | 1.285E-10 |
| SULT1A3 | -12.81 | 7.887E-13 | 2.118E-10 |
| CYP17A1 | -13.21 | 1.617E-12 | 3.634E-10 |
| SLC25A20 | -11.85 | 3.547E-12 | 6.662E-10 |
| EPHX2 | -11.25 | 3.762E-12 | 6.948E-10 |
| ALDH3B1 | -11.44 | 1.552E-11 | 2.134E-09 |
| ALDH3A2 | -10.17 | 1.7E-11 | 2.293E-09 |
| PRNP | 10.81 | 2.023E-11 | 2.638E-09 |
| TCN2 | -11.08 | 2.657E-11 | 3.258E-09 |
| GRIN2A | -8.278 | 2.977E-11 | 3.567E-09 |
| CFTR | -9.057 | 3.31E-11 | 3.878E-09 |
| TJP3 | 10.05 | 5.87E-11 | 6.163E-09 |
| CYP7A1 | 4.409 | 0.000001039 | 0.00008464 |
| ALDH3B1 | -13.72 | 0.000002559 | 0.0002068 |
| NOTUM | 4.488 | 0.000046 | 0.003444 |
| COMT | 4.116 | 0.0005541 | 0.03509 |

Genes_of_interest_ADFVSCTLF_Gene_differential_expression

| Gene name | Log 2F | P value | Q value |
|---|---|---|---|
| CYP7A1 | 4.344 | 7.211E-07 | 0.004885 |

Genes_of_interest_ADFVSCTLF_Novel_lncRNA_differential_expression

| Gene name | Log 2F | P value | Q value |
|---|---|---|---|
| MT2A | 18.35 | 2.797E-12 | 3.785E-09 |

Genes_of_interest_ADFVSCTLF_Transcript_differential_expression

| Gene name | Log 2F | P value | Q value |
|---|---|---|---|
| TXNRD1 | -14.26 | 4.974E-12 | 5.611E-09 |
| NOTCH1 | 12.77 | 1.539E-11 | 1.099E-08 |
| ABCB1 | -12.97 | 2.632E-11 | 1.599E-08 |
| ABCC1 | -12.19 | 4.066E-11 | 2.067E-08 |
| TXNRD1 | -13.38 | 7.657E-11 | 3.075E-08 |
| SLC25A20 | -12.14 | 3.654E-10 | 8.535E-08 |
| NOTCH1 | 10.88 | 8.198E-10 | 1.559E-07 |
| CYP2U1 | 11.14 | 8.623E-10 | 1.612E-07 |
| TCN2 | -11.82 | 1.015E-09 | 1.783E-07 |
| GRIN2A | -9.015 | 1.104E-09 | 1.891E-07 |

| | | | |
|---|---|---|---|
| COMT | -10.56 | 1.767E-09 | 2.667E-07 |
| ABCC1 | 10.88 | 6.946E-09 | 8.746E-07 |
| VDR | 10.92 | 1.156E-08 | 0.000001375 |
| PRNP | 10.81 | 2.438E-08 | 0.000002564 |
| AOC2 | 10.52 | 2.807E-08 | 0.000002896 |
| TJP3 | 10.05 | 4.778E-08 | 0.000004478 |
| CYP2D6 | 3.72 | 0.0001411 | 0.01135 |
| CYP7A1 | 4.344 | 0.0002629 | 0.02041 |

Genes_of_interest_ADFVSCTLM_Gene_differential_expression

| Gene name | Log 2F | P value | Q value |
|---|---|---|---|
| CYP1A1 | 4.302 | 2.426E-08 | 5.451E-05 |
| SULT1E1 | 12.54 | 3.76E-08 | 0.0000714 |
| PTGIS | -2.848 | 5.326E-07 | 0.0004661 |
| CYP7A1 | 4.511 | 0.000001136 | 0.0008057 |
| VWF | -2.667 | 0.000003261 | 0.001608 |
| NOTUM | 4.619 | 0.000008505 | 0.003215 |
| CYP2E1 | 2.446 | 0.00001263 | 0.004366 |
| CYP1A2 | 3.702 | 0.00009259 | 0.01842 |
| CBS | 1.613 | 0.0001662 | 0.0259 |
| ALDH1L1 | 2.553 | 0.0002799 | 0.03567 |
| FMO3 | 1.511 | 0.0003902 | 0.043 |
| CYP2A13 | 10.45 | 0.0004294 | 0.04499 |

Genes_of_interest_ADFVSCTLM_Known_lncRNA_differential_expression

| Gene name | Log 2F | P value | Q value |
|---|---|---|---|
| CYP4F12 | 14.55 | 2.613E-09 | 1.236E-06 |
| CYP3A7 | -3.15 | 0.0006751 | 0.04623 |

Genes_of_interest_ADFVSCTLM_Novel_lncRNA_differential_expression

| Gene name | Log 2F | P value | Q value |
|---|---|---|---|
| MT2A | 18.35 | 1.328E-09 | 8.674E-07 |

Genes_of_interest_ADFVSCTLM_Transcript_differential_expression

| Gene name | Log 2F | P value | Q value |
|---|---|---|---|
| APC | -14.73 | 1.179E-10 | 2.303E-07 |
| CYP17A1 | -14.36 | 1.347E-09 | 8.731E-07 |
| ABCC1 | -13 | 1.457E-09 | 0.000000908 |
| GSTO1 | 13.96 | 1.576E-08 | 0.000005169 |
| CFTR | -13.56 | 3.297E-08 | 0.000009347 |
| CYP2U1 | 11.14 | 1.068E-07 | 0.00002404 |
| TXNRD1 | 11.6 | 0.000000353 | 0.00006431 |

| | | | |
|---|---|---|---|
| MTHFR | 3.118 | 3.783E-07 | 0.00006781 |
| CYP1A1 | 5.068 | 4.428E-07 | 0.00007703 |
| GSTA2 | 14.31 | 6.654E-07 | 0.0001044 |
| ATP7B | 10.29 | 8.607E-07 | 0.0001271 |
| DNMT3B | 10.18 | 8.851E-07 | 0.0001296 |
| PRNP | 10.81 | 0.000001222 | 0.000164 |
| ALDH3B1 | -13.76 | 0.000001592 | 0.0001968 |
| STS | 9.128 | 0.000001761 | 0.0002127 |
| TJP3 | 10.05 | 0.000002184 | 0.0002494 |
| CYP2A7 | 13.61 | 0.00001002 | 0.0009963 |
| CLDN5 | 4.375 | 0.00001103 | 0.00109 |
| PTGIS | -2.856 | 0.0000344 | 0.003176 |
| ALDH3B1 | -12.76 | 0.00007432 | 0.006495 |
| ALDH1L1 | 12.31 | 0.0001703 | 0.01383 |
| VWF | -2.672 | 0.0003555 | 0.02667 |
| CYP3A4 | 3.788 | 0.0004877 | 0.03506 |
| SULT1E1 | 12.58 | 0.000641 | 0.04429 |

Genes_of_interest_ADMVSADF_Known_lncRNA_differential_expression

| Gene name | Log 2F | P value | Q value |
|---|---|---|---|
| DNAH5 | -9.814 | 1.14E-08 | 1.153E-06 |

Genes_of_interest_ADMVSADF_Novel_lncRNA_differential_expression

| Gene name | Log 2F | P value | Q value |
|---|---|---|---|
| MT2A | -18.35 | 7.705E-13 | 1.831E-09 |

Genes_of_interest_ADMVSADF_Transcript_differential_expression

| Gene name | Log 2F | P value | Q value |
|---|---|---|---|
| ABCC1 | -14.19 | 3.126E-12 | 4.992E-09 |
| APC | 13.04 | 5.206E-12 | 6.44E-09 |
| CFTR | 13.34 | 2.039E-11 | 1.429E-08 |
| TXNRD1 | -11.6 | 2.005E-10 | 6.304E-08 |
| ABCC1 | 11.92 | 2.202E-10 | 6.738E-08 |
| CYP2U1 | -11.14 | 2.472E-10 | 7.219E-08 |
| NOTCH1 | 11.42 | 3.049E-10 | 8.338E-08 |
| ABCC1 | -10.84 | 6.304E-10 | 1.438E-07 |
| GCLM | -11.62 | 1.082E-09 | 2.063E-07 |
| ALDH3A2 | 11.66 | 1.548E-09 | 2.623E-07 |
| VDR | -10.92 | 2.285E-09 | 3.483E-07 |
| SULT1A3 | 12.99 | 2.362E-09 | 3.552E-07 |
| GSS | 12.35 | 1.137E-08 | 0.000001152 |
| MTHFR | -11.37 | 2.732E-08 | 0.000002592 |
| GPX2 | -13.13 | 0.000003886 | 0.0003531 |

| | | | |
|---|---|---|---|
| TXNRD2 | -12.88 | 0.00001585 | 0.001401 |
| TXNRD1 | 10.05 | 0.00003103 | 0.002699 |
| ALDH1L1 | -12.31 | 0.00004736 | 0.004062 |
| CYP2D6 | -3.138 | 0.0002201 | 0.01748 |

Genes_of_interest_ADMVSCTL_Gene_differential_expression

| Gene name | Log 2F | P value | Q value |
|---|---|---|---|
| CYP7A1 | 6.47 | 3.25E-11 | 1.372E-06 |
| CYP4F22 | 3.854 | 0.00000397 | 0.002501 |
| CYP26B1 | -2.779 | 8.938E-06 | 0.004387 |
| NOTUM | 4.143 | 9.279E-06 | 0.004502 |
| CYP2S1 | -2.139 | 0.00001045 | 0.004958 |
| MT1A | -2.091 | 0.0001261 | 0.02554 |

Genes_of_interest_ADMVSCTL_Known_lncRNA_differential_expression

| Gene name | Log 2F | P value | Q value |
|---|---|---|---|
| CYP2C8 | -3.431 | 0.00001651 | 0.001086 |
| DNAH5 | -9.294 | 0.0001547 | 0.009025 |
| ABCC1 | 2.614 | 0.0004313 | 0.02247 |
| SLC22A5 | 1.917 | 0.0007985 | 0.03826 |

Genes_of_interest_ADMVSCTL_Transcript_differential_expression

| Gene name | Log 2F | P value | Q value |
|---|---|---|---|
| ABCC1 | -14.95 | 2.683E-19 | 1.166E-14 |
| TXNRD1 | -13.53 | 6.24E-12 | 3.013E-09 |
| NOTCH1 | 11.42 | 8.292E-12 | 3.66E-09 |
| STS | -10.7 | 1.342E-11 | 4.943E-09 |
| CYP17A1 | -13.21 | 2.637E-11 | 7.794E-09 |
| ABCB1 | -12.23 | 3.445E-11 | 9.385E-09 |
| TJP3 | 12.22 | 4.674E-11 | 1.161E-08 |
| PRNP | 11.67 | 8.362E-11 | 1.775E-08 |
| EPHX2 | -11.25 | 2.475E-10 | 3.695E-08 |
| SLC25A20 | -11.85 | 3.106E-10 | 4.274E-08 |
| ADSL | 10.67 | 3.287E-10 | 4.452E-08 |
| GSS | 12.35 | 4.053E-10 | 5.079E-08 |
| KIF17 | 10.1 | 5.438E-10 | 6.327E-08 |
| MTHFR | -11.06 | 1.28E-09 | 1.284E-07 |
| CFTR | -9.057 | 1.533E-09 | 1.488E-07 |
| TCN2 | -11.08 | 1.656E-09 | 1.583E-07 |
| CYP7A1 | 6.47 | 2.642E-07 | 0.00001923 |
| GCLM | -13.27 | 0.000008139 | 0.0005528 |
| GPX2 | -13.79 | 0.00009642 | 0.005799 |
| CYP2C8 | -3.564 | 0.0001654 | 0.009564 |

| | | | |
|---|---|---|---|
| CYP26B1 | -2.819 | 0.0002094 | 0.01184 |
| CYP2C18 | 2.06 | 0.0002239 | 0.01261 |
| CYP2S1 | -2.141 | 0.0003269 | 0.01769 |
| ADSL | -1.756 | 0.0003663 | 0.01951 |
| GPX1 | -2.051 | 0.0003702 | 0.01966 |
| NOTUM | 4.248 | 0.0003861 | 0.02038 |

Genes_of_interest_ADMVSCTLF_Gene_differential_expression

| Gene name | Log 2F | P value | Q value |
|---|---|---|---|
| GPX2 | -2.797 | 0.000005331 | 0.005155 |
| CYP4F22 | 3.883 | 0.00003176 | 0.01679 |
| CYP26B1 | -2.921 | 0.00003538 | 0.01842 |
| CYP7A1 | 6.405 | 0.0000766 | 0.02803 |
| NOTUM | 4.019 | 0.00008412 | 0.02971 |
| MT1A | -2.306 | 0.00009929 | 0.03279 |

Genes_of_interest_ADMVSCTLF_Known_lncRNA_differential_expression

| Gene name | Log 2F | P value | Q value |
|---|---|---|---|
| CYP2C8 | -3.307 | 0.00004635 | 0.003353 |

Genes_of_interest_ADMVSCTLF_Transcript_differential_expression

| Gene name | Log 2F | P value | Q value |
|---|---|---|---|
| ABCC1 | -14.47 | 2.474E-14 | 9.685E-11 |
| STS | -11.43 | 3.192E-11 | 3.593E-08 |
| TXNRD1 | -14.26 | 1.575E-10 | 0.000000118 |
| APC | 13.04 | 1.605E-10 | 0.000000119 |
| CFTR | 13.34 | 3.696E-10 | 2.124E-07 |
| ABCB1 | -12.97 | 6.739E-10 | 3.132E-07 |
| NOTCH1 | 11.99 | 1.66E-09 | 0.000000565 |
| NOTCH1 | 12.36 | 2.822E-09 | 7.718E-07 |
| TXNRD2 | -11.49 | 3.641E-09 | 9.094E-07 |
| SLC25A20 | -12.14 | 6.196E-09 | 0.000001291 |
| NOTCH1 | 11.42 | 1.482E-08 | 0.000002289 |
| MTHFR | -11.77 | 1.503E-08 | 0.00000231 |
| TCN2 | -11.82 | 1.672E-08 | 0.000002478 |
| TJP3 | 12.22 | 2.096E-08 | 0.000002834 |
| ALDH3A2 | 11.66 | 2.359E-08 | 0.000003069 |
| GSS | 12.35 | 2.808E-08 | 0.000003418 |
| PRNP | 11.67 | 3.367E-08 | 0.000003841 |
| GPX2 | -14.13 | 7.128E-08 | 0.000006741 |
| ADSL | 10.67 | 9.38E-08 | 0.000008384 |
| CYP7A1 | 6.405 | 0.00008019 | 0.005606 |
| CYP4F22 | 15.8 | 0.000346 | 0.02175 |

| | | | |
|---|---|---|---|
| SULT1E1 | 2.142 | 0.0003573 | 0.0224 |

Genes_of_interest_ADMVSCTLM_Gene_differential_expression

| Gene name | Log 2F | P value | Q value |
|---|---|---|---|
| SULT1E1 | 13.9 | 2.432E-11 | 1.89E-08 |
| CYP26B1 | -2.538 | 4.149E-09 | 0.000001497 |
| VWF | -2.199 | 7.003E-07 | 0.0001044 |
| CBS | 1.953 | 0.000001728 | 0.0002136 |
| CYP2A13 | 11.34 | 0.000003283 | 0.0003568 |
| CYP4F12 | 1.914 | 0.000009481 | 0.0008125 |
| CYP2S1 | -2.467 | 0.000009786 | 0.0008325 |
| PTGIS | -1.737 | 0.00001478 | 0.001175 |
| VDR | -1.983 | 0.00004083 | 0.002538 |
| CYP2B6 | 2.708 | 0.0002122 | 0.00901 |
| AOC2 | 1.743 | 0.0005122 | 0.01752 |
| CYP7A1 | 6.572 | 0.0005152 | 0.01758 |
| CYP17A1 | -5.441 | 0.001 | 0.02864 |
| CYP2W1 | 3.028 | 0.00106 | 0.02978 |
| TCN2 | -1.825 | 0.001174 | 0.03207 |
| CYP27A1 | 2.002 | 0.001204 | 0.03269 |
| CYP1A1 | 2.562 | 0.001244 | 0.0336 |
| CGN | 1.471 | 0.001299 | 0.03464 |
| ATP7B | 1.545 | 0.001879 | 0.04498 |
| ALDH1L1 | 2.068 | 0.002053 | 0.04759 |

Genes_of_interest_ADMVSCTLM_Known_lncRNA_differential_expression

| Gene name | Log 2F | P value | Q value |
|---|---|---|---|
| CYP4F12 | 15.75 | 3.702E-12 | 1.294E-08 |
| CYP2C8 | 4.953 | 0.000007482 | 0.0006291 |

Genes_of_interest_ADMVSCTLM_Transcript_differential_expression

| Gene name | Log 2F | P value | Q value |
|---|---|---|---|
| ABCC1 | -15.46 | 8.248E-13 | 4.533E-09 |
| CYP17A1 | -14.36 | 3.252E-09 | 0.000001622 |
| SULT1E1 | 13.97 | 7.295E-09 | 0.000002339 |
| EPHX2 | -12.58 | 8.657E-09 | 0.000002619 |
| NOTCH1 | 11.42 | 2.996E-07 | 0.0000552 |
| ATP7B | 12.07 | 3.593E-07 | 0.00006425 |
| UGT2A1 | 12.5 | 5.599E-07 | 0.00008868 |
| GSS | 12.35 | 0.000001059 | 0.0001401 |
| SULT1A3 | 12.99 | 0.000001189 | 0.0001537 |
| PRNP | 11.67 | 0.000001234 | 0.0001577 |
| TJP3 | 12.22 | 0.000002588 | 0.0002664 |

| | | | |
|---|---|---|---|
| ADSL | 10.67 | 0.000002948 | 0.0002924 |
| VDR | 11.3 | 0.000003001 | 0.0002951 |
| CYP2A13 | 11.34 | 0.000003641 | 0.0003399 |
| CYP3A4 | 5.296 | 0.000004367 | 0.0003871 |
| TXNRD2 | -12.07 | 0.00000444 | 0.0003908 |
| GCLM | -13.06 | 0.000005333 | 0.0004636 |
| KIF17 | 10.1 | 0.00002102 | 0.001615 |
| CYP26B1 | -2.58 | 0.00002677 | 0.002009 |
| GPX1 | -2.563 | 0.00003982 | 0.002898 |
| PTGS1 | -11.96 | 0.00004992 | 0.003537 |
| PTGS1 | -11.57 | 0.00007496 | 0.005087 |
| TXNRD2 | 4.48 | 0.0001148 | 0.007465 |
| VDR | -10.94 | 0.0001283 | 0.008199 |
| CYP3A4 | 4.889 | 0.0001637 | 0.01011 |
| MTHFR | -9.412 | 0.000336 | 0.0188 |
| CBS | 4.394 | 0.0003708 | 0.02042 |
| CYP17A1 | -7.184 | 0.0003738 | 0.02056 |
| CYP7A1 | 6.572 | 0.0005142 | 0.02682 |
| CYP1A1 | 3.571 | 0.0006767 | 0.03357 |
| CYP2A7 | 15.38 | 0.0009761 | 0.04509 |

Genes_of_interest_ADVSCTL_Gene_differential_expression

| Gene name | Log 2F | P value | Q value |
|---|---|---|---|
| CYP7A1 | 5.647 | 1.423E-10 | 1.545E-06 |
| NOTUM | 4.301 | 2.808E-07 | 0.001179 |
| CYP4F22 | 2.996 | 0.00005591 | 0.02791 |
| CYP2S1 | -1.774 | 0.00006244 | 0.02994 |
| CYP17A1 | -4.465 | 0.0001458 | 0.0449 |

Genes_of_interest_ADVSCTL_Known_lncRNA_differential_expression

| Gene name | Log 2F | P value | Q value |
|---|---|---|---|
| ABCC1 | 2.294 | 4.44E-05 | 0.003967 |

Genes_of_interest_ADVSCTL_Novel_lncRNA_differential_expression

| Gene name | Log 2F | P value | Q value |
|---|---|---|---|
| MT2A | 17.55 | 4.107E-17 | 5.166E-14 |

Genes_of_interest_ADVSCTL_Transcript_differential_expression

| Gene name | Log 2F | P value | Q value |
|---|---|---|---|
| TXNRD1 | -13.53 | 1.152E-16 | 1.198E-13 |
| STS | -10.7 | 1.31E-16 | 1.302E-13 |
| CYP17A1 | -13.21 | 5.747E-16 | 3.893E-13 |
| ABCB1 | -12.23 | 7.623E-16 | 4.95E-13 |

| | | | |
|---|---|---|---|
| SLC25A20 | -11.85 | 1.31E-14 | 4.264E-12 |
| EPHX2 | -11.25 | 1.38E-14 | 4.379E-12 |
| TCN2 | -11.08 | 1.419E-13 | 2.805E-11 |
| CFTR | -9.057 | 1.813E-13 | 3.408E-11 |
| NOTCH1 | 10.19 | 2.553E-13 | 4.556E-11 |
| TJP2 | 10.64 | 5.151E-13 | 8.091E-11 |
| TJP3 | 11.37 | 7.337E-13 | 1.084E-10 |
| PRNP | 11.24 | 8.218E-13 | 1.195E-10 |
| GSS | 11.13 | 3.068E-12 | 3.734E-10 |
| ABCG2 | 9.103 | 3.411E-11 | 3.537E-09 |
| CYP7A1 | 5.647 | 1.082E-07 | 0.00001076 |
| NOTUM | 4.39 | 0.00001585 | 0.001483 |
| CYP2S1 | -1.796 | 0.00007904 | 0.006842 |
| GPX4 | 2.823 | 0.0006082 | 0.04334 |

Supplementary Table S23

#### Comprehensive Differentially Expressed Genes (DGE) Comparison between GEO and Tissue Studies

| Total Unique Genes (162) | GEO Study (154) | Tissue Study (69) | Shared Genes (61) |
|---|---|---|---|
| ABCB1 | ABCB1 | ABCB1 | ABCB1 |
| ABCC1 | ABCC1 | ABCC1 | ABCC1 |
| ABCC2 | ABCC2 | ABCG2 | ABCG2 |
| ABCG2 | ABCG2 | ADSL | ADSL |

| | | | |
|---|---|---|---|
| ADSL | ADSL | ALDH1L1 | ALDH3A2 |
| AFDN | AFDN | ALDH3A2 | ALDH3B1 |
| AKR1C2 | AKR1C2 | ALDH3B1 | AOC2 |
| AKR7A3 | AKR7A3 | AOC2 | APC |
| ALAD | ALAD | APC | ATP7B |
| ALDH1A1 | ALDH1A1 | ATP7B | CBS |
| ALDH1L1 | ALDH3A2 | CBS | CFTR |
| ALDH3A2 | ALDH3B1 | CFTR | CGN |
| ALDH3B1 | ALOX15 | CGN | CLDN5 |
| ALOX15 | AOC2 | CLDN5 | COMT |
| AOC2 | AOX1 | COMT | CYP1A1 |
| AOX1 | APC | CYP17A1 | CYP26B1 |
| APC | APOE | CYP1A1 | CYP27A1 |
| APOE | ARSG | CYP1A2 | CYP2A7 |
| ARSG | ATP7B | CYP26B1 | CYP2C18 |
| ATP7B | BDNF | CYP27A1 | CYP2C8 |
| BDNF | CBS | CYP2A13 | CYP2D6 |
| CBS | CES3 | CYP2A7 | CYP2E1 |
| CES3 | CFTR | CYP2B6 | CYP2S1 |
| CFTR | CGN | CYP2C18 | CYP2U1 |
| CGN | CHST12 | CYP2C8 | CYP3A4 |
| CHST12 | CHST14 | CYP2D6 | CYP3A7 |
| CHST14 | CHST5 | CYP2E1 | CYP4F12 |
| CHST5 | CLDN3 | CYP2S1 | CYP4F22 |
| CLDN3 | CLDN5 | CYP2U1 | CYP7A1 |
| CLDN5 | COMT | CYP2W1 | DNAH5 |
| COMT | CPOX | CYP3A4 | DNMT3B |
| CPOX | CYP11A1 | CYP3A7 | EPHX2 |
| CYP11A1 | CYP19A1 | CYP4F12 | FMO3 |
| CYP17A1 | CYP1A1 | CYP4F22 | GCLM |
| CYP19A1 | CYP1B1 | CYP7A1 | GPX1 |
| CYP1A1 | CYP20A1 | DNAH5 | GPX2 |
| CYP1A2 | CYP21A2 | DNMT3B | GPX4 |
| CYP1B1 | CYP24A1 | EPHX2 | GRIN2A |
| CYP20A1 | CYP26A1 | FMO3 | GSS |
| CYP21A2 | CYP26B1 | GCLM | GSTO1 |
| CYP24A1 | CYP26C1 | GPX1 | KIF17 |
| CYP26A1 | CYP27A1 | GPX2 | MT1A |
| CYP26B1 | CYP27B1 | GPX4 | MT2A |
| CYP26C1 | CYP27C1 | GRIN2A | MTHFR |
| CYP27A1 | CYP2A6 | GSS | NOTCH1 |
| CYP27B1 | CYP2A7 | GSTA2 | NOTUM |
| CYP27C1 | CYP2C18 | GSTO1 | PRNP |
| CYP2A13 | CYP2C8 | KIF17 | PTGIS |

CYP2A6
CYP2A7
CYP2B6
CYP2C18
CYP2C8
CYP2D6
CYP2E1
CYP2J2
CYP2R1
CYP2S1
CYP2U1
CYP2W1
CYP39A1
CYP3A4
CYP3A5
CYP3A7
CYP46A1
CYP4A11
CYP4F11
CYP4F12
CYP4F22
CYP4F3
CYP4V2
CYP4X1
CYP51A1
CYP7A1
CYP7B1
DNAH11
DNAH5
DNAH7
DNMT3B
DRD1
EPHX1
EPHX2
EPHX4
FMO3
FOXO3
GCLC
GCLM
GPX1
GPX2
GPX4
GRIN2A
GRIN2B

CYP2D6
CYP2E1
CYP2J2
CYP2R1
CYP2S1
CYP2U1
CYP39A1
CYP3A4
CYP3A5
CYP3A7
CYP46A1
CYP4A11
CYP4F11
CYP4F12
CYP4F22
CYP4F3
CYP4V2
CYP4X1
CYP51A1
CYP7A1
CYP7B1
DNAH11
DNAH5
DNAH7
DNMT3B
DRD1
EPHX1
EPHX2
EPHX4
FMO3
FOXO3
GCLC
GCLM
GPX1
GPX2
GPX4
GRIN2A
GRIN2B
GSS
GSTM1
GSTM4
GSTO1
GSTP1
GSTT2

MT1A
MT2A
MTHFR
NOTCH1
NOTUM
PRNP
PTGIS
PTGS1
SLC22A5
SLC25A20
STS
SULT1A3
SULT1E1
TCN2
TJP2
TJP3
TXNRD1
TXNRD2
UGT2A1
VDR
VWF

PTGS1
SLC22A5
SLC25A20
STS
SULT1A3
SULT1E1
TCN2
TJP2
TJP3
TXNRD1
TXNRD2
VDR
VWF

GSS
GSTA2
GSTM1
GSTM4
GSTO1
GSTP1
GSTT2
GSTT2B
GUSB
HFE
HSD17B1
IFT74
JAM2
KIF17
KIFC1
KLHL1
LOXL4
LRP1
MGMT
MT1A
MT1M
MT2A
MTHFR
NAT2
NOS2
NOTCH1
NOTCH3
NOTUM
PRNP
PTGES
PTGIS
PTGS1
RELN
SGF29
SLC16A1
SLC22A5
SLC25A20
SLC2A6
SLC6A4
SLC7A5
SLX1A
SLX1B
STS
SULT1A1

GSTT2B
GUSB
HFE
HSD17B1
IFT74
JAM2
KIF17
KIFC1
KLHL1
LOXL4
LRP1
MGMT
MT1A
MT1M
MT2A
MTHFR
NAT2
NOS2
NOTCH1
NOTCH3
NOTUM
PRNP
PTGES
PTGIS
PTGS1
RELN
SGF29
SLC16A1
SLC22A5
SLC25A20
SLC2A6
SLC6A4
SLC7A5
SLX1A
SLX1B
STS
SULT1A1
SULT1A2
SULT1A3
SULT1A4
SULT1B1
SULT1C2
SULT1C4
SULT1E1

SULT1A2
SULT1A3
SULT1A4
SULT1B1
SULT1C2
SULT1C4
SULT1E1
SULT2B1
SULT4A1
SUOX
SYCE1L
TCN2
TDO2
TJP1
TJP2
TJP3
TRIM64B
TXNRD1
TXNRD2
TXNRD3
UGT2A1
UGT2B11
UGT2B4
VDR
VWF
XDH

SULT2B1
SULT4A1
SUOX
SYCE1L
TCN2
TDO2
TJP1
TJP2
TJP3
TRIM64B
TXNRD1
TXNRD2
TXNRD3
UGT2B11
UGT2B4
VDR
VWF
XDH

Summary Statistics:

- Total unique genes across both studies: 162
- Genes identified in GEO study: 154
- Genes identified in Tissue study: 69
- Genes found in both studies: 61
- Genes unique to GEO study: 93
- Genes unique to Tissue study: 8

**Tissue study methods - details.**

Despite the challenges in procuring liver tissue from neurologically afflicted patients, we identified such tissue and purchased tissue from Amsbio, in Cambridge, Massachusetts, from seven AD patients, and five non-AD controls, all deceased. The research was conducted in accordance with prevailing legal requirements, following the rules of the Declaration of Helsinki of 1975 and as reviewed most recently in 2024.

I assessed the need for IRB approval. Under the United States Code of Regulations, Title 45, up to date as of 3/22/2024, this project falls into an exempt category since "The information obtained is recorded by the investigator in such a manner that the identity of the human subjects cannot readily be ascertained, directly or through identifiers linked to the subjects" as noted in Code of Federal Regulations 46.104 exempt research section d, (2) (i) (https://www.ecfr.gov/current/title-45/subtitle-A/subchapter-A/part-46/subpart-A/section-46.104). The samples did not include patient identifiers. I determined that IRB approval was not required due to the lack of patient identifiers, the lack of any attempt to identify patients by persons associated with our investigations, and the samples being acquired from deceased individuals. The samples were collected in Russia, an ethics committee (the Russian equivalent of an IRB) approving the collections. In an Amsbio 12/20/22 email, the consent forms utilized for this data collection, and the ethics committee review form are available below. Liver tissue metadata is also available. Documentation of the procurement process is included below.

*Tissue collection and consent forms*

### Consent form for data collection

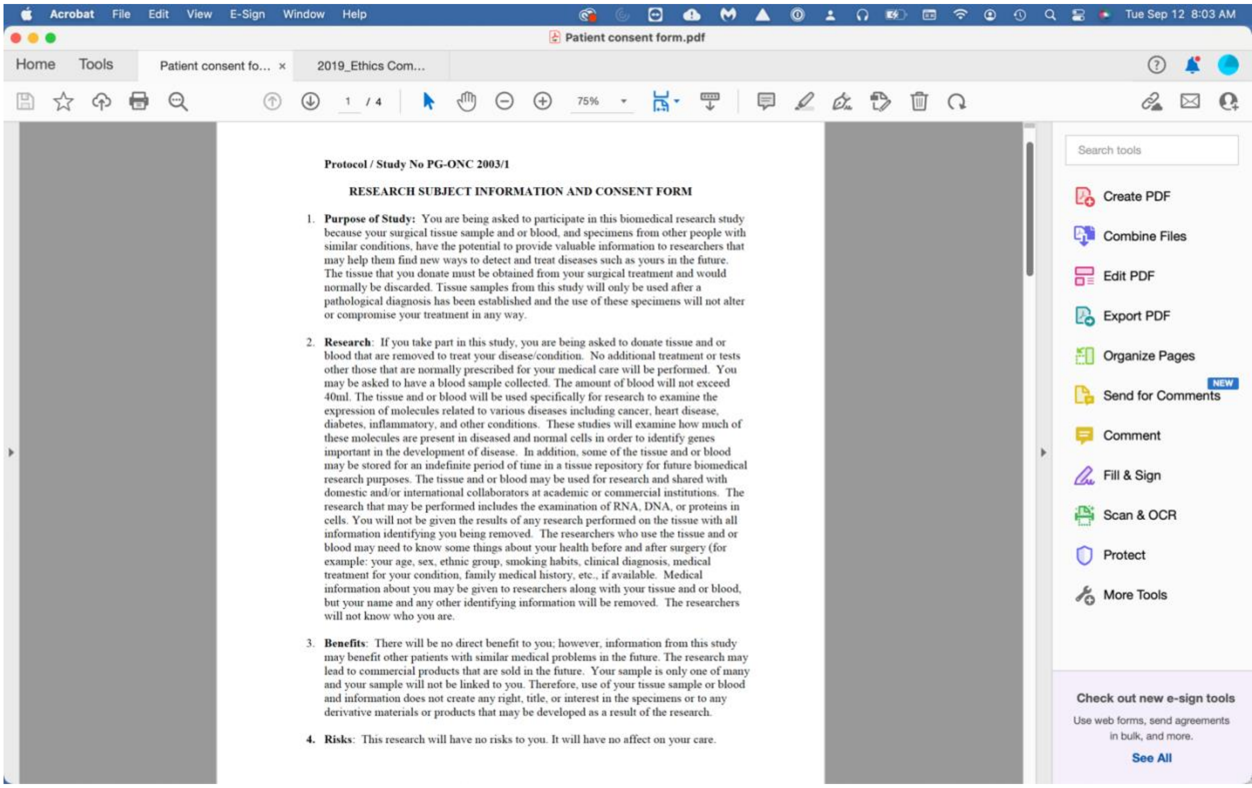

**Protocol / Study No PG-ONC 2003/1**

**RESEARCH SUBJECT INFORMATION AND CONSENT FORM**

1. **Purpose of Study:** You are being asked to participate in this biomedical research study because your surgical tissue sample and or blood, and specimens from other people with similar conditions, have the potential to provide valuable information to researchers that may help them find new ways to detect and treat diseases such as yours in the future. The tissue that you donate must be obtained from your surgical treatment and would normally be discarded. Tissue samples from this study will only be used after a pathological diagnosis has been established and the use of these specimens will not alter or compromise your treatment in any way.

2. **Research**: If you take part in this study, you are being asked to donate tissue and or blood that are removed to treat your disease/condition. No additional treatment or tests other those that are normally prescribed for your medical care will be performed. You may be asked to have a blood sample collected. The amount of blood will not exceed 40ml. The tissue and or blood will be used specifically for research to examine the expression of molecules related to various diseases including cancer, heart disease, diabetes, inflammatory, and other conditions. These studies will examine how much of these molecules are present in diseased and normal cells in order to identify genes important in the development of disease. In addition, some of the tissue and or blood may be stored for an indefinite period of time in a tissue repository for future biomedical research purposes. The tissue and or blood may be used for research and shared with domestic and/or international collaborators at academic or commercial institutions. The research that may be performed includes the examination of RNA, DNA, or proteins in cells. You will not be given the results of any research performed on the tissue with all information identifying you being removed. The researchers who use the tissue and or blood may need to know some things about your health before and after surgery (for example: your age, sex, ethnic group, smoking habits, clinical diagnosis, medical treatment for your condition, family medical history, etc., if available. Medical information about you may be given to researchers along with your tissue and or blood, but your name and any other identifying information will be removed. The researchers will not know who you are.

3. **Benefits**: There will be no direct benefit to you; however, information from this study may benefit other patients with similar medical problems in the future. The research may lead to commercial products that are sold in the future. Your sample is only one of many and your sample will not be linked to you. Therefore, use of your tissue sample or blood and information does not create any right, title, or interest in the specimens or to any derivative materials or products that may be developed as a result of the research.

4. **Risks**: This research will have no risks to you. It will have no affect on your care.

5. **Voluntary Participation/Withdrawal**: Taking part in this study is voluntary. You may choose not to take part, or if you decide to take part, you can later change your mind and withdraw from the study. Your decision will not change the present or future health care or other services that you receive. The study doctor, or the sponsor, may stop your participation in this study without your consent. Whether or not you choose to take part in the study, you will receive the standard treatment for patients with your condition.

6. **Costs**: There will be no additional costs for this procedure.

7. **Compensation**: You will not be paid for participating in this study.

8. **Confidentiality**: All information collected about you during the course of this study will be kept confidential to the extent permitted by law. Only a code number will identify you in the research records. Information, which identifies you personally, will not be released without your written permission. Your health records may be reviewed by the study sponsor, its agents, the Human Investigation Committee, approved persons from other organizations taking part in this study, and appropriate federal agencies. Information from this study may be published, but your identity will be kept confidential in any publications.

9. **Questions**: This study has been explained to you and all of your questions have been answered. If you have any questions in the future or in the case of a research related injury or illness, you may contact the PI ______________________________ or one of his/her associates at the following telephone number ________________. If you have any questions about your rights as a research subject, the Chair of the Human Investigation Committee can be contacted at ______________________________.

10. **Consent to Participate in a Research Trial**: To voluntarily agree to take part in this study, you must sign on the line below. If you choose to take part in this study, you may withdraw at any time. You are not giving up any of your legal rights by signing this form. Your signature below indicates that you have read, or have been read this entire consent form, including the risks and benefits, and have had all your questions answered. You will be given a copy of this consent form.

| | |
|---|---|
| ______________________________<br>**Signature of Study Patient** | ________________<br>**Date** |
| ______________________________<br>**Printed Name of Study Patient** | |
| ______________________________<br>**Signature of Legally Authorized Representative** | ________________<br>**Date** |

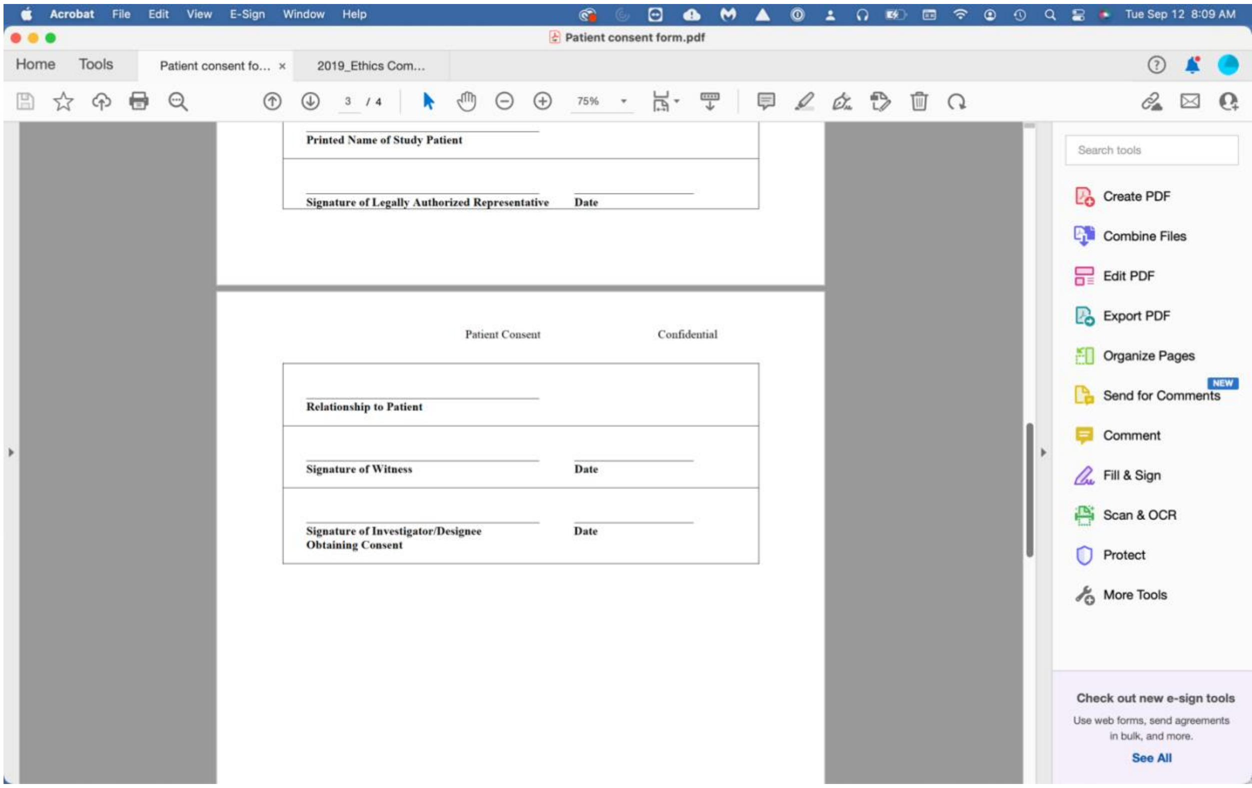

| |
|---|
| Printed Name of Study Patient |
| Signature of Legally Authorized Representative Date |

Patient Consent     Confidential

| |
|---|
| Relationship to Patient |
| Signature of Witness Date |
| Signature of Investigator/Designee Obtaining Consent Date |

The Ethics Committee of the Russian Oncological Research Center n.a. N.N. Blokhin approved  the collection of the samples, which were purchased from AMSBIO.

**RUSSIAN ONCOLOGICAL RESEARCH CENTER n.a. N.N. BLOKH**
**ETHICS COMMITTEE REVIEW FORM**

| Protocol/Study No | PG-ONC 2003/1 |
|---|---|

**Title:** COLLECTION OF TISSUE, BLOOD AND BONE MARROW SAMPLES FOR DIAGNOSTIC AND PHARMACEUTICAL RESEARCH

**developed by** [redacted]

**Following documents have been submitted to the Ethics Committee on date** 20.[illegible]

| | | | |
|---|---|---|---|
| ☑ Protocol | version | | dated |
| ☑ Patient Information/ Consent Form | version | | dated |
| ☐ Investigator's Brochure | version | | dated |
| ☐ Amendment(s) | number | | dated |
| ☐ Insurance | number | | dated |
| ☐ Regulatory approval | protocol | | dated |
| ☑ CV of Investigator(s) | Name | N.A. Dyakova | dated |
| | Name | | dated |

☑ Other (specify) Annual Report

The Continuation of Study was:
- ☑ Approved
- ☐ Conditionally approved (see attached letter)
- ☐ Deferred pending further information (see att[illegible]
- ☐ Rejected (see attached letter)

Is review of study status required ☑ Yes ☐ No

If Yes, ☑ Annually ☐ Periodically (when) ______

The investigator(s) none
is /are EC member(s) and therefore did not vote

**Date:** 9 January 2019

**Chairman of EC** D.Z. Kupchan **Signature** [signature]

| |
|---|
| The EC is organized and operates in compliance with the ICH GCP requirem[illegible] and Russian Law |

Documentation of the tissue acquisition process is available from the author.

**Liver samples metadata.**

| Sample ID | Sex | Age | Ethnicity | Sample type | Tissue type | Diagnosis | PMI, hours | BMI | Cause of death |
|---|---|---|---|---|---|---|---|---|---|
| 091584A(2) | F | 96-100 | Caucasian | FF | liver | AD | 3.5 | 20.2 | Alzheimer's disease, cerebral edema |
| 091499A(1) | F | 76-80 | Caucasian | FF | liver | AD | 2 | 25.5 | Alzheimer's disease, cerebral edema |
| 091486A(2) | M | 81-85 | Caucasian | FF | liver | AD | 4 | 25.7 | Alzheimer's disease, cerebral edema |
| 091485A(3) | M | 85-90 | Caucasian | FF | liver | AD | 4 | 25.6 | Alzheimer's disease, cerebral edema |
| 091483A(3) | M | 81-85 | Caucasian | FF | liver | AD | 3.5 | 25.6 | Alzheimer's disease, cerebral edema |
| 091361A(4) | F | 61-65 | Caucasian | FF | liver | AD | 2.5 | 26.1 | Alzheimer's disease, pneumonia |
| 091260A(4) | F | 71-75 | Caucasian | FF | liver | AD | 2.5 | 20.8 | Alzheimer's disease, cachexia, bronchopneumonia |
| 091216A(2) | M | 86-90 | Caucasian | FF | liver | normal tissue | 3 | 24.8 | heart failure |
| 091207A(5) | F | 81-85 | Caucasian | FF | liver | normal tissue | 2.5 | 25.7 | postinfarction cardiosclerosis, chronic heart failure |
| 091121A(3) | M | 71-75 | Caucasian | FF | liver | normal tissue | 2.5 | 25.9 | cardiac aneurism, heart failure |
| 09968A(6) | F | 71-75 | Caucasian | FF | liver | normal tissue | 4 | 23.0 | PE |
| 09630A(2) | F | 66-70 | Caucasian | FF | liver | normal tissue | 5.5 | N/A | PE |

After acquiring the needed tissue, and ensuring that it was high quality, and after proper shipping to LC Sciences (LCS) in Houston, Texas, RNA-seq according to LCS protocol was carried out, the reports organized, tabulated, and reported here.

The analysis was carried out with comparisons of AD and non-AD tissue, and male/female and combined samples. Significance is expressed in P values which control family-wise error rate and may result in more false negatives, and Q values, which control false discovery rate and are more likely to produce occasional false positives. Given that there may be several splicing isoforms transcripts found in one gene, the result tables report more transcripts than genes.

#### STREGA Reporting Recommendations, Extended from STROBE Statement.

| Item | Item Number | STROBE Guideline | Extension for Genetic Association Studies (STREGA) |
|---|---|---|---|
| **Title and Abstract** | 1 | (a) Indicate the study's design with a commonly used term in the title or the abstract. | |
| | | (b) Provide in the abstract an informative and balanced summary of what was done and what was found. | |
| **Introduction** | | | |
| *Background rationale* | 2 | Explain the scientific background and rationale for the investigation being reported. | |
| *Objectives* | 3 | State specific objectives, including any pre-specified hypotheses. | ***State if the study is the first report of a genetic association, a replication effort, or both.*** |
| **Methods** | | | |
| *Study design* | 4 | Present key elements of study design early in the paper. | |
| *Setting* | 5 | Describe the setting, locations and relevant dates, including periods of recruitment, exposure, follow-up, and data collection. | |
| *Participants* | 6 | (a) **Cohort study** – Give the eligibility criteria, and the sources and methods of selection of participants. Describe methods of follow-up. **Case-control study** – Give the eligibility criteria, and the sources and methods of case ascertainment and control selection. Give the rationale for the choice of cases and controls. **Cross-sectional study** – Give the eligibility criteria, and the sources and methods of selection of participants. | ***Give information on the criteria and methods for selection of subsets of participants from a larger study, when relevant.*** |
| | | (b) **Cohort study** – For matched studies, give matching criteria and number of exposed and unexposed. **Case-control study** – For matched studies, give matching criteria and the number of controls per case. | |
| *Variables* | 7 | (a) Clearly define all outcomes, exposures, predictors, potential confounders, and effect modifiers. Give diagnostic criteria, if applicable. | ***(b) Clearly define genetic exposures (genetic variants) using a widely-used nomenclature system. Identify variables likely to be associated with population stratification (confounding by ethnic origin).*** |
| *Data sources/measurement* | 8* | (a) For each variable of interest, give sources of data and details of methods of assessment (measurement). Describe comparability of assessment methods if there is more than one group. | ***(b) Describe laboratory methods, including source and storage of DNA, genotyping methods and platforms (including the allele calling algorithm used, and its version), error rates and call rates. State the laboratory/centre where genotyping was done. Describe comparability of laboratory methods if there is more than one group. Specify whether genotypes were assigned using all of the data from the study simultaneously or in smaller batches.*** |
| *Bias* | 9 | (a) Describe any efforts to address potential sources of bias. | ***(b) For quantitative outcome variables, specify if any investigation of potential bias resulting from pharmacotherapy was undertaken. If relevant, describe the nature and magnitude of the potential bias, and explain what approach was used to deal with this.*** |
| *Study size* | 10 | Explain how the study size was arrived at. | |
| *Quantitative variables* | 11 | Explain how quantitative variables were handled in the analyses. If applicable, describe which groupings were chosen, and why. | ***If applicable, describe how effects of treatment were dealt with.*** |
| Statistical methods | 12 | (a) Describe all statistical methods, including those used to control for confounding. | ***State software version used and options (or settings) chosen.*** |
| | | (b) Describe any methods used to examine subgroups and interactions. | |
| | | (c) Explain how missing data were addressed. | |
| | | (d) **Cohort study** – If applicable, explain how loss to follow-up was addressed. **Case-control study** – If applicable, explain how matching of cases and controls was addressed. **Cross-sectional study** – If applicable, describe analytical methods taking account of sampling strategy. | |
| | | (e) Describe any sensitivity analyses. | |
| | | | ***(f) State whether Hardy-Weinberg equilibrium was considered and, if so, how.*** |
| | | | ***(g) Describe any methods used for inferring genotypes or haplotypes.*** |
| | | | ***(h) Describe any methods used to assess or address population stratification.*** |
| | | | ***(i) Describe any methods used to address multiple comparisons or to control risk of false positive findings.*** |
| | | | ***(j) Describe any methods used to address and correct for relatedness among subjects*** |
| **Results** | | | |
| *Participants* | 13* | (a) Report the numbers of individuals at each stage of the study – e.g., numbers potentially eligible, examined for eligibility, confirmed eligible, included in the study, completing follow-up, and analysed. | ***Report numbers of individuals in whom genotyping was attempted and numbers of individuals in whom genotyping was successful.*** |
| | | (b) Give reasons for non-participation at each stage. | |
| | | (c) Consider use of a flow diagram. | |
| *Descriptive data* | 14* | (a) Give characteristics of study participants (e.g., demographic, clinical, social) and information on exposures and potential confounders. | ***Consider giving information by genotype.*** |
| | | (b) Indicate the number of participants with missing data for each variable of interest. | |
| | | (c) **Cohort study** – Summarize follow-up time (e.g., average and total amount). | |
| *Outcome data* | 15* | **Cohort study** – Report numbers of outcome events or summary measures over time. | ***Report outcomes (phenotypes) for each genotype category over time*** |
| | | **Case-control study** – Report numbers in each exposure category, or summary measures of exposure. | ***Report numbers in each genotype category*** |
| | | **Cross-sectional study** – Report numbers of outcome events or summary measures. | ***Report outcomes (phenotypes) for each genotype category*** |
| *Main results* | 16 | (a) Give unadjusted estimates and, if applicable, confounder-adjusted estimates and their precision (e.g., 95% confidence intervals). Make clear which confounders were adjusted for and why they were included. | |
| | | (b) Report category boundaries when continuous variables were categorized. | |
| | | (c) If relevant, consider translating estimates of relative risk into absolute risk for a meaningful time period. | |
| | | | ***(d) Report results of any adjustments for multiple comparisons.*** |
| *Other analyses* | 17 | (a) Report other analyses done – e.g., analyses of subgroups and interactions, and sensitivity analyses. | |
| | | | ***(b) If numerous genetic exposures (genetic variants) were examined, summarize results from all analyses undertaken.*** |
| | | | ***(c) If detailed results are available elsewhere, state how they can be accessed.*** |
| **Discussion** | | | |
| *Key results* | 18 | Summarize key results with reference to study objectives. | |
| *Limitations* | 19 | Discuss limitations of the study, taking into account sources of potential bias or imprecision. Discuss both direction and magnitude of any potential bias. | |
| *Interpretation* | 20 | Give a cautious overall interpretation of results considering objectives, limitations, multiplicity of analyses, results from similar studies, and other relevant evidence. | |
| *Generalizability* | 21 | Discuss the generalizability (external validity) of the study results. | |
| **Other Information** | | | |
| *Funding* | 22 | Give the source of funding and the role of the funders for the present study and, if applicable, for the original study on which the present article is based. | |

STREGA = STrengthening the REporting of Genetic Association studies; STROBE = Strengthening the Reporting of Observational Studies in Epidemiology.
* Give information separately for cases and controls in case-control studies and, if applicable, for exposed and unexposed groups in cohort and cross-sectional studies.
doi:10.1371/journal.pmed.1000022.t001

The above table is from reference 29 which contains the following text: "This is an open-access article distributed under the terms of the Creative Commons Public Domain declaration, which stipulates that, once placed in the public domain, this work may be freely reproduced, distributed, transmitted, modified, built upon, or otherwise used by anyone for any lawful purpose."

**Declarations**

Clinical trial number: not applicable

Ethical approval and consent to participate, human ethics: Phase 1 of the study utilized resources of the National Center for Biotechnology Information's Gene Expression Omnibus, for which no additional approval is required. From the NCBI website, "The NCBI places no restrictions on the use or distribution of the GEO data. Unless otherwise stated, documents and files on NCBI Web servers may be freely downloaded and reproduced." For phase 2, see pages 92-98, above.

Consent for publication: not applicable for GEO data, for phase 2 data, see above.

Funding: The research in this study was funded by Daniel Foley, Steamboat Springs, Colorado, in memory of Suzanne Foley. Mr. Foley has no competing interests. There was no outside funding.

Author's contributions: This manuscript was entirely written and organized by Mark E. McCaulley who was the sole author.

Competing interests: none.

Data and materials availability: All data are available in the manuscript, or Supplementary Material. Any additional data concerns or requests may be forwarded to the author.
